# Bayesian generative modeling for heterogeneous wastewater data applied to COVID-19 forecasting

**DOI:** 10.64898/2026.02.23.26346887

**Authors:** Kaitlyn E. Johnson, George Vega Yon, Samuel P.C. Brand, Christian O. Bernal Zelaya, Damon Bayer, Igor Volkov, Zachary Susswein, Andrew Magee, Katelyn M. Gostic, Kayla M. English, Isaac Ghinai, Arran Hamlet, Scott W. Olesen, Juliet R.C. Pulliam, Sam Abbott, Dylan H. Morris

## Abstract

Infectious disease forecasts can inform public health decision-making. Wastewater monitoring is a relatively new epidemiological data source with multiple potential applications, including forecasting. Incorporating wastewater data into epidemiological forecasting models is challenging, and relatively few studies have assessed whether this improves forecast performance. We present and evaluate a semi-mechanistic wastewater-informed forecasting model. The model forecasts COVID-19 hospital admissions at the state and territorial levels in the United States, based on incident hospital admissions data and, optionally, SARS-CoV-2 wastewater concentration data from multiple wastewater sampling sites. From February through April 2024, we produced real-time wastewater-informed COVID-19 forecasts using development versions of the model and submitted them to the United States COVID-19 Forecast Hub (“the Hub”). We then published an open-source R package, `wwinference`, that implements the model with or without wastewater as an input. Using proper scoring rules and measures of model calibration, we assess both our real-time submissions to the Hub and retrospective hypothetical forecasts from `wwinference` made with and without wastewater data. While the models performed similarly with and without the wastewater signal included, there was substantial heterogeneity for individual locations and dates where wastewater data meaningfully improved or degraded the model’s forecast performance. Compared to other models submitted to the Hub during the period spanned by our submissions, the real-time wastewater-informed version of our model ranked fourth of 10 models, with the hospital admissions-only version of our model ranking second out of 10 models. Across the 2023-2024 winter epidemic wave, retrospective forecasts from `wwinference` would have performed similarly with and without the wastewater signal included: fifth and fourth out of 10 models, respectively. To better understand the drivers of differential forecast performance with and without wastewater, we performed an exploratory analysis investigating the relationship between characteristics of the input data and improved and reduced performance in our model. Based on that analysis, we identify and discuss key areas for further model development. To our knowledge, this is the first work that conducts an evaluation of real-time and retrospective infectious disease forecasts across the United States both with and without wastewater data and compared to other forecasting models.

**Author Summary:** Wastewater-based epidemiology, in combination with clinical surveillance, has the potential to improve situational awareness and inform outbreak responses. We developed a model that uses data on the pathogen concentration in wastewater from one or more wastewater treatment plants in combination with hospital admissions to produce short-term forecasts of hospital admissions. We produced and submitted forecasts of 28-day ahead COVID-19 hospital admissions from this model to the U.S. COVID-19 Forecast Hub during the spring of 2024 and found that it performed well in comparison to other models during that limited time period. To assess the added value of incorporating wastewater data into the model and to investigate how it would have performed had we submitted it during the entire 2023-2024 winter epidemic wave, we performed a retrospective analysis in which we produced forecasts from the model with and without including wastewater data, using data that would have been available in real-time as of each forecast date. Both versions of the model would have been median overall performers had they been submitted to the Hub throughout the season. When comparing the model’s performance with and without wastewater data included, we found that overall forecast performance was very similar, with wastewater data slightly reducing overall average forecast performance. Within this result, there was significant heterogeneity, with clear instances of wastewater data improving and detracting from forecast performance. We used trends in the observed data to generate hypotheses as to the drivers of improved and reduced relative forecast performance within our model. We conclude by suggesting future work to improve the model and more broadly the application of wastewater-based epidemiology to forecasting.

## Introduction

Epidemiological forecasts provide quantitative predictions that can be used to inform public health decision-making. During the COVID-19 pandemic, short-term forecasts informed assessments of healthcare staffing needs, school closure policies, and allocation of limited resources such as ventilators and therapeutics (1–3). In April 2020, the United States COVID-19 Forecast Hub (“the Hub”) began collecting and creating ensemble forecasts (4), with the aim of facilitating systematic forecast evaluation and synthesis (5,6).

Producing reliable forecasts of public health-relevant quantities is challenging. Developing and testing models and methods can be particularly difficult during an emerging, rapidly changing outbreak—precisely when those models are most critically needed. Changes in policies, behavior, and reporting requirements can influence the data availability, reliability, and timeliness of data, which can lead to unreliable predictions. For example, changes in availability of diagnostic tests or changes to case definitions can influence data on case counts in ways that can be difficult to predict and which, if not accounted for, can bias forecasts.

Incorporating wastewater surveillance data into forecasting could help address some of these challenges(7–9). For many human pathogens, infected individuals shed fragments of pathogen genetic material into the sewage system. Wastewater from a community sampled prior to treatment at a wastewater treatment plant can detect the concentration of pathogen genetic material via polymerase chain reaction (PCR). Tracking concentrations of pathogen genetic material in wastewater over time is thus a potential way to monitor changes in infection prevalence. This approach has several potential strengths as a form of epidemiological surveillance: it is continuously-updated, passive, and cost-effective (10,11). It can potentially capture both symptomatic and asymptomatic infections, as well as infections in those who do not have access to healthcare or do not seek healthcare when sick (12). Wastewater surveillance systems can also be adapted quickly to monitor novel pathogens, as with the recent expansions of wastewater monitoring to include targets for the monkeypox virus, H5N1 influenza viruses, and respiratory syncytial virus (13–16). Some analyses suggest wastewater concentrations can be a leading indicator of infection incidence relative to traditional metrics such as incident hospital admissions or deaths (17–19), which lag infection incidence due to biological delays. We therefore hypothesized that including wastewater data might improve forecasts of incident hospital admissions, particularly at critical changepoints, such as the start of a surge or when infections are reaching the peak and beginning to decrease. Wastewater surveillance systems can also be adapted quickly to monitor novel pathogens, as with the recent expansions of wastewater monitoring to include targets for the monkeypox virus, H5N1 influenza viruses, and respiratory syncytial virus (13–16).

Data on pathogen concentration in wastewater can be challenging to integrate into infectious disease forecasting models, which typically use more traditional epidemiological data such as incident hospital admissions or case counts. In most cases, wastewater catchment areas do not correspond directly to healthcare reporting regions. To use wastewater to forecast health outcomes at the level of the healthcare reporting region, a model must integrate data from multiple wastewater treatment plants (sites). Additionally, wastewater sites can drop in and out from monitoring programs and have different laboratory processing methods, collection frequencies, reporting latencies, and degrees of population coverage. Thus, combining data from multiple sampling sites is not straightforward (20,21). While methods to fit wastewater data from multiple sites (22) and methods to combine wastewater data with other epidemiological signals (23) do exist, there is no overlap in these methods to enable inference across multiple sites and integrate traditional epidemiological signals. Moreover, while a number of methods to use wastewater data for epidemiological nowcasting, forecasting, and effective reproductive number estimation have recently been developed (19,24–30), most of these have not evaluated forecast performance of the method with and without wastewater data included. The most similar existing work to this study is Rademacher et al. (31), which evaluates several statistical and machine learning models for 7-day-ahead national hospital admissions forecasts in Germany, with one of these methods using an aggregated national wastewater signal as an input. Differences between our work and theirs include the models examined (semi-mechanistic renewal model versus statistical and machine learning models), the use of non-aggregated data from individual sampling sites versus aggregated data, the size and number of the locations forecast (52 subnational jurisdictions versus a national forecast), and the forecast horizon (28 days versus 7 days).

We developed a hierarchical Bayesian renewal model that infers unobserved infection dynamics from either incident hospital admissions alone (which we refer to as the “hospital admissions-only model”) or jointly with wastewater concentration data from multiple sites and hospital admissions (the “wastewater-informed model”) to produce forecasts of incident hospital admissions. We evaluated the models’ performance both compared to other forecasts submitted to the U.S. COVID-19 Forecast Hub (6) and in a head-to-head comparison with and without wastewater. The within-model comparison with and without wastewater addresses how much the wastewater signal affects the model’s predictions and thus its forecast performance. The comparison to other Hub forecasts puts this in context; within-model forecast performance differences are only meaningful if the model itself performs reasonably well compared to other available forecasting models, e.g. an improvement in forecast performance from including wastewater is not meaningful if both models are very poor-performers.

We submitted real-time wastewater-informed forecasts of COVID-19 hospital admissions from our model to the Hub (6) from February 5, 2024 to April 29, 2024, when the Hub paused accepting submissions. We produced forecasts for 52 jurisdictions (the 50 U.S. states, the District of Columbia, and Puerto Rico). We assessed performance of these real-time forecasts compared to forecasts from other models submitted in real-time to the Hub during that period. We also evaluated how forecasts from the hospital admissions-only version of the model would have performed, had we submitted it in real-time, using outputs saved from real-time runs (but not submitted to the Hub).

After the Hub paused submissions, we continued to develop our model and published it as an open-source R package, `wwinference` (32). To assess model performance over a longer time period, we used `wwinference` to produce hypothetical weekly forecasts for the 2023-24 season, using only data that would have been available as of the hypothetical forecast date. We refer to these as “retrospective forecasts” because they were produced after the forecast date had passed and based on a version of the model that had been updated in light of our real-time submission experience.

We evaluated retrospective forecasts from both the hospital admissions-only and wastewater-informed versions of our model. We compared the forecast performance of both the hospital admissions-only and wastewater-informed versions of our model to forecasts from other models submitted to the Hub throughout the season and assessed the value of wastewater for forecasting within our model by comparing forecast performance between our models with and without incorporating wastewater. Based on these analyses, we discuss areas for future development in wastewater-informed forecasting.

## Methods

Our model predicts hospital admissions and viral concentrations in wastewater based on inferred latent infection dynamics. The model accounts for the collection site and analyzing laboratory of each wastewater sample. We use a Bayesian approach to fit the model and generate forecasts from the posterior predictive distribution. Both in real-time and retrospectively, we fit the model to hospital admissions alone and jointly to incident hospital admissions and wastewater concentrations.

In this section, we describe the data sources, the generative model, the workflow to produce forecasts, and our evaluation metrics and forecast score comparisons.

### Data Sources

#### Hospital admissions data

For the duration of the study period, from October 16, 2023 through April 30, 2024, hospitals in the U.S. reported daily incident hospital admissions with COVID-19 to the U.S. Department of Health and Human Services (HHS) via the National Healthcare Safety Network (NHSN) (33) on a mandatory basis. These data were made available in a standardized format at the state, territorial, and national level each Friday, with daily resolution timeseries data for dates through the preceding Friday. We used data at the state or territorial level for the following jurisdictions: the 50 states, the District of Columbia, and Puerto Rico. We archived “vintaged” datasets (snapshots of the dataset as it appeared on a particular date), every Monday from October 16, 2023 to April 29, 2024. These vintaged datasets allow us to produce retrospective forecast submissions using only data that would have been available at the time for teams submitting to the Hub (5,6). The forecasts were evaluated against the dataset as it appeared on healthdata.gov on March 10^th^, 2025 (33).

On May 1, 2024, reporting of incident hospital admissions to HHS via NHSN transitioned from mandatory to voluntary (34). Our analysis covers only the mandatory reporting period.

#### Hospital admissions outlier exclusion

For real-time submissions to the Hub, we visually inspected the hospital admissions data as of each week’s data release. We excluded (i.e., treated as missing) data points that we judged to be outliers or appeared to be reporting artifacts (e.g. a repeated series of identical values for multiple days in a row). While we used outlier detection algorithms (35) to inform our decisions, the ultimate decision was by human judgment.

For the retrospective forecasts, we did not exclude any data points. For more details on the differences in production between the real-time and retrospective forecasts, see Table S1. Overview of the differences between real-time and retrospective analyses.

### Wastewater data

We used a dataset of SARS-CoV-2 RNA concentrations in wastewater maintained by CDC’s National Wastewater Surveillance System (NWSS) (36). The dataset is described on the NWSS website (36), and a dashboard (21) with summary statistics at the regional and national level available publicly. Data are available at data.cdc.gov; additional data may be available to researchers upon request to NWSS.

Wastewater sampling sites regularly report their wastewater data to NWSS, with weekly reporting recommended. Sites report estimated viral genome copies per unit volume of wastewater or per unit mass of dry sludge at the level of individual samples. Metadata includes not only the site where a sample was collected but also the lab where the sample was processed. Some sites use multiple labs or switch labs, and many labs serve more than one site; these details are accounted for in our hierarchical Bayesian modeling framework, as described below.

At the time of this analysis, the NWSS dataset was updated every day at 9 pm U.S. Eastern Standard Time. For real-time submissions, we used the data available as of Saturday after 9 pm EST to generate forecasts submitted to the COVID-19 Forecast Hub on Mondays.

To enable future retrospective forecasting and model testing, we created vintaged datasets from the complete NWSS data at regular intervals. We vintaged the NWSS dataset every Monday at 9 pm from October 16, 2023 to April 29, 2024. We used those vintaged datasets to produce our retrospective forecasts.

#### Wastewater data pre-processing

The NWSS dataset consists of samples collected at wastewater treatment plants. Our model takes as a primary input raw measurements of viral concentrations in wastewater, reported in genome copies per unit volume, from primary wastewater treatment plants.

Our model also uses as inputs data on sample collection dates, collection sites, processing labs, limits of detection, and population sizes of wastewater plant catchment areas. For full details on how these data are pre-processed and how outliers are handled, see S1. Text Additional pre-processing details.

### Generative Model

#### High-level overview

Figure 1 provides an overview of the generative model used to produce the retrospective forecasts of COVID-19 hospital admissions. We made changes to our model throughout the real-time submission period and continued to develop and change the model thereafter based on our performance evaluation of the 2023-2024 winter epidemic wave (October 16, 2023-March 25, 2024). We summarize model changes over time in S2. Text: Model changes over time.

**Figure. 1.**
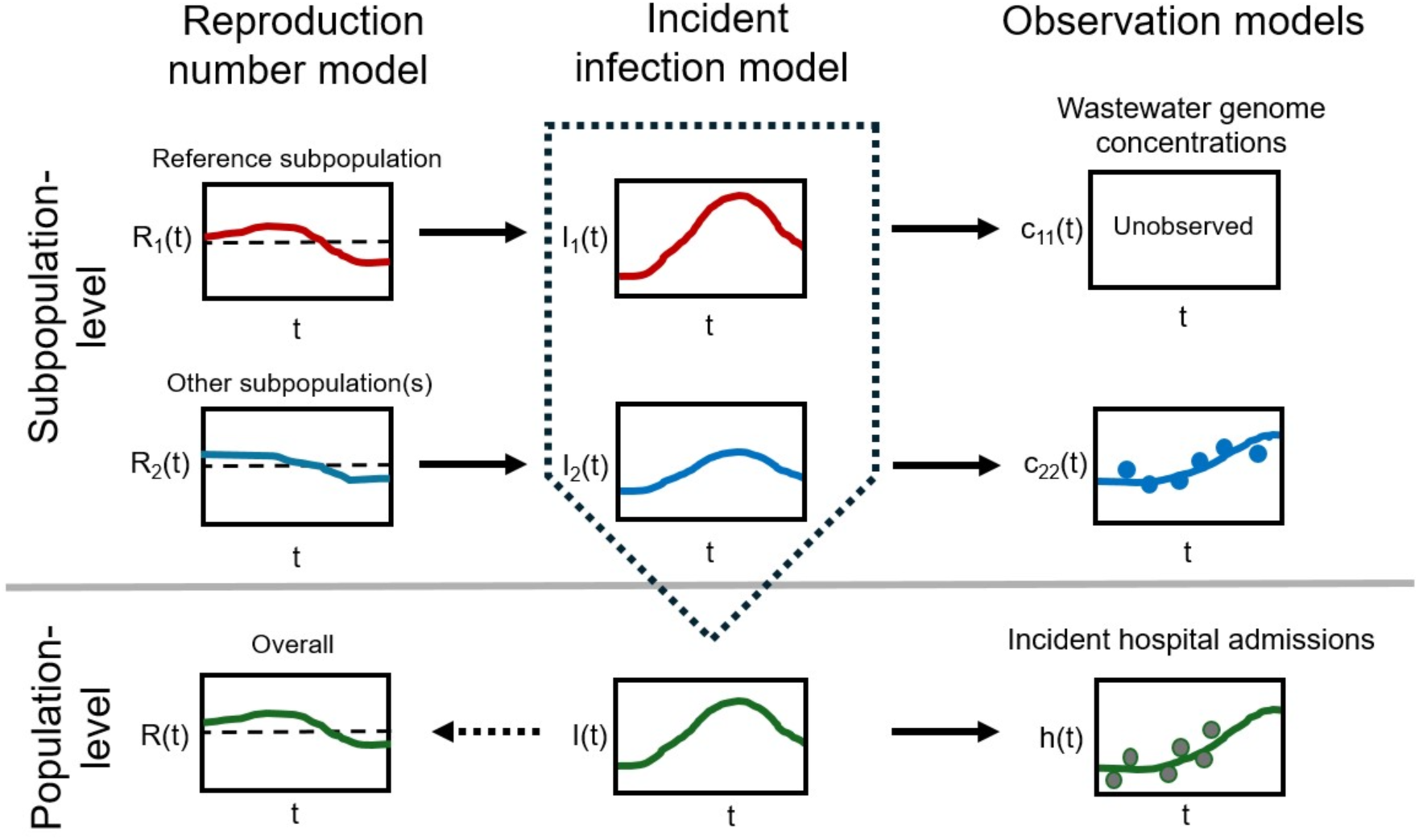
Schematic diagram of the wastewater-informed forecasting model. The ℛ_k_(𝑡) (k = 2, 3, …), the effective reproductive numbers for each subpopulation are modeled as deviations from a reference subpopulation (k = 1, typically the subpopulation representing all individuals who do not belong to any wastewater sampling site). Subpopulation-level incident infections are generated based on the ℛ_k_(𝑡) via a renewal equation. Total population-level incident infections are the sum of the subpopulation-level incident infections. An implied population-level reproduction number can be back-calculated from that incidence timeseries. Expected wastewater concentrations for each site are generated from the associated subpopulation’s incident infections via a wastewater shedding model. Expected population-level incident hospital admissions are generated from population-level incident infections via a model of the probability of admission given infection and the delay between infection and hospital admission.

Wastewater concentration data corresponds to one or more site-level subpopulations. In our model, we assume each subpopulation is a subset of the overall jurisdiction population, and that incident COVID-19 hospital admissions come from the jurisdiction’s entire population. For our wastewater-informed model, we divide the total population into 𝐾_total_ non-overlapping subpopulations: 𝐾_sites_subpopulations representing the people in each site’s wastewater catchment area, and one subpopulation representing all people in the jurisdiction not covered by the wastewater surveillance system.

Infections are generated via a semi-mechanistic renewal model approach (26,37–41). The variables of the renewal model for each subpopulation are:

- The effective reproductive number (Fig 1 left column).
- The incident infections (Fig 1 middle column).

We used a hierarchical model to model potential associations between the infection dynamics in each subpopulation (26,37–41). Each subpopulation *k*’s effective reproductive number at time *t* is estimated relative to the ℛ_k=0_(𝑡) in the subpopulation not covered by wastewater, which we refer to as the “reference subpopulation”. This structure results in subpopulation-level infection dynamics that are partially pooled across subpopulations, with deviations from the reference subpopulation occurring to a greater or lesser degree, depending on the data. Jurisdiction-level latent incident infections are the sum of the latent infections in the subpopulations. We fit the model independently for each jurisdiction (state or territory); there was no pooling of information across jurisdictions (e.g. California and Connecticut are fit separately, any correlations across state boundaries are therefore not taken into account). This approach to jointly fit wastewater and hospital admissions data differs from models that incorporate wastewater and other signals as covariates, as in this case each data stream is modeled from latent infections with an approach that represents our understanding of how each type of data is generated.

The hospital admissions-only version of the model is a special case of this hierarchical model with only one subpopulation: the reference (Fig 1, see S3. Text Model Definition: Subpopulation Definition).

The two types of observable data, wastewater concentrations at the subpopulation level and hospital admissions at the jurisdiction level (Fig 1C), are generated conditional on latent subpopulation-level and jurisdiction-level infections, respectively.

A full description of the model is provided in the S3. Text: Model Definition.

#### Implementation & forecast production

We implemented our model in the probabilistic programming language `Stan` (42) based on the semi-mechanistic renewal model in the `EpiNow2` R package (38,39), version 1.3.4. We used `cmdstan` version 2.35.0, `cmdstanr` (43) version 0.8.0, and `R` version 4.4.0 (44) to fit the model to data. We performed inference using a No-U-Turn Sampler (NUTS), a form of Markov chain Monte Carlo (MCMC) (45).

For each model run, we ran 4 NUTS chains for 750 warm-up iterations and 500 sampling iterations, with a target average acceptance probability of 95% and a maximum tree depth of 12.

Both in real-time and retrospectively, we generated forecasts every Monday (the Hub submission day). We calibrated the model to 90 days of observed hospital admissions data and, where relevant, all wastewater concentration data for that period available as of the forecast date. As of Monday, the most recent available daily hospital admissions counts were those for the Friday 10 days earlier (i.e. two Fridays previous). The lag between the last observed wastewater concentration data and the forecast date varied across time, location, and wastewater treatment plant.

Every Monday, we predicted 38 days of unobserved hospital admissions counts: 10 “nowcast” days from the last observed data day through the Monday the forecast was made, and 28 forecast days following. The 28-day forecast horizon was the Hub standard (6); the Hub did not accept nowcasts. We refer to the difference between the forecast date and the target date for a particular prediction as “horizon days”.

Figure 2 describes the workflow for forecast production and forecast evaluation.

**Figure 2.**
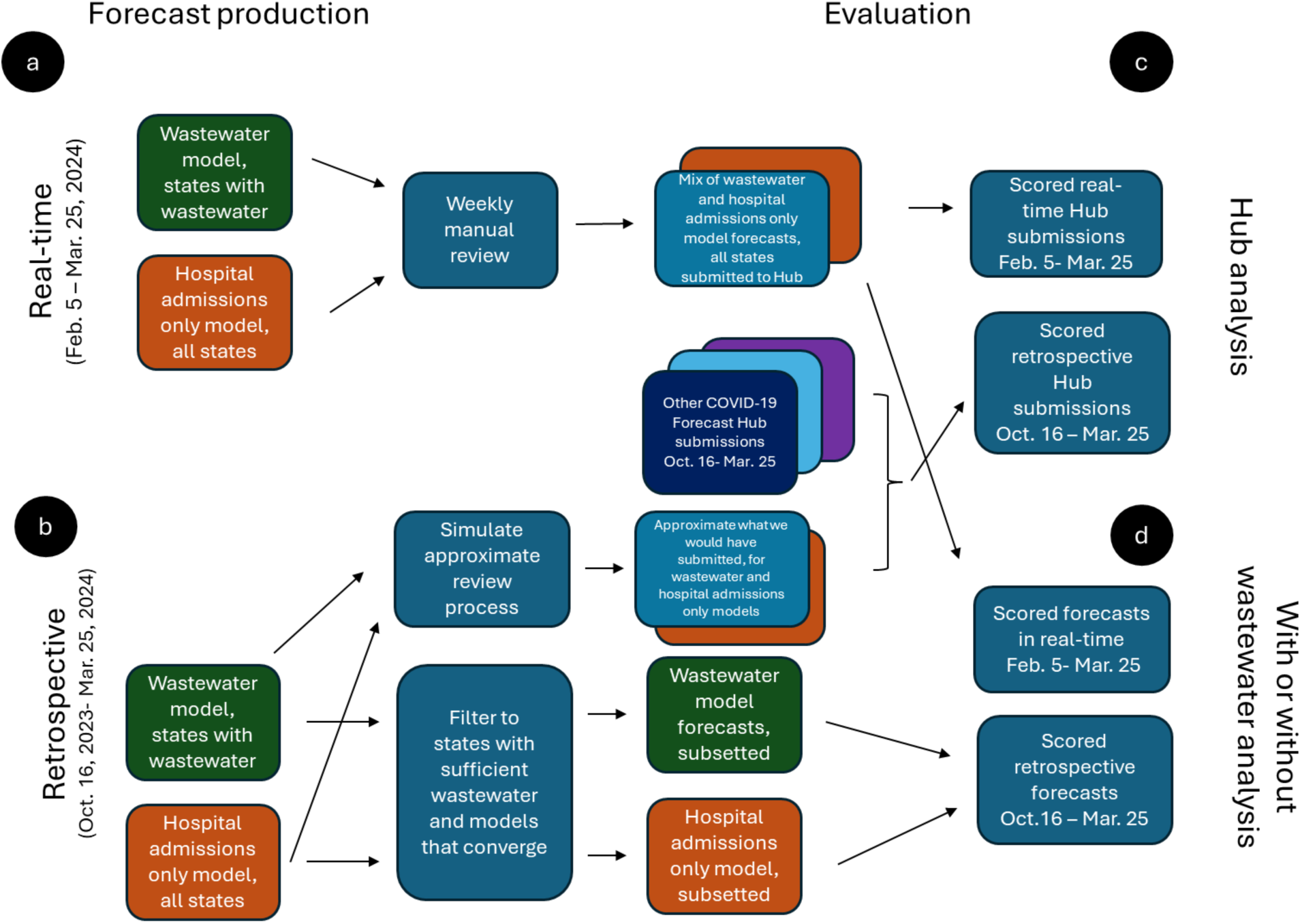
Workflow diagram for production of real-time and retrospective forecasts and comparison to other Hub models and with and without wastewater data. A. Workflow for the real-time forecast production of both models. B. Workflow for the production of retrospective forecasts for both models across the 2023-2024 winter epidemic wave. C. Workflow to compare forecast performance to models submitting to the Hub, in real-time and retrospectively. D. Workflow for comparing forecast performance with and without wastewater data, in real-time and retrospectively.

#### Real-time forecast production

We submitted forecasts to the Hub every Monday from February 5, 2024 to April 29, 2024. We generated forecasts on Saturdays for all 52 jurisdictions and then reviewed and submitted forecasts on Mondays.

We developed a software pipeline to generate forecasts using the R package `targets` (46), with a `crew` (47) backend to parallelize jobs. The pipeline was set up to compute the Hub’s requested quantiles (6) from the 2,000 posterior draws for each of the 28 days after the forecast date.

Each week, we recorded which jurisdictions either did not have wastewater data or had insufficient wastewater data to inform a forecast. We performed manual review of the outputs to decide which version of the model (wastewater-informed or hospital admissions-only) to submit for each jurisdiction (Fig. S1, see S4. Text Criteria for forecast inclusion for further details). We archived all hospital admissions-only forecasts locally.

We submitted forecasts to the COVID-19 Forecast Hub GitHub (6) as the model `cfa-wwrenewal`. We also provided the quantile forecasts on our own GitHub repository with additional metadata: a decision record of the model version used for each jurisdiction and metadata on the underlying data (48). See https://github.com/CDCgov/wastewater-informed-covid-forecasting/output/forecasts/2024-02-05/metadata.yaml for an example from February 5, 2024.

#### Retrospective forecast production

Using the model as implemented in `wwinference` `v0.1.1` (49), we produced retrospective forecasts for the 2023-2024 winter epidemic wave from both the wastewater-informed model and the hospital admissions-only model (Fig. 2B). The version of the model used in this analysis was developed in part using the data available and experience from the 2023-24 season, as we repeatedly made a change to the model, refit the model to the data, evaluated forecast performance out-of-sample, and used this information to inform model changes.

We used vintaged NHSN hospital admissions datasets and vintaged NWSS data to produce weekly forecasts for every Monday from October 13, 2023 through March 25, 2024 (24 weeks). Only wastewater-informed forecasts that met the criteria for sufficient wastewater and model convergence were included in the evaluation analysis (see S4. Text Criteria for forecast inclusion).

We fit and postprocessed individual forecast date-location pairs in parallel using Azure Batch (50). We combined and analyzed output from these model runs using `targets` (46).

Retrospective manual data exclusion for forecasting cannot be guaranteed to mimic a real-time process because the people curating the data retrospectively may be biased by their knowledge of what subsequently occurred. When creating retrospective forecasts, we therefore took a conservative approach and did not perform any manual exclusions of input hospital admissions data (as we had done in real-time).

##### Construction of retrospective Hub forecast submissions

We constructed retrospective Hub forecast submission files for all 52 jurisdictions for each week during the 2023-24 season for both the wastewater-informed model (`cfa-wwrenewal(retro)`) and the hospital admissions only model (`cfa-hosponlyrenewal(retro)`) (Fig. 2C). As in real-time, we computed the Hub requested quantiles (6) from the 2,000 posterior draws for the 28 days ahead of the forecast date.

For `cfa-wwrenewal(retro)`, we attempted to replicate as closely as possible our real-time submission process resulting in a forecast “submission” that contained a mix of wastewater-informed forecasts and forecasts from the hospital admissions-only model (Fig. S2, right panel). In real-time, we recorded all forecast date location-pairs for which we swapped the wastewater-informed model for the hospital admissions-only model on our GitHub (48)(Fig. S1). We used this record to ensure we used the same model in the creation of the retrospective version of the submission during that overlapping time period (Fig. S2, right panel). For the dates outside of the real-time submission period, we used the hospital admissions-only model instead of the wastewater-informed model whenever any of our objective criteria for forecast inclusion were not met (e.g. convergence, availability of sufficient wastewater, see S4. Text: Criteria for forecast inclusion for further details). See Fig. S3 for a record of these substitutions and the reasons for them by forecast date and location.

### Evaluation

#### Scoring

We evaluated forecasts against the most recent available NHSN hospital admissions dataset from the mandatory reporting period (data through April 27^th^ 2024; retrieved on March 10^th^, 2025). We evaluated all forecast dates for which we had evaluation data for the full 28-day forecast period. The last evaluated forecast date was therefore March 25^th^, 2024.

For all forecast scoring, we transformed both the observed hospital admissions data and the posterior predictions by the function 𝑓(𝑥) = ln (𝑥 + 1); the offset of 1 handles zero counts, since ln (0) is undefined. Previous work has found that scoring predictions of log-scaled disease incidence is more robust to the absolute magnitude of the quantity being forecasted and reduces biases that favor underprediction near the peak when the process being forecast is exponential (51). We computed forecast scores and other evaluation metrics using the R package `scoringutils` (`v2.1.2`) (52,53).

We evaluated forecasts quantitatively using the continuous ranked probability score (CRPS), weighted interval score (WIS), quantile coverage, interval coverage, and bias. CRPS is a sample-based proper scoring rule that is a generalization of the absolute error, used to evaluate probabilistic forecasts. We used CRPS for the retrospective analysis of model performance with and without wastewater, since all posterior draws were available. WIS is a quantile-based proper scoring rule that is an approximation the CRPS for quantile-based forecasts (54). We used WIS to evaluate forecasts submitted to the Hub as these were submitted in a quantile format. Quantile and interval coverage assess the calibration of a forecast (e.g. what percent of data fall within the predicted quantile or interval and how does this deviate from what is expected?). We calculated quantile coverage for all Hub quantiles (6) and interval coverage for the 30, 60 and 90 percent prediction intervals stratified by horizon. Bias assesses a forecast’s tendency to overpredict (positive bias) or underpredict (negative bias)(53). We computed bias for all forecasts, both real-time and retrospective.

In all instances where we visualize or report a relative score (rCRPS or rWIS), this is a simple ratio: the average absolute score for each model summarized across the dimensions of interest divided by the corresponding average score for the baseline model (this is in contrast to some forecasting analyses where rCRPS or rWIS corresponds to a WIS or CRPS based “scaled relative skill”, see for instance (53)). In the text, we report the fold-change improvement or reduction in performance, as well as the relative score and ratio of absolute scores. When visually summarizing the distribution of relative scores, we use the geometric mean and the 68% and 95% intervals.

#### Comparison to the COVID-19 Forecast Hub

To evaluate the forecast performance of the model compared to other models submitting forecasts to the Hub, we analyzed the real-time submissions we submitted to the Hub from February 5, 2024 through March 25, 2024 (`cfa-wwrenewal(real-time)`), the real-time hospital admissions only forecasts we produced but did not submit to the Hub (`cfa-hosponlyrenewal(real-time*)`), and retrospective forecasts generated using `wwinference` fit to the historical vintaged datasets from the full 2023-24 season for both the wastewater-informed model (`cfa-wwrenewal(retro)`) and hospital admissions-only model (`cfa-hosponlyrenewal(retro)`). See Table S2 for a description of each of the wastewater-informed and hospital admissions only renewal models analyzed in this study.

We compared our real-time and retrospective forecasts to forecasts from models submitted to the Hub that met the criteria described in S5. Text Further details on forecast evaluation: Selection of Hub submissions for comparison. Eight individually submitted models, meaning models that were submitted by individual teams rather than created by the Hub either from the data alone or from other submitting models, met these criteria: `CEPH-Rtrend_covid`, `CMU-TimeSeries`, `MUNI-ARIMA`, `SGroup-RandomForest`, `UMass-gbq`, `UMass-sarix`, `UMass-trends_ensemble`, and `UT-Osiris`. See Table S2 for a description of each of the Hub models. When comparing the rank of our model we always compare to these eight models plus the hospital admissions-only and the wastewater-informed model, for a total of 10 models. We also compared our performance against the standard Hub ensemble model created from individually submitted models (`COVIDhub-4_week_ensemble`) and the Hub baseline model (`COVIDhub-baseline)`. Using the `COVIDhub-4_week_ensemble` as a comparator, we computed relative WIS (rWIS) values for each individual model; rWIS is the ratio of the target model’s average score to that of the comparator model for all and only the cases in which both models made a forecast (53). We used the full set of 12 models to compute the standardized rank of the models at each forecast date and location (defined as 1 minus the quantile rank, so 1 indicates first place and 0 last place by WIS) for each forecast-date location pair. To simplify main text visualizations, we included only the Hub ensemble (`COVIDhub_4_week_ensemble`) and the best-performing individual models by overall WIS for the full October through March evaluation period (`CMU-TimeSeries` and `UMass-sarix`) alongside our own models in most figures.

### Forecast comparison with and without wastewater

#### Real-time February-March 2024

For the real-time wastewater-informed forecast submissions, we pulled wastewater-informed forecasts directly from the Hub GitHub and scored them alongside other model submissions. To compare real-time predictions from the wastewater-informed and hospital admissions-only models, we used our archived real-time output from the hospital admissions-only model, labeling it as `cfa-hosponly(real-time*)`, with an asterisk to indicate that, while produced in real-time, it was not submitted to the Hub. In this analysis, we only scored the 28 day-ahead quantile-based forecasts for consistency with forecasts submitted to the Hub in real-time. We only evaluated forecasts for which the wastewater-informed model was submitted, and wastewater data was both present and sufficient to inform the model. We define “sufficient wastewater” in S3. Text: Criteria for forecast inclusion.

#### Retrospective across the 2023-2024 winter epidemic wave

For the retrospective analysis, we produced daily probabilistic sample-based nowcasts and forecasts using both the wastewater-informed and hospital admissions only models for the entire 2023-2024 winter epidemic wave for all forecast dates and locations that met the criteria for sufficient wastewater and model convergence (S4. Text: Criteria for forecast inclusion). We scored both the 10-day nowcasts and the 28-day ahead sample-based forecasts using the CRPS.

To understand the distribution of relative performance with and without wastewater, we computed a relative mean CRPS or WIS (rCRPS or rWIS) for the two models for each individual forecast. Specifically, we computed the arithmetic mean of the CRPS or WIS score for each model at each forecast date and jurisdiction, across horizon days. We then computed relative scores with the hospital admissions-only model as the baseline. For example, when stratifying by location in the retrospective analysis across the 2023-24 epidemic season, we expect to have at most 24 relative scores in each location, corresponding to the relative score of the wastewater-informed model to the admissions-only model at each of the 24 forecast dates analyzed.

#### Exploratory investigation of drivers of prediction and forecast performance heterogeneity

In an exploratory analysis, we analyzed potential drivers of reduced and improved performance of our retrospective wastewater-informed model compared to our hospital admissions-only model across the 2023-2024 winter epidemic wave. Using a hierarchical linear regression on input data for each forecast, we inferred log-linear trends in wastewater data (inferred hierarchically across lab-sites) and jurisdiction-wide admissions data and compared these recent trends to the observed differences in forecasts and performance between the models. See S6 Text: Exploratory investigation of drivers of prediction and forecast performance heterogeneity for the full details of the regression model.

We analyzed visually the relationship between regression coefficients and model-estimated standard deviations as proxies for trends and variability in trends in wastewater and hospital admissions data. We visualized the following:

- The variability in recent trends across wastewater sites (as quantified by a hierarchical standard deviation in the slopes of the log-linear trends) and the relative performance of the wastewater-informed model compared to the hospital admissions-only model to investigate whether a high correlation between wastewater sites impacted relative forecast performance
- The relative forecast performance compared to the trends in the hospital admissions and wastewater data (as quantified by the global slope in the wastewater trend and the slope of the hospital admissions trend) to investigate whether the relationship between the trends in both datasets impacted forecast performance

We also investigated particular instances of poor forecast performance when we observed that wastewater data exhibited trends that did not coincide with hospital admissions trends via a visual analysis of the trend in flow population normalized wastewater concentration values.

Based on these exploratory visual analyses, we developed hypotheses for the drivers of forecast performance heterogeneity.

## Results

### Visual comparison of forecasts from three example jurisdictions on January 15th, 2024

We highlight examples of retrospective `wwinference` forecasts made from both the wastewater-informed model (green) and hospital admissions-only model (orange) for three jurisdictions on January 15, 2024 (Fig. 3). We selected these jurisdictions as illustrative examples of how wastewater data can improve, minimally affect, or worsen forecast performance.

**Figure 3.**
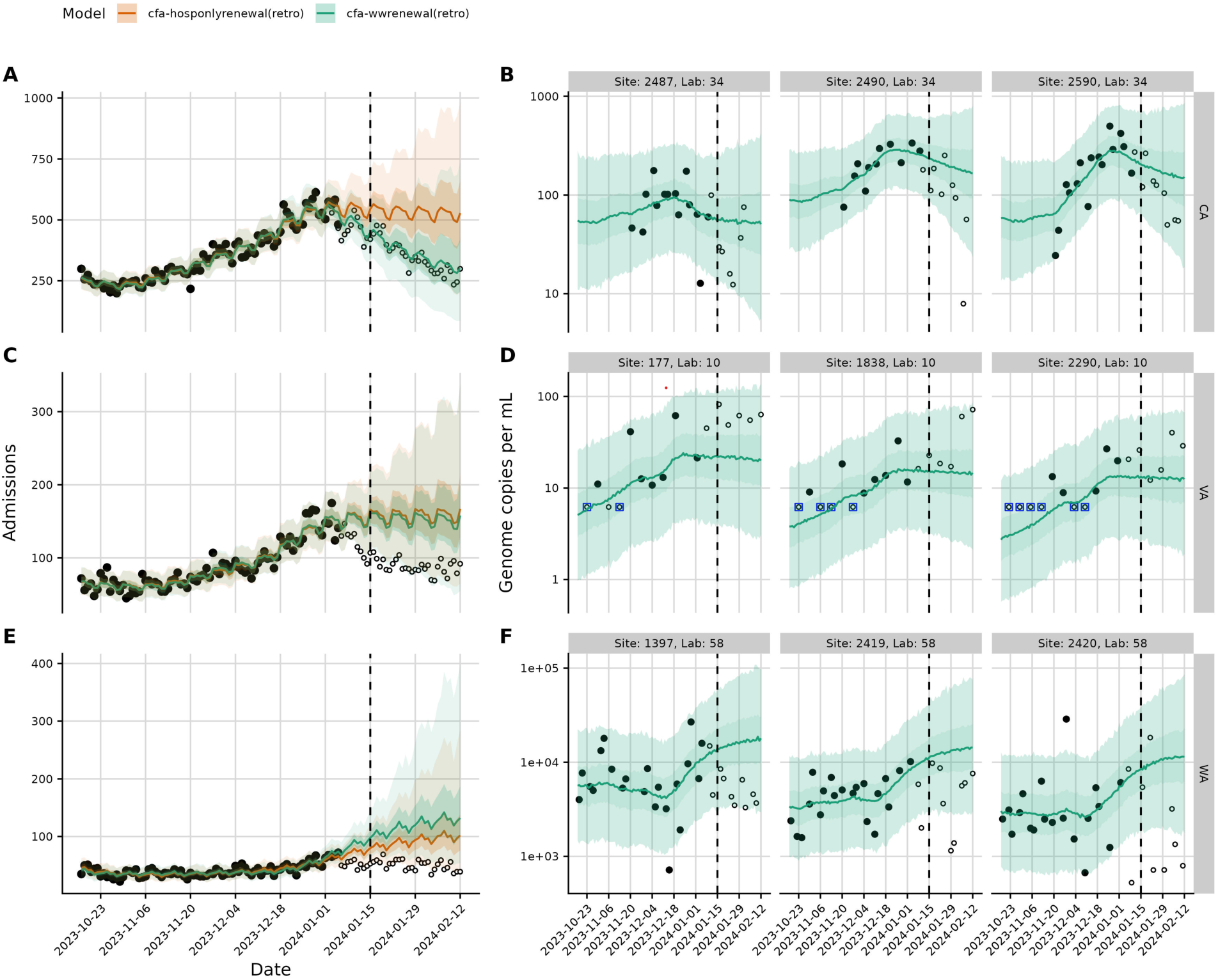
Example retrospective fits and forecasts of hospital admissions and wastewater concentrations from three jurisdictions on January 15, 2024. Closed circles indicate data available as of the forecast date, open circles indicate data that became available retrospectively, which is used to evaluate the forecast performance of each of the models. Blue squares indicate wastewater concentrations below the limit of detection. Rows correspond to California, Virginia, and Washington respectively (top to bottom) A. C. E.) Calibrated and forecasted hospital admissions from the wastewater-informed model (green) and the hospital admissions only model (orange) for a single forecast. B.D.F). Fit and forecasted wastewater concentrations from the wastewater-informed model for three individual sites and labs in each jurisdiction. Site and lab numbers are arbitrary and based on unique identifiers assigned to the anonymized wastewater concentration data.

Figure 3A (California) shows an example of the wastewater-informed model improving performance compared to the hospital admissions-only model. Wastewater concentrations show a downward trend in multiple site-lab pairs that is not yet evident in the hospital admissions trend (Fig. 3B). The wastewater-informed model therefore correctly projects that hospital admissions will trend down; the hospital admissions-only model does not anticipate the downturn.

Figure 3C (Virginia) shows an example of the wastewater-informed model performing similarly to the hospital admissions-only model. In this case, the wastewater data appear to have provided little additional information on the direction and certainty of the infection trend (Fig. 3D), and the two models’ predictions are very similar.

Figure 3E shows an example of the wastewater-informed model performing worse than the hospital admissions-only model. Both models overpredict future hospital admissions, but the overprediction is more extreme for the wastewater-informed model. Visual inspection of the wastewater concentration data (Fig. 3F) shows an increase in average concentration prior to the forecast date. This earlier increase likely led the wastewater-informed model to project a more extreme upward trend in admissions than the hospital admissions-only model, and this trend was ultimately not realized in the hospital admissions.

The performance of these three example jurisdictions across the 2023-2024 winter epidemic wave is assessed visually and via CRPS for both models in S7. Text Additional results: Visual and quantitative evaluation of forecast performance across three example jurisdictions (Fig. S4).

### Real-time forecast performance compared to other models submitted to the Hub

We evaluated both what the wastewater-informed model that was submitted to the Hub in real-time and the hospital admissions-only model forecasts we generated and saved locally in real-time compared to other models submitted to the Hub (Fig. 4).

**Figure 4.**
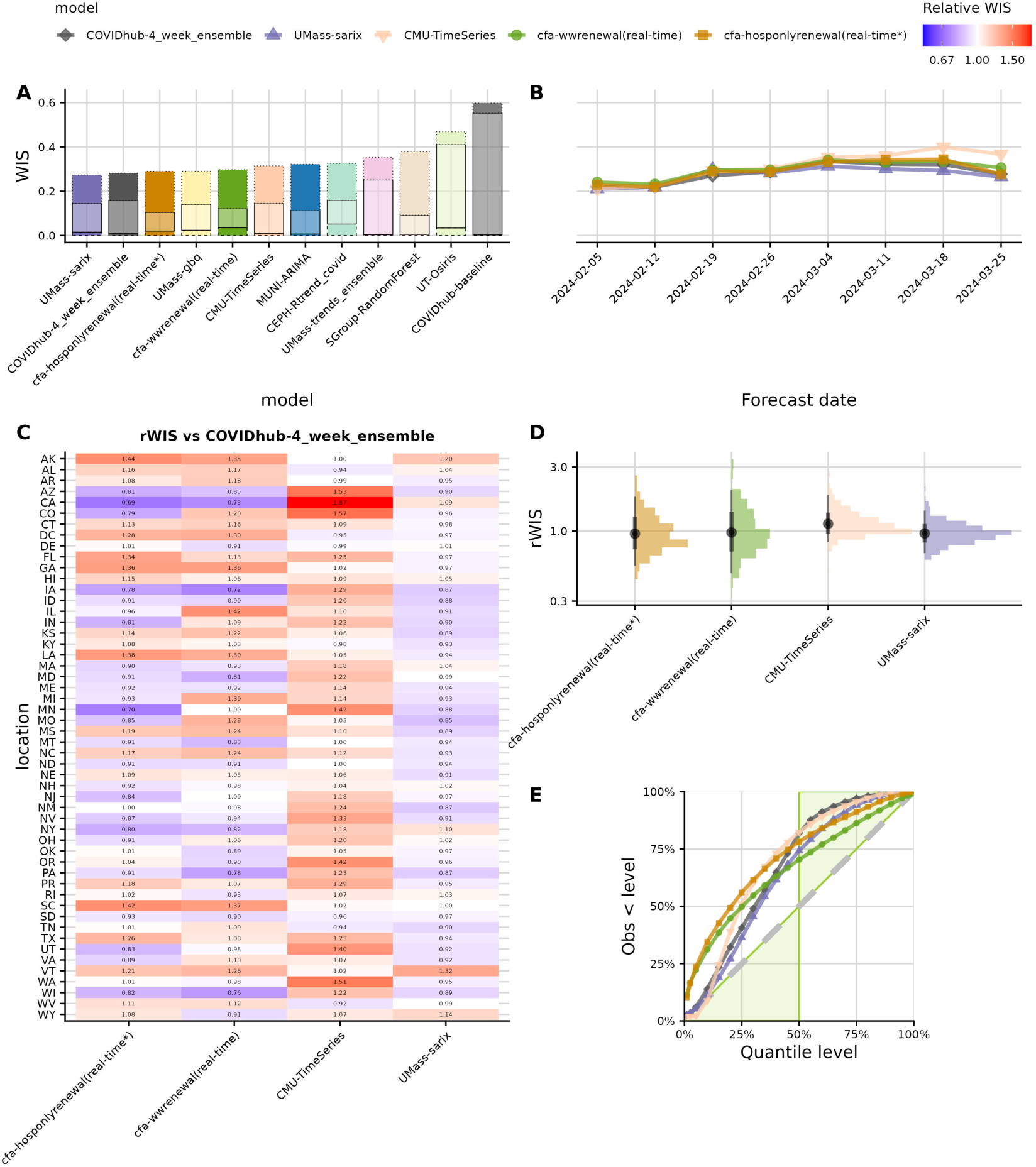
**Real-time forecast performance compared to other models submitted to the Hub** from forecast dates ranging from February 5^th^, 2024 to March 25^th^, 2024. Colors in panels A, B, D, and E correspond to a particular model and are consistent across panels, with the wastewater-informed model (`cfa-wwrenewal(real-time)`) in green and the hospital admissions only model (`cfa-hosponlyrenewal(real-time*)`) in orange. A. Overall average WIS by model, ordered from lowest (best) to highest (worst) WIS and decomposed into penalties for underprediction (lower solid bar), dispersion (middle transparent bar), and overprediction (top solid bar) averaged across all jurisdictions and forecast dates. This plot contains 12 bars because it includes the Hub baseline and Hub ensemble (in grays) in addition to 10 individually-submitted models (in non-gray colors). B. Average WIS across jurisdictions and horizon days at each forecast date, colored by model. C. Relative WIS (rWIS) by jurisdiction for each model, compared to the Hub ensemble. rWIS is computed as the ratio of the mean WIS across all predictions shared between the target model and the comparator model (here, the Hub ensemble) overlap. Blue indicates improved performance by rWIS; red indicates reduced performance. D. Distribution of rWIS versus Hub ensemble for individual forecast date-location pairs, by model. Points indicate the geometric mean, thick bars indicate the central 68% interval, thin bars indicate the central 95% interval E. Empirical quantile coverage colored by model. A well-calibrated forecast should have empirical quantiles that match the theoretical ones (gray dashed diagonal line). Green shaded area corresponds to conservative forecasts.

The model output we submitted to the Hub in real-time had the 4^th^ best score by WIS of 10 individually-submitted models that met analysis criteria (Fig. 4A). The 10 models include the hospital admissions-only model, which we did not actually submit in real-time, but which we include in the ranking for full context. Scored submissions are a mix of wastewater-informed and admissions-only forecasts, as determined by model convergence, wastewater data availability, and manual review (Fig. S1, Fig. S3). Had we submitted forecasts from the hospital admissions-only model for all jurisdictions (whenever that model converged and passed review), those hypothetical submissions would have ranked second of 10 (Fig. 4A).

The real-time wastewater-informed model performed 1.05 times worse by WIS than the Hub ensemble (rWIS = 1.05 [0.297/0.281] Fig. 4A, Fig. S6), averaged across all forecast dates and locations, whereas the hospital admissions-only model would have performed 1.03 times worse than the Hub ensemble (rWIS = 1.03 [0.290/0.281] Fig. 4A). The only individually-submitted model to outperform the Hub ensemble by overall average WIS during this period was the `UMass-sarix` model, which performed 1.03 times better (rWIS = 0.97 [0.273/0.281]). For individual forecasts (forecast date-location pairs), performance ranged from 3.16x better to 3.43x worse than the Hub ensemble for the wastewater-informed model and 2.30x better to 2.60x times worse for the hospital admissions-only model (Fig. 4D).

Across the 8 forecast dates for which real-time forecasts were submitted and scored, the wastewater-informed model was never the top-performing individual model by WIS for a given forecast date (Fig. 4B). The model tended to perform better when top performing individual models (e.g. `CMU-TimeSeries` and `UMass-sarix,` the two top performers by WIS across the season) also performed better and worse when they likewise performed worse (Fig. 4B). Performance by forecast date ranged from 1.02x worse to 1.10x worse by WIS than the Hub ensemble. Had we submitted it, the hospital admissions-only model would have ranged in its performance from 1.01x better to 1.07x worse than the Hub ensemble.

Our real-time submissions also showed heterogeneous performance by jurisdiction. The wastewater-informed model’s performance ranged from 1.39x better to 1.42x worse by WIS than the Hub ensemble and performed better than the Hub ensemble in 24 of 52 jurisdictions (Fig. 4C, D). The hospital admissions-only model would have ranged from 1.44x better to 1.44x worse and performed better than the ensemble in 26 of 52 jurisdictions.

Quantile coverage analysis showed that the wastewater-informed and hospital admissions-only models, `CMU-TimeSeries,UMass-sarix`, and the Hub ensemble were all biased upward at most or all predictive quantiles for the period analyzed (Fig. 4E). The real-time wastewater-informed model was less well-calibrated (more biased upward) than these comparator models at predictive quantiles lower than the 45^th^ percentile, but better calibrated (less biased upward) at higher predictive quantiles (Fig 4E). The wastewater-informed model was less biased upward than the hospital admissions-only model at all quantiles assessed (Fig. 4E, Fig S8).

Both the real-time wastewater-informed and hospital admissions-only had a bimodal distribution of standardized ranks for individual forecast date-location pairs (Fig. S5). Both were often below-median performers (25th percentile ranking of 0.36, 0.46 for the wastewater-informed model and hospital admissions-only model, respectively), but both were also often the top-performing model (75th percentile ranking of 0.91, 0.91) This bimodal performance contrasts with that of the Hub ensemble, which had a unimodal distribution of standardized rank, consistently in the top third of models but rarely best overall (25th percentile ranking of 0.64, 75th percentile ranking of 0.73).

### Evaluation of forecast performance across 2023-24 season

#### Retrospective forecast performance compared to other models submitted to the Hub across the 2023-24 epidemic season

For the retrospective comparison to the Hub submissions, we produced forecasts for all 52 jurisdictions across all 24 forecast dates from October 13, 2023, to March 25, 2024, for both models, resulting in 2,288 potential forecasts for evaluation. The wastewater-informed retrospective submissions contained a mix of wastewater-informed and hospital admissions-only model generated forecasts, as wastewater-informed models that failed to converge or had insufficient wastewater data were replaced with the corresponding forecast from the hospital admissions-only model (see Fig. S3), mirroring the Hub submission practice we followed in real-time (Fig. S1). The most important difference from our real-time submission process was that we performed no manual review for data outliers or forecast plausibility.

Had both the wastewater-informed and hospital admissions-only models been submitted across the 2023-24 epidemic season, they would have had nearly identical average WIS: WIS = 0.271 for the retrospective wastewater-informed model and 0.269 for the retrospective hospital admissions-only model. They would have ranked 5th and 4th, respectively, out of 10 qualifying individual models submitted to the Hub (Fig. 5A). Both would have outperformed the Hub baseline and performed less well than the Hub ensemble. The retrospective wastewater-informed and retrospective hospital admissions-only model would both have performed 1.06 times worse by overall WIS than the Hub ensemble (WIS = 0.255). For individual forecast problems (forecast date-location pairs), the wastewater-informed model performed from 4.26x better to 5.64x worse than the Hub ensemble across individual forecast dates and locations, whereas the relative performance of the hospital admissions-only model ranged from 2.43x better to 4.39x worse (Fig. 5D).

**Figure 5.**
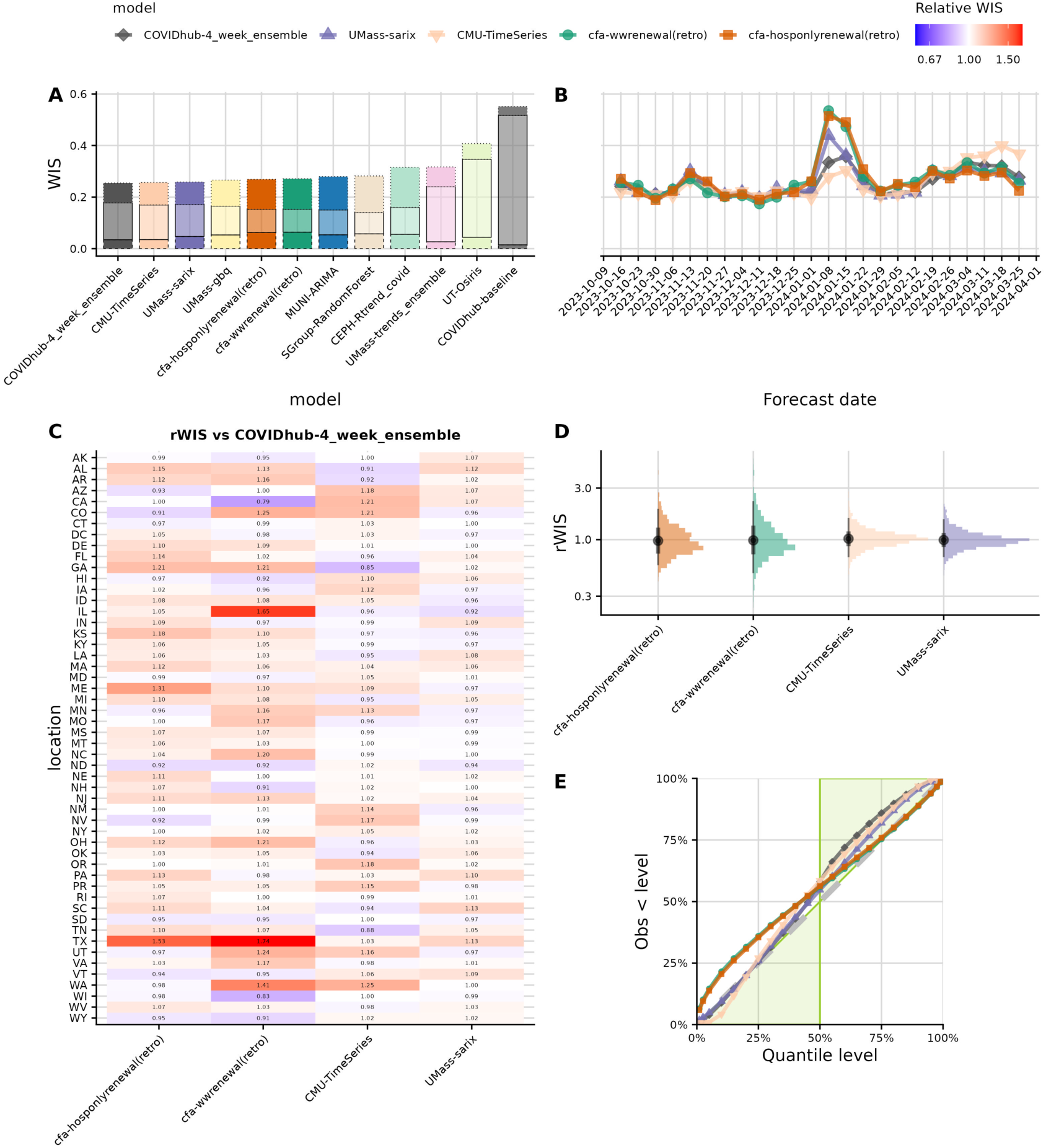
Retrospective performance compared to other models submitted to the COVID-19 Forecast Hub across the 2023-24 epidemic season. Colors in panels A, B, D, and E correspond to a particular model and are consistent across panels, with the wastewater-informed model (`cfa-wwrenewal(retro)`) in green and the hospital admissions only model (`cfa-hosponlyrenewal(retro)`) in orange. A. Overall average WIS by model, ordered from lowest (best) to highest (worst) WIS and decomposed into penalties for underprediction (lower solid bar), dispersion (middle transparent bar), and overprediction (top solid bar). Plot contains 12 bars because it includes the Hub baseline and Hub ensemble (in grays) in addition to 10 individually-submitted models (in non-gray colors). B. Average WIS across jurisdictions and horizon days at each forecast date, colored by model. C. Relative WIS (rWIS) by jurisdiction for each model, compared to the Hub ensemble. rWIS is computed by averaging the model’s WIS for the jurisdiction across all forecast dates and then computing the ratio of that mean WIS to the Hub ensemble mean WIS. Blue indicates improved performance by rWIS; red indicates reduced performance. D. Distribution of rWIS versus Hub ensemble for individual forecast date-location pairs, by model. Points indicate the geometric mean, thick bars indicate the central 68% interval, thin bars indicate the central 95% interval E.) Empirical quantile coverage colored by model. A well-calibrated forecast should have empirical quantiles that match the theoretical ones (gray dashed diagonal line). Green shaded area corresponds to conservative forecasts.

Both the wastewater-informed and hospital admissions-only model had their worst absolute performance for the 2024-01-08 and 2024-01-15 forecast dates (Fig. 5B), shortly after the peak in incident admissions (Fig. 6A). Most other Hub models also struggled on those forecast dates; in consequence, so did the Hub ensemble (Fig. 5B, S6, S7, S8). The `CMU-TimeSeries` model was the best performing model for those dates. The wastewater-informed model’s performance by forecast date ranged from 1.15x better to 1.61x worse by WIS than the Hub ensemble and outperformed the ensemble on 12 of 24 forecast dates. The hospital admissions-only model’s performance ranged from 1.23x better to 1.55x worse than the ensemble, also outperforming the ensemble on 12 of 24 forecast dates. Ten of these forecast dates of outperforming the ensemble were shared between our two retrospective models.

**Figure 6.**
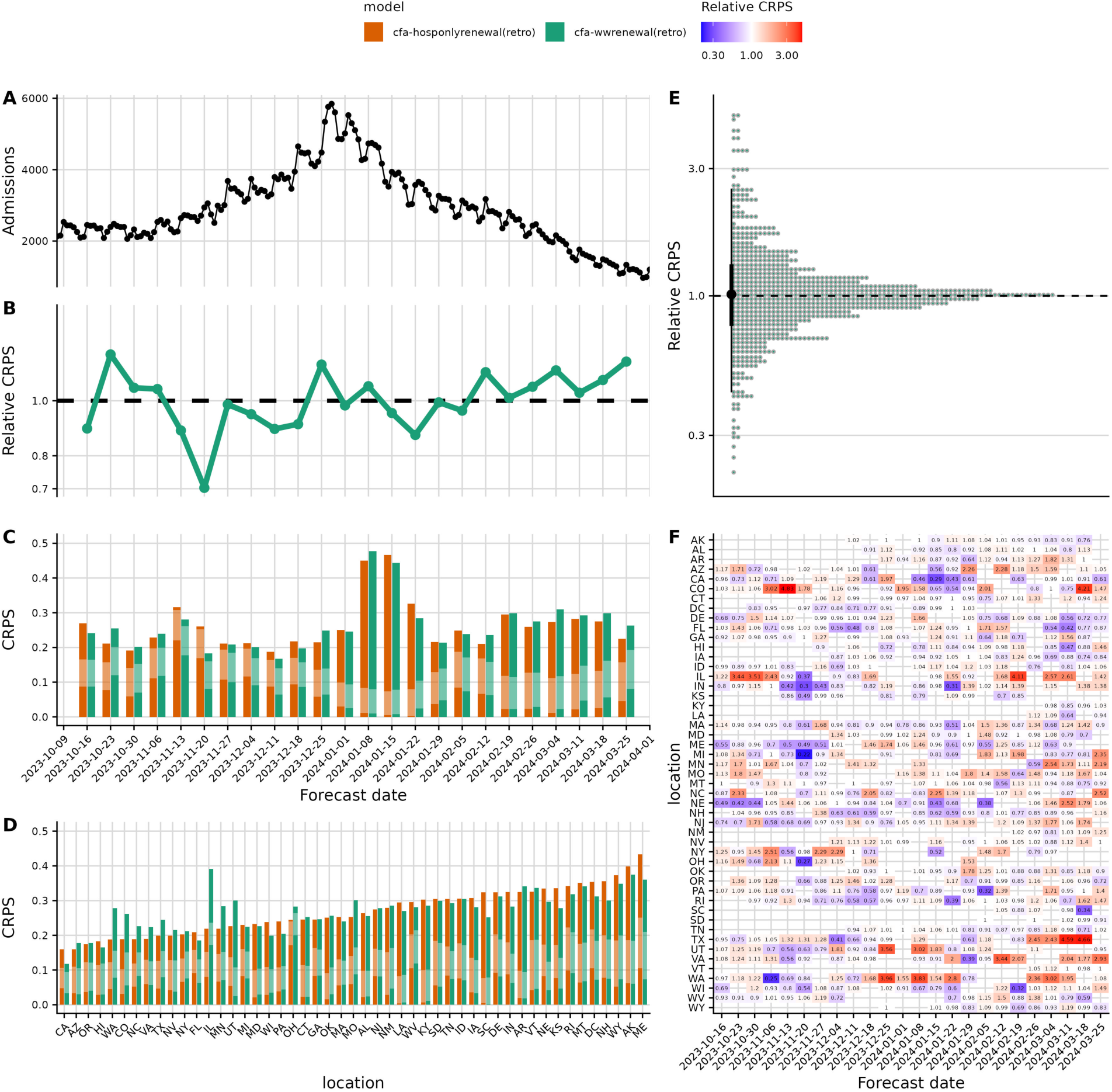
Retrospective forecast performance comparison with and without wastewater data from forecast dates ranging from October 16^th^, 2023 to March 25^th^, 2024. Colors in panels A, B, C, and D correspond to the wastewater-informed model (`cfa-wwrenewal(retro)`) in green and the hospital admissions only model (`cfa-hosponlyrenewal(retro)`) in orange. Relative continuous ranked probability score (rCRPS) computed by first averaging across the 38 horizon days and then computing the ratio of wastewater-informed mean CRPS to the hospital admissions-only model mean CRPS. For rCRPS, a value less than 1 indicates that the wastewater-informed model outperforms the hospital admissions-only model. A. Timeseries of observed U.S. national daily hospital admissions during the period evaluated. B. rCRPS by forecast date. C, D. Absolute CRPS by forecast date (C) and jurisdiction (D) for each model, decomposed into penalties for underprediction (bottom solid bar), dispersion (middle transparent bar), and dispersion (top solid bar). E. Distribution of rCRPS values for individual forecast date-jurisdiction pairs. F. Heatmap of rCRPS across forecast dates and jurisdictions. Blue indicates improved performance by rCRPS; red indicates reduced performance. Gaps indicate forecasts that were excluded from the analysis because the wastewater-informed model did not make a forecast due to absent or insufficient wastewater data, convergence issues, or the exclusion of the corresponding forecast in real-time.

The wastewater-informed model’s performance was heterogeneous across jurisdictions. Performance by jurisdiction ranged from 1.26x better to 1.74x worse by WIS than the Hub ensemble (Fig. 5C), and this model outperformed the ensemble in 17 of 52 jurisdictions. The hospital admissions-only model was slightly less variable; performance ranged from 1.10x better to 1.53x worse than the ensemble, and the model outperformed the ensemble in 19 of 52 jurisdictions.

The retrospective wastewater-informed and hospital-admissions only model overpredicted admissions at predictive quantiles below the 50^th^ percentile; other individual Hub models and the ensemble were better calibrated (Fig. 5E). At higher quantiles, the wastewater-informed and hospital admissions-only models were better calibrated than the other Hub models (Fig. 5E), while other models were more likely to overpredict admissions. All models tended to be biased upwards in January and again in late February and March (Fig. S20).

As was the case for the real-time models (Fig, S5), the distribution of forecast date-location standardized ranks was bimodal for both the wastewater-informed and hospital admissions-only retrospective models (Fig. S9), with both models often ranking low or high for a given forecast date and location. The Hub ensemble was consistently an above-median-performing model.

### Retrospective forecast performance with and without wastewater across in all jurisdictions across the 2023-24 epidemic season

The retrospective analysis spans 52 jurisdictions and 24 forecast dates from October 16, 2023 to March 25, 2024 (the 2023-24 season), resulting in 1,248 possible forecast date-location pairs. Of these, 42 were excluded because we had excluded one or both models from Hub submission in real time, an additional 140 had convergence flags for one of the two models, 109 had no wastewater data available, and 183 had insufficient wastewater data available. These exclusions leave 774 paired forecasts for head-to-head comparison of our two models (Fig. S2, S3).

The wastewater-informed model performed nearly identically to the hospital admissions-only model in the aggregate by continuous ranked probability score (CRPS): relative CRPS (rCRPS) = 1.01 (0.266 for the wastewater-informed model / 0.265 for the hospital admissions-only model], Fig. 6E & B). The distribution of log relative CRPS for individual forecasts (forecast date-location pairs) was approximately symmetrical (Fig. 6E, Fig. S10). The wastewater-informed model’s performance ranged 4.59x better to 4.83x worse by CRPS than the hospital admissions-only model (Fig. 6E & F).

Results were similar when we compared the real-time version of the subset of paired forecasts for which the wastewater-informed model was submitted, with an average overall WIS of 0.282 for the wastewater-informed real-time model and 0.281 for the hospital admissions-only real-time model (rWIS = 1.00, Fig. S11 A&B). See S7. Text: Additional results: Real-time forecast performance with and without wastewater (Fig. S11, S12, S13, S14, S15, S16) for more details on this analysis.

The two models performed similarly for most of the 24 forecast dates; the wastewater-informed model tended to perform better in late fall and worse in late winter and early spring (Fig. 6B, C. Fig. S17). Overall, the wastewater informed model performed better on 12 forecast dates and worse on the other 12, with no systematic relationship between the difficulty of the forecast problem (as measured by the CRPS of the hospital admissions-only model, Fig. 6B, C) and the relative performance of the wastewater model (Fig. S18). Performance of the wastewater-informed model by forecast date ranged from 1.42x better to 1.21x worse by CRPS than the hospital admissions-only model (Fig. 6B). Both models had their worst scores by absolute CRPS for the forecast dates 2024-01-01 and 2024-01-15, shortly after the peak in incident admissions (Fig. 5C). In real-time, there was less variability in relative forecast performance across forecast dates, with the wastewater-informed model performance ranging from 1.08x better to 1.10x worse than the hospital-admissions only model (Fig. S11B,C).

Stratifying by jurisdiction, there was substantial heterogeneity in relative forecast performance (Fig. 6D). The wastewater-informed model performed better than the hospital admissions-only model in 28 jurisdictions and worse in 21 jurisdictions (Fig. S19). Performance by jurisdiction ranged from 1.35x better to 1.79x worse by CRPS than the hospital admissions-only model.

Both models were biased downwards in November and December when admissions were increasing (Fig. S20), biased upwards in early January when admissions peaked, and again biased upwards in late February and early March, when hospital admissions further declined (Fig. 6A). The wastewater-informed model was slightly less biased (Fig. S10). Overall, both models were similarly biased upward on average (average bias = 0.052 for the wastewater-informed model and 0.051 for the hospital admissions-only model, Fig. S21). Both models were overconfident (Fig. S22); empirical interval coverage was slightly worse for the wastewater-informed model than for the hospital admissions-only model across horizons and intervals.

#### Investigation of forecast prediction and performance heterogeneity

The high degree of heterogeneity in relative forecast performance between the wastewater-informed and hospital admissions-only model (Fig. 6F) led us to investigate potential drivers of differences in forecast performance.

We did not see a systematic pattern in relative versus absolute performance (Fig. S10, S18, S19); indicating the wastewater-informed model did not perform systematically better or worse when the forecasting problem was more or less difficult.

We visualized the relationship between the correlation in wastewater sites and relative forecast performance (Fig. S23) observing that the worst rCRPS values tended to cluster in regions which had consistent trends in wastewater concentrations across sites (low slope standard deviation) (Fig. S23, red points in the upper left).

We also visualized the agreement between trends in both wastewater and hospital admissions versus relative forecast performance (Fig. S24 & S25), observing that the worst rCRPS (highest) values tended to cluster around small differences between the trend in wastewater and hospital admissions, whereas the best rCRPS (lowest) values tended to have a larger difference between trends in the wastewater and hospital admissions, (Fig. S25, red points at the top of the graph and blue points at the bottom).

Particular instances of poor forecast performance were observed in the wastewater concentrations in many sites in Illinois and Ohio in February and March (Fig. S26 & S27), when concentrations declined rapidly then subsequently rebounded. This trend was consistent across wastewater treatment plants, but no such “divot” was observed in the hospital admissions. The retrospective wastewater-informed model performed poorly in Illinois and Ohio during that period (Fig. 6F). In real-time, we were skeptical of realism of these wastewater-informed model results and submitted forecasts from the hospital admissions-only model to the Hub for Ohio for all dates in February and March, and for Illinois on February 26 (Fig. S1 & S3). A visual comparison of flow population normalized wastewater concentrations versus the unnormalized concentrations used in the model showed them to be relatively similar and thus likely to have similarly poor performing forecasts (Fig. S28 & S29).

## Discussion

### Summary of key findings

We developed a hierarchical Bayesian generative renewal model that integrates hospital admissions and optionally wastewater data to produce forecasts of COVID-19 hospital admissions. We used the model to submit wastewater-informed forecasts to the United States COVID-19 Forecast Hub (the Hub) in real-time from February to April of 2024 and published a model implementation as an open-source R package, `wwinference`, which allows the user to apply the model to other locations and geographic granularities, for example at the sub-state level. We found that, compared to other models submitted to the Hub during the period we submitted, both our wastewater-informed model and the hospital admission-only model performed similarly, ranking 4^th^ and 2^nd^ respectively out of 10 individually submitted models, with the hospital admissions-only model scoring slightly better by overall WIS. When we retrospectively produced forecasts for the entire 2023-2024 winter epidemic wave, the findings were similar: either model would have been a median-performer overall had it been submitted to the Hub for the entire season. In a head-to-head analysis of all and only the forecasts where we could have submitted either a wastewater-informed or a hospital admissions-only model, the models showed comparable overall performance. Beneath this similar overall forecast performance, there was significant heterogeneity across locations and time periods, both in real-time and retrospectively.

### Insights and hypotheses from exploratory analysis

Our interpretation of the exploratory analysis of performance heterogeneity is that the wastewater-informed model tended to perform worse relative to the hospital admissions-only model when wastewater sites were highly correlated and when the trends between the wastewater and hospital admissions signals were similar. Based on this, we hypothesize that the model may exhibit overconfidence when multiple input data streams point in the same direction. This is consistent with our finding that the wastewater-informed model undercovered across prediction intervals compared to the hospital admissions-only model. In our model, individual wastewater sites are treated as independent random samples from the jurisdictional catchment areas, despite the fact that they are likely to be correlated with one another due to clustering of sampling in areas with wastewater surveillance programs. Trends in wastewater concentrations from wastewater sites may be more likely to be correlated with the jurisdictional hospital admissions signal if wastewater sites are preferentially located in densely populated cities compared to more rural areas (55). We therefore see adding more realistic correlation structures, for example based on geographic locations or human mobility networks, to our wastewater observation model as a potential area for future model development.

Additionally, we also observed instances when misaligned signals between wastewater and hospital admissions data appear to have misled the model into confidently projecting that wastewater trends would be reflected in future admissions (rather than, say, increasing the uncertainty appropriately to reflect the conflict between the present trends). In Ohio and Illinois in February and March 2024, we hypothesize that the decline and subsequent rebound in wastewater concentrations may have been driven by heavy rainfall (56), as rain can cause dilution of wastewater concentrations, particularly in communities with combined sewer systems (57).

### Relationship to previous literature

As a result of the expansion of wastewater surveillance during the COVID-19 pandemic, several methods have been developed to leverage wastewater concentration data to produce nowcasts, forecasts, and effective reproductive number estimates (19,23–31). To our knowledge, none of the existing real-time surveillance models developed have integrated raw wastewater observations from multiple sites with epidemiological count data like hospital admissions or cases, and few evaluations examine forecast performance for a single model with versus without wastewater data. An exception is (31), which evaluated including wastewater data as an exogenous predictor in a machine learning model to produce 7-day ahead national hospital admissions forecasts in Germany. The authors found similar results to those presented here: including wastewater did not improve their otherwise-best-performing forecast model. Similarly, recent work evaluating the predictive performance of wastewater for nowcasting retrospectively found that adding wastewater had minimal impact above that provided by case data (58).

### Limitations of this study

We note several limitations of the model presented here. First, our wastewater-informed model does not always successfully produce a forecast. In the retrospective analysis across the 2023-2024 winter epidemic wave, we found that the wastewater-informed model failed to converge more frequently than the hospital admissions-only model. Until the wastewater-informed version of the model can be made more reliably convergent, a back-up scheme, like the one we had which used the hospital admissions-only models as a back-up is likely needed for real-time forecasting.

Second, while our model allows for noise and bias in site-level wastewater measurements, in our model any consistent trends in wastewater concentrations can only arise from corresponding trends in subpopulation infection incidence. In reality, a number of other factors likely contribute to trends in wastewater concentrations, such as the example described above regarding the impact of rainfall on observations, changes in sample collection or lab processing methods, shedding from animal waste or animal byproducts, or changing shedding rates in different SARS-CoV-2 variants. In the same vein, hospitalization data also has factors that are poorly understood and not captured in the current model, for example, shifts in the infection to hospital admissions rate that may occur due to capacity constraints during surges or due to changes in disease severity from different variants.

Third, our model assumes that the populations represented by wastewater data are a representative and random sample of the jurisdictional population. While around 80 percent of U.S. households are served by municipal wastewater collection systems (36), the populations covered by wastewater surveillance may underrepresent some communities (e.g. rural areas, people in diapers) which might have different infection dynamics (59,60). The populations covered might also be correlated with one another, for example if there are multiple wastewater sites within a city. The model is fit independently to each jurisdiction and therefore doesn’t account for correlations between jurisdictions which are likely to be present.

Fourth, our model treats populations within a wastewater catchment area as homogenous, with no age, risk, or contact structure imposed, and does not explicitly model the age structure in the jurisdiction-level population that gives rise to the reported hospital admissions. We know that the hospital admissions are more representative of infections from older and higher-risk individuals and thus are not always reflective of transmission levels if infections are not evenly distributed throughout the population (61,62).

Fifth, we used a mix of weakly informative and informative priors based on external literature, and due to computational costs, a full prior sensitivity analysis was not performed.

Finally, the current implementation of the model does not account for inter-individual variability in shedding rates. This may be of particular importance in smaller wastewater catchment areas, where it is likely that a small number of infected individuals are contributing to the overall measured wastewater pathogen concentrations.

There are also several key limitations of our evaluation approach. As mentioned above, we developed the version of the model tested in the retrospective analysis using data from the 2023-2024 winter epidemic wave, as we did not have previous vintages of historical wastewater data, and subsequently performed a retrospective evaluation of the model using this same dataset. In an ideal world, we would have iterated on our model using a subset of the data and evaluated it against an entirely held-out set of testing data. Additionally, our evaluation period spanned a limited time period of less than a year for the retrospective analysis and eight weeks for the real-time analysis. We describe additional model and analysis limitations in S8 Text: Additional limitations.

### Unanswered questions and future research

Given our findings, we see several key areas for future work in wastewater-informed forecasting, both in terms of development of this model specifically and more broadly in the field of wastewater-based epidemiology. First, we saw that our model reacted to trends in wastewater dynamics that did not appear to reflect changes in infections and thus admissions but instead are a result of the characteristics of the sewer system and its interaction with the environment. For example, a combined drainage and sewage system would be more affected by stormwater from rainfall than separate systems. Sewersheds can also have external inputs such as industrial input and animal waste/byproducts (63,64). Another factor to consider is whether the population served is transient or whether large gatherings occurred around the sampling events, all of which could impact the signal of pathogen concentration. Models that explicitly account for these extrinsic factors and the characteristics of the sewer system could perform better, though we note that simple commonly performed normalizations (such as flow-population normalization) did not suffice to correct these “divots”.

Second, our analysis into the drivers of forecast and performance heterogeneity has led us to hypothesize that our wastewater-informed model is overconfident and over-weights a high correlation within wastewater sites and between wastewater concentrations and hospital admissions, which can lead to extreme, over-confident forecasts that result in over- or under-prediction. This phenomenon would be more likely to occur in jurisdictions with many highly correlated sites (e.g. those from the same or nearby cities), explaining why we saw more variability in relative performance with and without wastewater across jurisdictions than we did across forecast dates. Addressing this issue with the model structure may improve performance. Additional investigation into characteristics of jurisdictions and their wastewater surveillance system (e.g. characteristics like population coverage, number of sites, latency from sample collection to reporting, etc.) and its relation to relative forecast performance should also be investigated further.

Third, poor performance of both of our models during times of peak hospital admissions may be due to the renewal model structure and model for the time evolution of the effective reproductive number, as these are shared between both variations of the model. This observation is of particular importance for future model improvement because important healthcare capacity decisions are made during peak transmission times (1–3), and focusing efforts to improve peak performance (65) could improve overall performance for both models.

Fourth, based on literature indicating that wastewater data could be a leading indicator for hospital admissions (17–19), we initially hypothesized that including the wastewater would improve the model’s ability to detect trend changes in hospital admissions. While we saw examples where this appeared to be the case, our finding that overall, incorporating wastewater did not improve forecast performance, indicates that it was not broadly true. Delays between wastewater sample collection and reporting, which we made sure to account for here by using vintaged wastewater datasets may explain this finding, but the intrinsic delays from infection to shedding and from shedding to arrival in a wastewater treatment plant could also be contributing factors. A recent analysis of hypothetical zero-reporting-lag wastewater concentration curves found that they neither led nor lagged several incident case metrics (66), and incident cases can be a leading indicator for more severe outcomes (67). Future analyses could assess whether reducing lags from sample collection to reporting meaningfully improve wastewater-informed epidemiological forecasts, both in this model and other model types.

While our findings are conditional on one dataset and one model, the field of wastewater-based epidemiology is likely to benefit from assessing the value of wastewater for epidemiological forecasting across a range of data sources, surveillance settings, and modeling methodologies. Studies that assess the impact of different characteristics of the wastewater surveillance system on predictive accuracy, such as lag, sample collection frequency, and other factors, could help inform resource prioritization to improve the utility of wastewater surveillance systems. Additional utility may come from development and evaluation of models that account for the wastewater collection, sampling, quantification, concentration, and molecular detection processes as well as how the number of contributing infected individuals impacts biological and measurement variability. Identifying factors that contribute to improved and reduced forecast performance with wastewater data, with the goal of optimizing both model inputs and resource allocation for wastewater surveillance, may eventually lead to both more efficient data collection and better performing forecasts.

As seen in this study, quantitative evaluations of forecast performance can facilitate evidence-based model development and support investigation of key scientific questions. Improvements in the composability of models to develop new features—such as accounting for spatial correlation structure or fitting jointly to multiple clinical datasets— may help maximize the utility of available data and improve our understanding of the value of additional data. Wastewater-based epidemiology, in particular, may benefit from these approaches because wastewater-surveillance systems differ greatly across geographies and pathogens, requiring the ability to flexibly modify models to cater to a specific surveillance system and pathogen setting. Finally, access to vintaged (time-stamped) data is essential for principled testing of nowcasting and forecasting models (68,69), as we need to know exactly how the data would have appeared to a forecaster on a given day. For this study we manually created wastewater data snapshots—a resource-intensive process that presents a significant barrier to researchers. Broader access to vintaged datasets, via efforts like the CMU Delphi data archive and associated `epidatr` R package (68) could help researchers develop and test models.

### Conclusion

While our results are limited by the single epidemic season, set of locations, model choices, and data surveillance system studied, they point to the promise and the challenge of incorporating novel epidemiological signals into forecasting models. Our model’s overall average forecast performance was very similar when the wastewater signal was included versus when it was excluded, but that overall similarity masked heterogeneity. There were times when including wastewater meaningfully improved model performance and times when including it meaningfully detracted from performance, as well as instances in which it had minimal effect. With the aim of improving future versions of our model, we investigated potential drivers of reduced and improved relative forecast performance and identified several areas of further exploration. This study highlights the role that forecast evaluation can play in developing and testing novel epidemiological models and data streams.

## Data Availability

The wwinference R package v0.1.1 implementing our model, which we used for retrospective forecasts, is available at: https://github.com/CDCgov/ww-inference-model/releases/tag/v0.1.1.
All code used to make the manuscript figures are available in our pipeline repository's GitHub at: https://github.com/CDCgov/wastewater-informed-covid-forecasting. In the README.md, we describe how the evaluation pipeline code depends on the wwinference R package, and how one would run the full pipeline to reproduce the results. The code used to produce the real-time forecasts (via older versions of the model) is no longer maintained, though it can be examined via the git commit history of the pipeline repo. For this reason, reproducing the real-time results using the current evaluation pipeline is not easily done.
The real-time forecasts as submitted are available in the pipeline repository's output/forecasts folder as well as on the COVID-19 Forecast Hub's GitHub (https://github.com/reichlab/covid19-forecast-hub/tree/master/data-processed/cfa-wwrenewal/).
The NWSS full analytics dataset is available to researchers on request to NWSS: https://www.cdc.gov/nwss/about-data.html. All hospital admissions data used in this analysis is publicly available from https://healthdata.gov/Hospital/COVID-19-Reported-Patient-Impact-and-Hospital-Capa/sgxm-t72h/about_data. Vintages of the admissions are available via the R package epidatr (formerly covidcast).

https://github.com/CDCgov/ww-inference-model/releases/tag/v0.1.1

https://github.com/CDCgov/wastewater-informed-covid-forecasting

https://github.com/reichlab/covid19-forecast-hub/tree/master/data-processed/cfa-wwrenewal/

https://www.cdc.gov/nwss/about-data.html

https://healthdata.gov/Hospital/COVID-19-Reported-Patient-Impact-and-Hospital-Capa/sgxm-t72h/about_data

## Acknowledgments

We would like to thank the following individuals for their feedback and contributions to this work and to various related projects: Hannah Cohen, John Glasser, Adam Howes, Sasha Keyel, Chirag Kumar, Adrian Lison, Agastya Mondal, Eric Mooring, Nimesh Patel, Cristin Young and Zoe Yu. We would specifically like to thank the members of CDC’s National Wastewater Surveillance program (NWSS) for collaborating with us on this work: Dan Cornforth and Heather Reese, as well as all wastewater surveillance programs submitting their data to NWSS, and all facilities submitting hospital admission data to the National Healthcare Safety Network (NHSN).

We acknowledge the financial support from CDC Grant NU38FT0008 (K.E.J & S.A.) and CDC Grant NU23FT0069 (G.V.Y).

## Data & Code Availability

The wwinference R package v0.1.1 implementing our model, which we used for retrospective forecasts, is available at: https://github.com/CDCgov/ww-inference-model/releases/tag/v0.1.1.

All code used to make the manuscript figures are available in our pipeline repository’s GitHub at: https://github.com/CDCgov/wastewater-informed-covid-forecasting (48). In the README.md, we describe how the evaluation pipeline code depends on the `wwinference` R package, and how one would run the full pipeline to reproduce the results. The code used to produce the real-time forecasts (via older versions of the model) is no longer maintained, though it can be examined via the git commit history of the pipeline repo. For this reason, reproducing the real-time results using the current evaluation pipeline is not easily done.

The real-time forecasts as submitted are available in the pipeline repository’s output/forecasts folder as well as on the COVID-19 Forecast Hub’s GitHub (https://github.com/reichlab/covid19-forecast-hub/tree/master/data-processed/cfa-wwrenewal/).

The NWSS full analytics dataset is available to researchers on request to NWSS: https://www.cdc.gov/nwss/about-data.html. All hospital admissions data used in this analysis is publicly available from https://healthdata.gov/Hospital/COVID-19-Reported-Patient-Impact-and-Hospital-Capa/sgxm-t72h/about_data. Vintages of the admissions are available via the R package `epidatr` (68) (formerly `covidcast` (70)).

## Notes

### Competing Interest Statement

The authors have declared no competing interest.

### Funding Statement

We acknowledge the financial support from CDC Grant NU38FT00008 (K.E.J., S.A.) and CDC Grant NU23FT0069 (G.V.Y.).

