## Supplement for "Bayesian generative modeling for heterogeneous wastewater data applied to COVID-19 forecasting"

### S1. Text: Additional pre-processing details

NWSS requires that only one observation is reported per day in a particular wastewater treatment plant and lab.

We pre-processed the full wastewater dataset for use in our model by filtering for:

- Inclusion of only primary wastewater treatment plants.
- Inclusion of only SARS-CoV-2 amplification targets.
- Exclusion of samples flagged for quality issues.
- Exclusion of samples from solids.

Including only primary wastewater treatment plants ensures that overlapping catchment areas are not included in the data.

The following variables from the pre-processed dataset were used as data in the model:

- The reported Polymerase Chain Reaction (PCR) concentration (in genome copies per unit volume).
- The units of the reported concentration.
- The date the sample was collected.
- The limit of detection associated with the sample.
- The unique identifier for the wastewater treatment plant where the sample was collected.
- The population size associated with the wastewater catchment area where the sample was collected.

- The unique identifier for the lab where the sample was processed.
- The jurisdiction of the wastewater treatment plant.

All reported concentrations and limits of detection were converted to genome copies per milliliter. An indicator variable identifying whether a measurement was above the limit of detection was assigned to each observation. A unique identifier was assigned to each site and lab pair. We did not aggregate the data across sites or labs within a jurisdiction as this is subject to biases due to differences in sites' reporting latency and drop-out. Instead, we group the observed concentrations by unique site-lab combinations and fit directly to these observations (see S1. Text Model Definition: Wastewater concentration observation model).

#### *Wastewater data outlier exclusion*

We identify outlier viral genome concentrations for each unique site and lab pair with an approach based on $z$-scores.

Briefly, we compute $z$-scores for the concentrations and their finite differences and remove any observations above specified threshold values for either metric. For each set of wastewater concentration observations from an individual site-lab pair:

- We only attempt to detect outliers in wastewater observations above the limit of detection (LOD). Sub-LOD observations are still included in the data, modeled as a censored process (See S1. Text Model Definition: Censoring wastewater observations below the limit of detection).
- Exclude observations more than 90 days before the forecast date.
- Compute the change per unit time between successive observations $t$ and $t'$: $(\log[c_{t'}] - \log[c_t])/(t' - t)$.
- Compute $z$-scores for $\log[c_t]$ across all sites $i$ and timepoints $t$. Flag values with $z$-scores over 3 as outliers and remove them from model calibration.
- Compute $z$-scores for the change per unit time values across all sites and pairs of timepoints. For values with $z$-scores over 2, flag the corresponding wastewater concentrations $c_t$ as outliers and remove them from model calibration.

The $z$-score thresholds were chosen by retrospective visual inspection of the data from July - October 2023. All flagged outliers were excluded from the model inputs (i.e. treated as missing).

### S2. Text: Model changes over time

We made changes to the model both during and after the period when we submitted real-time forecasts to the COVID-19 Forecast Hub. While some of these changes were made without explicit reference to data to evaluate the change, other changes came from fitting the model to subsets of our vintaged datasets, producing forecasts, and evaluating the change in forecast performance with the new model development. This process ideally would have been done with training and test datasets that were entirely separate from the vintaged data we used to produce retrospective forecasts for evaluation (e.g. data from the prior epidemic season), so that the evaluation represented a fully out-of-sample prediction exercise. However, we did not have time-stamped datasets as of each forecast date for the wastewater data to facilitate this. For this reason, all retrospective results should be interpreted with caution, as they were partially tuned to the subsamples of data from the same season being forecasted.

Below, we provide a high-level overview of major model changes. All of these major changes occurred after the first 6 weeks of submitting the real-time model; that is, they are the key differences between the real-time models and final model used for the retrospective runs. We did also make model changes during the real-time submission period. We refer the reader to the GitHub history in our model definition (Johnson et al., 2024) in the pipeline repository for the model changes made during the real-time submission period and to the GitHub history in our model definition in the `wwinference` package for the full set of changes made after the submission period.

- Modified the way that the jurisdiction-level infections are generated such that instead of generating incident infections from the central $\mathcal{R}(t)$ estimate, subpopulation-level incident infections are generated from subpopulation $\mathcal{R}(t)$ estimates, and then subpopulation-level incident infections are summed to get the total jurisdiction level infections
- Changes to priors in the auto-correlation coefficient in the time-evolution of the central $\mathcal{R}(t)$ and the auto-correlation coefficient in the deviation of the subpopulation $\mathcal{R}(t)$ from the central $\mathcal{R}(t)$. These previously had the same prior.
- Bug fix to the bounds on the IHR to force it to be strictly between 0 and 1.
- Fixed a bug whereby draws from a standard normal distribution were incorrectly forced to be positive.
- Major change to the structure of how the subpopulation $\mathcal{R}(t)$s are estimated. Previously, each subpopulation, including the subpopulation covered by wastewater, was estimated as a deviation from a latent central $\mathcal{R}(t)$. As of the restructure, each of the subpopulations with wastewater observations are estimated as deviations from a reference subpopulation plus a static offset. This changes the meaning of the deviations, as this enforces that the deviation in the central $\mathcal{R}(t)$ and the deviation in the reference subpopulation are not the same magnitude or estimated in the same way as the wastewater subpopulation deviations.
- Major change to the prior on the infection feedback term and the prior on the average step size in the $\mathcal{R}(t)$ weekly random walk. Previously, we had used a high and strong prior on infection feedback that reduced $\mathcal{R}(t)$ significantly at periods of high incidence. We have modified this to be more moderated, at the same time

increasing the prior on the step size in the $\mathcal{R}(t)$ weekly random walk to allow for more deviation that is independent of the infection feedback.

### S3. Text: Model definition

#### A. Effective reproductive number $\mathcal{R}(t)$

##### *Unadjusted effective reproductive number $\mathcal{R}_0^{\mathrm{u}}(t)$*

We model incident infections and the effective reproductive number $\mathcal{R}(t)$ as latent variables. We decompose $\mathcal{R}(t)$ into two components: an unadjusted instantaneous reproduction number in the reference subpopulation $\mathcal{R}_0^{\mathrm{u}}(t)$ and a damping term that accounts for the effect of recent infections on the instantaneous reproduction number (Asher 2018).

We model the unadjusted reproduction number in the reference subpopulation $\mathcal{R}_0^{\mathrm{u}}(t)$ as piecewise-constant with weekly change points. We chose weekly change points as we did not expect more rapid fluctuations in the inferred infection trends to be relevant and to improve computational efficiency. The time evolution of log $[\mathcal{R}_0^{\mathrm{u}}(t)]$ is modelled as a once-differenced auto-regressive (AR) process, inspired by the once-differenced implementation option in `EpiNow2` (1), and the AR(1) process implementation in ( *et al.*, 2020)

$$\log[\mathcal{R}_0^{\mathrm{u}}(t)] = \log[\mathcal{R}_0^{\mathrm{u}}(t-1)] + \beta(\log[\mathcal{R}_0^{\mathrm{u}}(t-1)] - \log[\mathcal{R}_0^{\mathrm{u}}(t-2)]) + \mathrm{Normal}(0, \sigma_r)$$

where $t$, $t-1$, and $t-2$ are days in three successive weeks, $\beta$ is an autoregression coefficient which serves to make week-to-week changes correlated and $\sigma_r$ determines the overall variation in week-to-week changes and can be thought of as the typical magnitude of log-scale weekly change. A once-differenced AR process was chosen to impose some degree of correlation in the week-to-week trend in $\mathcal{R}(t)$. Our choice of the differenced AR process on the log-scale reflects that $\mathcal{R}_0^{\mathrm{u}}(t)$ is a non-negative quantity, and our belief that log $[\mathcal{R}_0^{\mathrm{u}}(t)]$ has no long-term stationary behavior, but that its log-scale weekly changes do. We bound $\beta$ to be between 0 and 1

$$0 < \beta < 1$$

Such that at $\beta = 0$ the dynamics revert to a random walk. This $\mathcal{R}_0^{\mathrm{u}}(t)$ is represented by the gray line in Fig. 1A denoted "Reference $\mathcal{R}(t)$".

##### *Subpopulation-level unadjusted effective reproductive number $\mathcal{R}_k^{u}(t)$*

We introduce hierarchical coupling of the subpopulation infection dynamics by modeling the unadjusted reproductive numbers of each subpopulation as deviations from the un-damped instantaneous reproduction number in the reference subpopulation, $\mathcal{R}_0^{\mathrm{u}}(t)$. This approach models the other subpopulations as having infection dynamics that are *a priori* believed to be *similar* to one another but can differ from the reference subpopulation dynamics.

The deviations of each subpopulation unadjusted reproductive number $\mathcal{R}_k^{\mathrm{u}}(t)$ from the reference subpopulation unadjusted reproductive number $\mathcal{R}_0^{\mathrm{u}}(t)$ are modelled as a log-scale AR(1) process. The subpopulation level reproductive numbers $\mathcal{R}_k^{\mathrm{u}}(t)$ are represented by the blue and red lines in the "Subpopulation $\mathcal{R}_k(t)$" figure in Fig. 1A.

Specifically, for subpopulation $k$:

$$\log[\mathcal{R}_k^{\mathrm{u}}(t)] = \log[\mathcal{R}_0^{\mathrm{u}}(t)] + m + \delta_k(t)$$

where $m$ is an "intercept" for the reference subpopulation, which is a fixed parameter and allows for the fact that $\log[\mathcal{R}_0^{\mathrm{u}}(t)\ ]$ may differ from the central dynamic by a constant value. An AR(1) process was chosen to impose some degree of correlation between subsequent deviations from the central dynamic, i.e. if a subpopulation had a $\log[\mathcal{R}_k^{\mathrm{u}}(t)]$higher than the central dynamic last week, it is likely to also be higher from the central dynamic the following week.

The time-varying subpopulation effect on $\log[\mathcal{R}_0^{u}(t)]$, $\delta_k(t)$ is modeled as:

$$\delta_k(t) = \varphi_{R(t)}\delta_k(t-1) + \epsilon_{kt}$$

where $\varphi_{R(t)}$ is an autoregression coefficient which serves to make week-to-week deviations from the central dynamic correlated within a subpopulation. We bound $\varphi_{R(t)}$ to be between 0 and 1

$$0 < \varphi_{R(t)} < 1$$

such that the successive weeks change in deviation from the log scale reference effective reproductive number $\log[\mathcal{R}_0^{\mathrm{u}}(t)]$ are correlated to a degree determined by $\varphi_{R(t)}$ , and that the long-term behavior is stationary. We model the random deviations $\epsilon_{kt}$ as

$$\epsilon_{kt} \sim \mathrm{Normal}\left(0, \sigma_{R(t)\delta}\right)$$

where $\sigma_{R(t)\delta}$ determines the magnitude of the weekly variation from the global dynamics. At $\varphi_{R(t)} = 0$ there is no auto-correlation in week-to-week deviations, and the weekly value of $\log[\mathcal{R}_k^{\mathrm{u}}(t)]$ normally distributed around $\log[\mathcal{R}_0^{\mathrm{u}}(t)] + m$. As $\sigma_{R(t)\delta}$ approaches 0 the subpopulation-to-subpopulation deviations approach 0.

The hierarchical structure of the sub-population reproductive numbers allows infection dynamics in each subpopulation to deviate away from the central dynamics if the data suggests it but enforces them ultimately to regress back toward the inferred central dynamic (3,4).

#### *Subpopulation-level effective reproductive number $\mathcal{R}_k(t)$ via infection feedback*

As mentioned in the high-level model summary (see main text Methods, Model: High-level overview), we constrain growth in infection incidence by defining the effective reproductive number for each subpopulation as a combination of the unadjusted reproductive number and an "infection feedback" damping term (5).

There exist various mechanistic approaches to model reductions in current infections because of high levels of past transmission. Other methods explicitly model susceptibles to account for effective susceptible depletion (1,4,6), both of which are alternate methods for turnover at high infection rates that we plan to explore. (Asher 2018). For each subpopulation k the effective reproductive number was modeled as:

$$\mathcal{R}_k(t) = \mathcal{R}_k^{\mathrm{u}}(t)\exp\left(-\gamma \sum_{\tau=1}^{\tau=T_G} I_k\,(t-\tau)g(\tau)\right)$$

where $\mathcal{R}_k^{\mathrm{u}}(t)$ is the unadjusted effective reproductive number for subpopulation k, $I_k(t)$ is the infection process in subpopulation k, $g(\tau)$ is the discrete generation interval, used here to impose the assumption that the time scale of infection feedback would occur on the time scale of secondary transmission, $\gamma$ is the infection feedback term controlling the strength of the damping on $\mathcal{R}_k(t)$ and $T_g$ is the maximum generation interval, which is the maximum time from infection to secondary infection that we consider, and is set to 15 days. We note that the timing of the infection feedback damping term, which here is assumed to occur on the time scale of secondary transmission, differs from the implementation in (5), which is assumed to occur directly on the time scale of incident infections.

### B. Infection process

#### *Subpopulation-level infections*

We model latent incident infections per capita in each subpopulation $I_k(t)$ (Fig. 1B) as generated from a deterministic renewal process (1,2,6–9):

$$I_k(t) = \mathcal{R}_k(t) \sum_{\tau=1}^{\tau=T_G} I_k\,(t-\tau)g(\tau)$$

Where $g(\tau)$ is the discrete generation interval, which describes the distribution of times from incident infection to secondary infection (i.e. infectiousness profile) and $\mathcal{R}_k(t)$ is the instantaneous reproduction number, representing the expected number of secondary infections occurring at time $t$, divided by the number of currently infected individuals, each scaled by their relative infectiousness at time $t$ (10). The incident infections in each subpopulation are represented by the red and blue lines labeled "Incident infections in subpopulation $k = 1, I_k(t)$" in Fig. 1B.

#### Infection initialization

This process is initialized by estimating an initial exponential growth rate (1) of infections for $\tau_{i0}$ days prior to the calibration start time $t_0$, the first day of observations:

$$I_k(t = t_0) = I_k(t = t_0 - \tau_{i0})exp(r_k t)$$

where $I_k(t = t_0 - \tau_{i0})$ is the initial per capita incident infections at $\tau_{i0}$ days before the first observed data and $r_k$ is the site-level exponential growth rate during that period. We infer the site level initial per capita incidence $I_k(t = t_0 - \tau_{i0})$ and the site-level exponential growth rate $r_k$ hierarchically. Specifically, we treat $\text{logit}[I_k(t = t_0 - \tau_{i0})]$ as Normally distributed about the corresponding global value $\text{logit}[I(t = t_0 - \tau_{i0})]$, with an estimated logit-scale standard deviation $\sigma_{i0}$:

$$\text{logit}[I_k(t = t_0 - \tau_{i0})] \sim \text{Normal}(\text{logit}[I(t = t_0 - \tau_{i0})], \sigma_{i0})$$

and we treat $r_k$ as Normally distributed about the corresponding global value $r$, with an estimated standard deviation $\sigma_{\text{growth}}$

$$r_k \sim \text{Normal}(r, \sigma_{\text{growth}})$$

We initialize the incident infections with an exponential growth process for a total of $\tau_{i0} = 55$ days. This is set to ensure we have a sufficient duration of incident infections in the unobserved time period to produce expected observations during the periods of observed data.

#### *Global infections*

To obtain the number of infections per capita $I(t)$ in the total population, we sum the $K_{\text{total}}$ subpopulations per capita infection counts $I_k(t)$ weighted by their population sizes, $n_k$:

$$I(t) = \frac{1}{\sum_{k=0}^{K_{\text{total}}-1} n_k} \sum_{k=0}^{K_{\text{total}}-1} n_k \, I_k(t)$$

Where $k = 0$ refers to the reference subpopulation and $k = 0 \ldots K_{total} - 1$ the $k$th subpopulations representing the wastewater catchment areas. This implementation has the effect of essentially weighing the impact of the signal from a wastewater catchment area proportional to its relative population size. This is indicated in Fig 1B by the dashed-lined rectangle to represent that the subpopulation level incident infections (blue and red lines) are summed to get the total incident infections in the population. We can use the total per capita incident infections $I(t)$ to back-calculate the realized global $\mathcal{R}(t)$ (see S1 Text Model Definition: Back calculation of the global effective reproductive number $\mathcal{R}(t)$).

### C. Observation process

We model two types of observations downstream of latent infections: hospital admissions coming from incident infections from the total population, and wastewater concentrations coming from individual sites, processed in individual labs, coming from incident infections in each subpopulation (Fig.1C).

#### *Wastewater generative model*

#### Viral genome concentration in wastewater

Latent incident infections in subpopulation $k$ are mapped to their corresponding site $i$, such that $i = k$ for all $n_{sites}$ . We model viral genome concentrations in wastewater in site $i$, $C_i(t)$ as a convolution of the expected latent incident infections per capita in the corresponding subpopulation $k$, $I_k(t)$ and a normalized shedding kinetics function $s(\tau)$, multiplied by $G$ the number of genomes shed per infected individual over the course of their infection and divided by $\alpha$ the volume of wastewater generated per person per day:

$$C_i(t) = \frac{G}{\alpha} \sum_{\tau=0}^{\tau=\tau_{shed}} I_k\,(t-\tau)s(\tau)$$

where $\tau_{\text{shed}}$ is the total duration of fecal shedding. This approach assumes that $G$ and $\alpha$ are constant through time and across individuals. In fact, there is substantial inter-individual variability in shedding kinetics and total shedding. This approximation is more accurate when population sizes are large.

We model the shedding kinetics $s(\tau)$ as a discretized, scaled triangular distribution (11).

$$\log_{10}[s^{\text{cont}}(\tau)] = \begin{cases} V_{\text{peak}} \dfrac{\tau}{\tau_{\text{peak}}} & \tau \leq \tau_{\text{peak}} \\ V_{\text{peak}} \left(1 - \dfrac{\tau - \tau_{\text{peak}}}{\tau_{\text{shed}} - \tau_{\text{peak}}}\right) & \tau_{\text{peak}} < \tau \leq \tau_{\text{shed}} \\ 0 & \tau > \tau_{\text{shed}} \end{cases}$$

where $V_{\text{peak}}$ is the peak number or viral genomes shed on any day of infection, $\tau_{\text{peak}}$ is the time in days from infection to peak shedding, and $\tau_{\text{shed}}$ is the total duration of shedding.

#### Wastewater concentration observation model

Viral genome concentration measurements can vary between sites because of differences in sample collection and lab processing methods. We account for systematic differences in the magnitude between the expected concentration in a site, $C_i(t)$, via a scaling factor $M_{ij}$, i.e. the expected observed concentration processed in site $i$ and lab $j$ at time $t$ is $M_{ij}C_i$. We account for differences in the variability in the measurement via a site-lab level observation error term $\sigma_{cij}$. We model the observed viral genome concentrations $c_{ijt}$ from site $i$ processed in lab $j$ sampled on day $t$ as lognormally distributed:

$$\log[c_{ijt}] \sim \text{Normal}(\log[M_{ij}C_i(t)], \sigma_{cij})$$

where $M_{ij}$ and $\sigma_{cij}$ are estimated hierarchically such that:

$$\log[M_{ij}] \sim \text{Normal}(0, \sigma_m)$$

such that $M_{ij}$ is centered around 1 with a log standard deviation $\sigma_m$ and the log standard deviation in the wastewater observation process $\sigma_{cij}$ is defined as:

$$\log(\sigma_{cij}) \sim \text{Normal}(\log(\hat{\sigma}_c, \sigma_{\log\sigma_c})$$

Where$\hat{\sigma}_c$and $\sigma_{\log\sigma_c}$ are the log mean and log standard deviation of the site-lab level observation model log standard deviation, respectively. See Priors and parameters section below for prior values on these hyperparameters.

The resulting observed site-lab level concentrations in each subpopulation are represented by the blue and red lines in Fig. 1C figure labeled "Wastewater concentration in site i=1, lab j = 1". In the rare cases when a site submits multiple concentrations for a single date and lab, we treat each record as an independent observation.

#### Censoring wastewater observations below the limit of detection

Lab processing methods have a finite limit of detection (LOD), such that not all wastewater measurements can be modeled using the log-normal approach described above. This limit of detection varies across sites, between methods, and potentially also over time.

If an observed value $c_{ijt}$ is above the corresponding LOD, then the likelihood of the wastewater observation is:

$$f_{\text{Normal}}(\log[c_{ijt}]; \log[M_{ij}C_i(t)], \sigma_{cij})$$

where $f_{\text{Normal}}(x; \mu, \sigma)$ is the probability density function of the Normal distribution.

When the observed value is below the LOD, we use the censored likelihood:

$$\int_{-\infty}^{\log\text{LOD}_{ijt}} f_{\text{Normal}}\,(x; \log[M_{ij}C_i(t)], \sigma_{cij})\text{d}x$$

### *Hospital admissions generative model*

#### Hospital admissions model

We model the expected hospital admissions per capita $H(t)$ in the total population as a convolution of the expected latent incident infections per capita $I(t)$, and a discrete infection to hospitalization distribution $d(\tau)$, scaled by the probability of being hospitalized $p_{\text{hosp}}(t)$ and a weekday effect $\omega(t)$

$$H(t) = \omega(t)p_{\text{hosp}}(t)\sum_{\tau=0}^{\tau=T_d} d\,(\tau)I(t-\tau)$$

Where $T_d$ is the maximum delay from infection to hospitalization that we consider and $\omega(t)$ is a weekday effect to account for day-of-week effects in hospital reporting. If $t$ and $t'$

are the same day of the week, $\omega(t) = \omega(t')$. The seven values that $\omega(t)$ takes on are constrained to be non-negative and have a mean of 1. This allows us to model the possibility that certain days of the week could have systematically high or low admissions reporting while holding the predicted weekly total reported admissions constant (i.e. the predicted weekly total is the same with and without these day-of-week reporting effects).

We define the discrete hospital admissions delay distribution $d(\tau)$ as a convolution of the incubation period distribution (12) and a separate estimate of the distribution of time from symptom onset to hospital admission (see S1. Text Model Definition: Distributional parameters for specific values and an explanation of how this was generated).

#### Infection-hospitalization rate

In the wastewater-informed model, we allow the population-level infection-hospitalization rate (IHR) to change over time. An inferred change in the IHR could reflect either a true change in the rate at which infections result in hospital admissions (e.g. the age distribution of cases could shift, a more or less severe variant could emerge, or vaccine coverage could shift) or a change in the relationship between infections and genomes shed in wastewater $G$ (which we currently treat as fixed, but which could change in time if, for example, immunity reduces wastewater shedding without reducing transmission, or a variant emerges with a different per infection wastewater shedding profile).

Therefore, we model the proportion of infections that give rise to hospital admissions $p_{\text{hosp}}(t)$ as a piecewise-constant function with weekly change points.

If $t$ and $t'$ are two days in the same week, then $p_{\text{hosp}}(t) = p_{\text{hosp}}(t')$.

The values $p_{\text{hosp}}(t)$ follow a logit-scale AR(1) process with linear scale median $\mu_{p_{\text{hosp}}}$:

$$\text{logit}\left[p_{\text{hosp}(t)}\right] = \text{logit}\left[\mu_{p_{\text{hosp}}}\right] + \delta_H(t)$$

where $\delta_H(t)$ is the time-varying site effect on $\text{logit}\left[p_{\text{hosp}}(t)\right]$, modeled as:

$$\delta_H(t) = \varphi_H \delta_H(t-1) + \epsilon_{Ht}$$

where $0 < \varphi_H < 1$ and

$$\epsilon_{Ht} \sim \text{Normal}(0, \sigma_{H\delta})$$

As $\sigma_{H\delta}$ approaches 0, the week-to-week variation in the IHR approaches 0, reverting to a constant IHR model.

#### Hospital admissions observation model

We model the observed hospital admissions counts $h_t$ as negative binomially distributed

$$h_t \sim \text{NegBinom}(nH(t), \phi)$$

with a mean of the product of the expected hospital admissions per capita in the jurisdiction and the total population size in the jurisdiction, $n$, i.e. $nH(t)$, and a dispersion parameter $\phi$. This is represented by the gray line in the figure denoted "Total Hospital admissions" in Fig. 1C.

#### Back calculation of the global effective reproductive number

The total per capita incident infections can be used to back-calculate the jurisdiction-level "realized" effective reproductive number, since we estimate only the unadjusted effective reproductive number as part of the hierarchical structure described above.

#### Subpopulation definition

The total population consists of $K_{\text{total}}$ subpopulations $k$ with corresponding population sizes $n_k$. We associate one subpopulation to each of the $K_{\text{sites}}$ wastewater sampling sites in the jurisdiction and assign that subpopulation a population size $n_k$ equal to the wastewater catchment population size reported for that wastewater catchment area.

Whenever the sum of the wastewater catchment population sizes $\sum_{k=1}^{K_{\text{sites}}} n_k$ is less than the total population size $n$, we use an additional subpopulation of size $n - \sum_{k=1}^{K_{\text{sites}}} n_k$ to model individuals in the population who are not covered by wastewater sampling.

The total number of subpopulations is then $K_{\text{total}} = K_{\text{sites}} + 1$: the $K_{\text{sites}}$ subpopulations with sampled wastewater, and the final subpopulation to account for individuals not covered by wastewater sampling. The model without wastewater (hospital admissions only model) is therefore a special case of the model where $K_{\text{sites}} = 0$ and $K_{\text{total}} = 1$, with subpopulation size $n_k = n$, the total population. In the case where the sum of the wastewater site catchment populations meets or exceeds the total population ($\sum_{k=1}^{K_{\text{sites}}} n_k \geq n$) the model does not use a final subpopulation without sampled wastewater. In that case, the total number of subpopulations $K_{\text{total}} = K_{\text{sites}}$.

This amounts to modeling the wastewater catchment populations as approximately non-overlapping; every infected individual either does not contribute to measured wastewater or contributes principally to one wastewater catchment. This approximation is reasonable if we restrict our analyses to primary wastewater treatment plants, which avoids the possibility that an individual might be sampled once in a sample taken upstream and then sampled again in a more aggregated sample taken further downstream.

When converting from predicted per capita incident hospital admissions $H(t)$ to predicted hospitalization counts, we use the jurisdiction population size $n$, even in the case where $\sum n_k > n$.

This amounts to making two key additional modeling assumptions:

- Any individuals who contribute to wastewater measurements but are not part of the total population are distributed among the catchment populations approximately proportional to catchment population size.

- Whenever $\sum n_k \geq n$, the fraction of individuals in the jurisdiction not covered by wastewater is small enough to have minimal impact on the jurisdiction-wide per capita infection dynamics.

The hierarchical structure linking infection dynamics in each subpopulation to a central or "global" dynamic is implemented using a reference subpopulation. The reference subpopulation is by default the subpopulation not covered by wastewater, or in the case where the sum of the wastewater site catchment populations meet or exceed the total population ($\sum_{k=1}^{K_{\text{sites}}} n_k \geq n$), the reference subpopulation is by default the wastewater catchment area with the largest population size.

### Priors and Parameters

| Parameter | Prior distribution | Source |
|---|---|---|
| Initial hospitalization probability | $\text{logit}[p_{\text{hosp}}(t_0)] \sim \text{Normal}(\text{logit}[0.01], 0.3)$ | (13) |
| Time to peak fecal shedding | $\tau_{\text{peak}} \sim \text{Normal}(5 \text{ days}, 1 \text{ day})$ | (14–16) |
| Peak viral shedding $V_{\text{peak}}$ | $\log_{10}[V_{\text{peak}}] \sim \text{Normal}(5.1, 0.5)$ | (17) |
| Duration of shedding | $\tau_{\text{shed}} \sim \text{Normal}(17 \text{ days}, 3 \text{ days})$ | (16,18) |
| Total genomes shed per infected individual | $\log_{10}[G] \sim \text{Normal}(9,2)$ | (19) |
| Initial infections per capita $I_0$ | $I_0 \sim \text{Beta}(1 + k i_{\text{est}}, 1 + k(1 - i_{\text{est}}))$ | where $i_{\text{est}}$ is the sum of the last 7 days of hospital admissions, divided by jurisdiction population, and divided by the prior mode for $p_{\text{hosp}}$, and $k = 5$ is a parameter governing the informativeness ("certainty") of the Beta distribution |
| Initial exponential growth rate | $r \sim \text{Normal}(0, 0.01)$ | Chosen to assume flat dynamics prior to observations |
| Infection feedback term | $\gamma \sim \text{logNormal}(4.50, 0.64)$ | Weakly informative prior chosen to have a mode of 90 in natural scale, |

| Parameter | Prior distribution | Source |
|---|---|---|
| | | based on posterior estimates of the infection feedback from model fits from March 2023 to March 2024 in Alaska, Massachusetts, New Jersey, New Hampshire, and Washington. |
| Day of the week effects | $\frac{\vec{\omega}}{7} \sim \text{Dirichlet}(5,5,5,5,5,5,5)$ | Weakly informative prior with a mode at even daily reporting (no effects) |
| Standard deviation of the log of the site-lab level multiplier $M_{ij}$ | $\sigma_m \sim \text{Normal}(0,0.25)$ | Weakly informative prior chosen to allow average magnitude of concentrations to be either similar or different among individual sites, depending on data |
| Modal site-level observation standard deviation | $\hat{\sigma}_c \sim \text{Normal}(1,1)$ | Weakly informative prior chosen to allow the mode to be either small or large |
| Standard deviation of the Normal distribution of individual log observation standard deviations $\log(\sigma_{\text{cij}})$ (site-lab combination specific, with an inferred modal standard deviation $\hat{\sigma}_{\text{c}}$) | $\sigma_{\log\sigma_c} \sim \text{Normal}(0, \log(2))$ | Weakly informative prior which allows for individual standard deviations to be either clustered around the modal standard deviation or more dispersed |

Table of prior values. We use informative priors for parameters that have been well characterized in the literature and weakly informative priors for parameters that have been less well characterized.  Full set of prior hyperparameters can be found at https://github.com/CDCgov/ww-inference-model/blob/v0.1.1/inst/extdata/example_params.toml .

| Parameter | Value | Source |
|---|---|---|
| Maximum generation interval | $T_g = 15$ days | |
| Maximum infection to hospital admissions delay | $T_d = 55$ days | |
| Wastewater produced per person-day | $\alpha = 378{,}500$ mL per person-day | (20) |

Table of scalar parameters. These are passed in as set values to the model.

#### *Discrete distributional parameters*

The generation interval probability mass function $g(\tau)$ approximates a log-normal distribution (12) with log-mean 2.9 and log-standard deviation of 1.64, chosen as currently circulating lineages are descendants of the Omicron lineage (which has a shorter generation time than the original strain). To approximate the double censoring process necessary to discretize the continuous log-normal distribution, we use a simulation-based approach as recommended by (21,22). This assumes that the primary event is uniformly distributed within the primary censoring interval(this ignores the influence of the growth rate within the primary interval but is a good approximation in most settings). The secondary event is then a sum of this primary interval and the continuous distribution and is observed within a day (see Figure 9 in (22)). As the renewal process is not defined if there is probability mass on day zero we further left truncate this distribution at 1.

We derive the distribution $\delta(\tau)$ of the delay from infection to hospital admission as the sum of the incubation period (delay from infection to symptom onset) and the period from symptom onset to hospital admission.

We model the incubation period with a discretized, Weibull distribution adjusted from the observed backwards distribution to the forwards distribution using the exponential growth rate to account for the estimate taking place during a period of exponential growth after the emergence of the Omicron lineage (12), with probability mass function $\delta(\tau)$:

$$\delta^{\text{cont}}(\tau) = \exp[0.15\tau] f_{\text{Weibull}}(\tau; \text{shape} = 1.5, \text{scale} = 3.6)$$

$$\delta(\tau) = \begin{cases} \delta^{\text{cont}}(\tau) / \left( \sum_{\tau'=0}^{23} g^{\text{cont}}(\delta') \right) & 0 \leq \tau \leq 23 \\ 0 & \text{otherwise} \end{cases}$$

We model the symptom onset to hospital admission delay distribution with a Negative Binomial distribution with probability mass function $\gamma(\tau)$ fit to line list patient data from (23).

$$\gamma(\tau) = f_{\text{NegBin}}(\tau; 6.99 \text{ days}, 2.49 \text{ days})$$

The infection-to-hospitalization delay distribution $d(\tau)$ is the convolution:

$$d(\tau) = \sum_{x=0}^{\tau} \delta(x)\gamma(\tau - x)$$

This resulting infection to hospital admission delay distribution has a mean of 12.2 days and a standard deviation of 5.67 days.

### S4. Text Criteria for forecast inclusion

#### *Sufficient wastewater*

We define a jurisdiction as having "sufficient" wastewater data if the jurisdiction has at least one site and lab pair with more than 5 observations and if the jurisdiction's most recent wastewater observation is less than 21 days from the forecast date.

#### *Convergence criteria*

Additionally, we required model fits to pass pre-specified MCMC convergence diagnostic thresholds: less than 5 percent of sampling iterations resulting in divergent transitions, an Expected Bayesian Missing Information (EBMFI) of less than 20 percent, and less than 5 percent of the model parameters with an $\hat{R}$ (a measure of the mixing of the chains) of greater than 1.05.

#### *Manual review process*

For real-time forecast production, we had a manual review process as part of our real-time production workflow (see below section, "Real-time forecast production"). If the wastewater-informed model's forecast appeared unreliable or implausible (with an initial review followed by a group discussion and final decision), we replaced it with the hospital admissions-only model.

#### *Application of inclusion criteria: real-time*

In real-time, when we produced the Hub submission labeled `cfa-wwrenewal`, we excluded wastewater-informed forecasts that failed to meet the convergence criteria and/or failed to pass the manual review process. These forecasts were replaced with the hospital admissions-only model's forecasts. These decisions were documented in our GitHub repository (24). See https://github.com/CDCgov/wastewater-informed-covid-forecasting.blob/7c30b603f5e57f2b728f23506fe7714d576c31c9/forecasts/2024-02-05/metadata.yaml for an example from February 5th, 2024.

For the real-time comparison of models with and without wastewater, we excluded wastewater-informed forecasts that were excluded in real-time due to convergence issues or manual review or had insufficient wastewater data, based on the criteria above.

#### *Application of inclusion criteria: retrospective*

In the retrospective analysis comparing forecast performance with and without wastewater data, we excluded all forecast date-location combinations that failed to meet the criteria for model convergence and sufficient wastewater.

In the retrospective comparison to the Hub submissions, we excluded all forecast date-location combinations that failed to meet the criteria for model convergence and sufficient wastewater. These forecasts were replaced with forecasts from the hospital admissions-only model.

### S5. Text Further details on forecast evaluation

#### *Selection of COVID-19 Forecast Hub submissions for comparison*

We compared our models' performance to that of models submitted to the COVID-19 Forecast Hub in real-time. We included models with submissions to the Hub on 20 or more of the 24 forecast dates from October 16, 2023 through March 25, 2024. To count toward the 20-forecast-date minimum, a submission had to include forecasts for 40 or more of the 52 jurisdictions.

We used the `zoltr` package (25) to query forecast submissions to identify models meeting these criteria. Those models were: `CEPH-Rtrend_covid`, `CMU-TimeSeries`, `COVIDhub-4_week_ensemble`, `COVIDhub-baseline`, `MUNI-ARIMA`, `SGroup-RandomForest`, `UMass-gbq`, `UMass-sarix`, `UMAss-trends_ensemble`, and `UT-Osiris`. We then pulled the forecast files for those models directly from the COVID-19 Forecast Hub GitHub (26,27). To reduce visual complexity, we mainly display the results for the top 2 individual models by overall average WIS across the 2023-24 season and the COVID Hub ensemble alongside our models in main text figures. Rankings were computed using all the models listed above, and all are included as part of the supplemental results.

To quantify the distribution of ranks of each model, we computed the relative rank of the average WIS across horizon days: the worst possible score is the reciprocal of the total number of models; the best possible score is 1. This approach can tell us if certain models tend always to rank similarly, or if some models perform better or worse relative to their peers at certain times.

### Further details on retrospective comparison of forecasts with and without wastewater

#### *Analysis of scored retrospective forecasts*

To understand the distribution of relative performance of the wastewater-informed model compared to the hospital admissions-only model, we compute relative scores by taking the arithmetic mean for each model across the dimension of interest and then dividing by the corresponding hospital admissions-only model score. To understand the magnitude of forecast performance differences across forecast dates and put the relative forecast performance in context, we compute the mean CRPS across all jurisdictions for each forecast date for both models.

To assess the calibration performance of each model, we calculate and visualize quantile coverage for the Hub quantiles (26,27), averaged across each forecast date, jurisdiction, and horizon day for both models. We calculate and visualize interval coverage for the 30 percent, 60 percent, and 90 percent prediction intervals stratified by horizon (nowcast period and 1 through 4 weeks ahead) for both models (Figure S13).

### S6. Text: Exploratory investigation of drivers of prediction and forecast performance heterogeneity

To generate hypotheses as to why and when the wastewater-informed model performs better or worse than the hospital admissions only model, we performed a strength-of-trend analysis on the recent wastewater and hospital admissions trends and their impact on the divergence in forecast predictions and the divergence in score between the two models. The goal of this analysis was to better understand and explain how the wastewater-informed model uses wastewater and to inform future model development.

To assess the correlates of reduced and improved model performance, we used Bayesian regression computed the following metrics for each retrospective forecast problem:

- The magnitude and direction of recent incident hospital admissions trends in the input data, quantified as an inferred slope relating time to log incident admissions.
- The magnitude and direction of recent wastewater trends in the input data, quantified as an inferred slope relating time to log viral genome concentration.
- The uniformity across sampling sites of recent wastewater trends in the input data, quantified as an inferred standard deviation of trend slopes across the individual wastewater sampling sites in a jurisdiction.

We inferred trends over the 23 days prior to the forecast date. Given the 9 day data latency in hospital admissions data, this corresponds to the last two weeks of available admissions data when the forecast was made.

We used `brms` (30)to specify and fit Bayesian regression models. In the discussion that follows, we use the same probabilitistic modeling notation and probability distribution parametrizations as we use to specify our main model.

For the hospital admission trends, we fit a simple single-level log-linear regression model with a negative binomial observation process and time as the sole predictor. $H(t)$ denotes predicted admissions. $h_t$ denotes observed admissions. Time is measured relative to the first datapoint and one-indexed (i.e. t = 1 is 23 days before the forecast date). We used the following model structure:

$$\log[H(t)] = \alpha_h + \beta_h t$$

$$h_t \sim NegativeBinomial(H(t),\ p_h)$$

$\beta_h$ is the inferred slope, which we use to quanify the magnitude and direction of the recent admissions trend. $\alpha_h$is an inferred intercept and $\phi_h$is an inferred concentration for the Negative Binomial obervation process.

We used the following priors:

$$\beta_h \sim Normal(0,\ 0.06)$$

$$\log[\phi_h] \sim Normal(\log[10],\ \log[10])$$

We used the brms default prior for the intercept $\alpha_h$.

For the wastewater trends, we used a hierarchical model across sampling sites with a censored Normal observation process for the observed log genome concentrations. $C_{ij}(t)$denotes the predicted concentration for site i and lab j at time t. $c_{ijt}$ denotes the observed concentration. Time is measured relative to the first datapoint and one-indexed (i.e. t = 1 is 23 days before the forecast date).

$$\log\left[c_{ijt}\right] \sim LeftCensoredNormal\left(\log\left(C_{ij}(t)\right), \sigma_{ij}, LOD_{ij}\right)$$

$$\log\left[C_{ij}(t)\right] = \alpha_{cij} + \beta_{cij} t$$

$$\alpha_{cij} \sim Normal(\alpha_c,\ \sigma_\alpha)$$

$$\beta_{cij} \sim Normal\left(\beta_c, \sigma_\beta\right)$$

$$\log\left[\sigma_{cij}\right] \sim Normal(\log[\sigma_c], s_\sigma)$$

The $\beta_{cij}$are site-lab specific slopes; they are distributed about an inferred global slope $\beta_c$ with an inferred standard deviation $\sigma_\beta$. We use the global slope $\beta_c$ to quantify the magnitude and direction of the overall wastewater trend. We use the inferred hierarchical standard deviation $\sigma_\beta$ to quantify the uniformity of the trend. The less variable the site-lab specific slopes $\beta_{cij}$ are about the global slope $\beta_c$ (smaller $\sigma_\beta$), the more uniform the trend across site-labs.

The $\alpha_{cij}$ are inferred site-lab-specific intercepts, distributed about a global intercept $\alpha_c$ with an inferred standard deviation $\sigma_\alpha$. The $\sigma_{cij}$ are inferred lab-site-specific observation standard deviations. They are log-Normally distributed about an inferred global observation standard deviation $\sigma_c$. That log-Normal for the observation standard deviations itself has an inferred standard deviation, $s_\sigma$.

We used the following priors. PosNormal(mu, sigma) denotes a positive-constrained Normal distribution with mode mu and standard deviation sigma.

$$\beta_c \sim Normal(0,\ 0.06)$$

$$\alpha_c \sim Normal(7,\ 4)$$

$$\sigma_\beta \sim PosNormal(0,\ 0.03)$$

$$\sigma_\alpha \sim PosNormal(0,\ 1)$$

$$\log[\sigma_c] \sim Normal(\log[1], \log[2])$$

$$\log[s_\sigma] \sim Normal(\log[1],\ \log[1.5])$$

We fit models using Stan's No-U-Turn sampler with the same configuration that we used for the forecast model fits (4 chains, each with 750 warmup iterations and 500 sampling iterations, yielding 2000 posterior samples).

To quantify the divergence in predictions: we computed the mean of the posterior of the log difference in the sum of the forecasted hospital admissions across the nowcast and forecast period. To quantify the relative model performance, we computed the difference between the log of the average CRPS from the wastewater-informed model and the hospital admissions only model.

### S7. Text Additional results

#### *Real-time forecast performance with and without wastewater*

We produced forecasts from both models in real-time for 12 forecast dates from February 5, 2024, to April 29, 2024 for all 52 jurisdictions each week. Only 8 of these forecast dates have complete evaluation data (i.e. reported admissions data for all 28 days following the forecast date). From these 8 dates, we therefore have 52 * 8 = 416 potential real-time forecasts (forecast date-jurisdiction pairs) available for analysis. Of these, 42 wastewater-informed forecasts were not submitted in real-time due to model convergence issues or rejection on manual review (Fig. S2 & S3). The hospital admissions-only model was substituted in 41 cases, in 1, we elected not to submit either model. An additional 54 lacked sufficient wastewater data to inform a forecast (31 lacked any wastewater data, 23 did not have recent data). That leaves 320 wastewater-informed forecasts (and 320 associated hospital admissions-only forecasts) for evaluation of real-time relative performance with and without wastewater data.

Across those 320 forecast pairs, the wastewater-informed model and the hospital admissions-only model had almost identical Weighted Interval Scores (WIS): 0.282 for the wastewater-informed model and 0.281 for the hospital admissions-only model, relative WIS (rWIS) = 1.00 (Fig. S11 A & B, Fig. S14). An rWIS value of less than 1 indicates that the wastewater-informed model outperformed the hospital admissions-only model, while a value of greater than 1 indicates that it performed worse. The distribution of log relative WIS for the 320 forecast pairs is approximately symmetrical around 1, with many instances of near-equivalent performance as well as some instances in which one model substantially outperformed the other (Fig. S11B).

The two models performed similarly by WIS on each of the 8 forecast dates (Fig. S11 D, E, F). Performance for the wastewater-informed model relative to the hospital admissions-only model ranged from 1.08x better to 1.10x worse. The wastewater-informed model performed better on 3 of the 8 forecast dates and worse on the other 5.

Stratifying by jurisdiction, there were 48 jurisdictions with paired forecasts, and 46 with paired forecasts for 4 or more of the 8 forecast dates. Among those 46 jurisdictions, there was substantially heterogeneity in relative forecast performance by WIS: the wastewater-informed model performed from 1.54x better to 1.57x worse than the admissions-only model. The wastewater-informed model performed better by WIS than the hospital admissions-only model for 27 of 46 jurisdictions and worse for the other 19 (Fig. S16).

Both models were biased towards overprediction (Fig. S12), but the wastewater-informed model was less biased than the hospital admissions only model (average bias of 0.283 and 0.382, respectively, Fig. S12). The hospital admissions-only model was modestly better calibrated than the wastewater-informed model across the central 30$^{th}$, 60$^{th}$, and 90$^{th}$ prediction intervals (Fig. S13). Both models were overconfident (empirical interval coverage worse than theoretical), particularly at longer forecast horizons (Fig. S13).

#### *Visual and quantitative evaluation of forecast performance across three example jurisdictions*

In the main text, we showed example forecasts from California, Virginia, and Washington for January 15, 2024 (Fig. 3), because incorporating wastewater data improved, had little impact on, and reduced forecast performance, respectively in those locations on that forecast date. Across forecast dates in California, the wastewater-informed model performed 1.34 times better than the hospital admissions-only model (rCRPS = 0.739 [0.118/0.160] Fig. S4 A, B, C). In Virginia, the wastewater-informed model performed 1.17 times worse than the admissions-only model (rCRPS=1.17 [0.224/0.190], Fig. S5, Fig. S1 D, E, F). In Washington, the wastewater-informed model performed 1.48 times worse (rCRPS = 1.48 [0.278/0.188] Fig. S4 G, H, I).

### S8. Text: Additional limitations

#### Limitations of the analysis

- We did not have vintaged wastewater data available from prior seasons, and required reporting of daily incident hospital admissions paused in May of 2024. Our retrospective and real-time evaluations thus span a short time window (9 months for the retrospective analysis and 8 weeks for the real-time analysis), covering only a single epidemic peak, which limits the generality of conclusions that can be drawn from our results. In future analyses, we would have iterated on our model using a subset of the data and then presented the results of this iteration on a held-out set of test data.

- The model we used to produce real-time forecasts changed throughout the submission period and is likewise different from the version of the model we used to produce the retrospective forecasts (S5. Text Model changes over time). This issue was compounded by not keeping all versions of our approach readily available meaning that without significant effort we cannot easily rerun earlier iterations of the model.

- Our analysis was also constrained by fixed data reporting delays, with hospital data only available up to 10 days prior to the forecast date. We did not explore scenarios with shorter or longer reporting delays, which could impact model performance, particularly for the hospital admissions-only model.

- Our comparison of the wastewater-informed model versus the hospital admissions-only model excluded forecasts for which the model had convergence issues, which occurred more frequently in the wastewater-informed model. This might lead to a bias in one direction or the other as this may systematically exclude poor forecasts from one of the models, and we did not investigate this further.

- Similarly, we did not investigate how varying the frequency and timeliness of wastewater reporting might affect the relative performance of models incorporating wastewater data.

- Our real-time forecasts involved additional manual review of input data and forecast outputs, which we did not attempt to simulate for the retrospective analysis, not least because an unbiased manual review of forecasts for robustness and plausibility is not possible when researchers already know the true outcome

- Our model code and its associated R package are not readily extensible despite our best efforts due to the challenges and resource requirements involved in developing software projects of this kind.

- We reused many elements from the `EpiNow2` R package but did not directly extend or depend on it, which meant we were not able to incorporate ongoing package and feature improvements, such as ingesting weekly rather than daily hospital admissions data. Likewise, this choice prevents existing `EpiNow2` users from readily accessing our wastewater extensions.

#### Model limitations

Model limitations include:

- The model does not account for the inter-individual variability in shedding rates. This biological variability would be expected to impact the level of noise in a particular site as a function of the number of contributing infected individuals in that site. See EpiSewer (28) for an example of how to model this variability explicitly.

- The model does not account for changes in lab processing methods within a lab that might impact the magnitude of observation error of the resulting measurements. For example, if a lab switched from qPCR to digital droplet PCR, our current model would be unaware of this change during the calibration period and would still estimate a single site-lab level multiplier $M_{ij}$ and a site-lab level observation error $\sigma_{cij}$.

- In the model, the only way for consistent trends in wastewater concentrations to arise is via a corresponding trend in subpopulation-level infections. For example, if an external process such as increased rainwater leads to a decline in wastewater concentrations in a particular site, the model will interpret that decline as evidence of a decline in subpopulation-level infections.

- The model does not account for incomplete reporting of hospital admissions via a reporting delay model, such as those that `EpiNow2` (1) and `epinowcast` (29) provide. This means that even if reported hospital admissions on a particular day are right-truncated (incomplete as of the date they were reported), the model will interpret the reported as the final total reported number of admissions; this can result in forecasts that underpredict final observed hospital admissions.

- The model cannot accept as an input wastewater concentrations measured in units of viral genomes per unit mass of dry sludge, but only concentrations measured in units of genomes per unit volume of liquid.

- The model does not explicitly account for a number of components of the wastewater data generating process that can affect the measurements reported. These include the mechanics of the sewer network, the sample collection type, the extraction method, the concentration method, and the quantification method. All of these processes may affect the measurement of the level of virus in a sewage plant systematically, and our mode relies on the assumption that these processes will on average remain the same during the calibration period within a particular site and a lab.

- The model does not account for changing population sizes. For example, in towns with populations that are highly transient such as seasonal destinations, the population contributing to the wastewater can vary significantly across a matter of weeks. Currently, the model requires passing in a single population size to represent the population of each wastewater catchment area.

- The model does not currently leverage any data on flow rate to incorporate flow normalized observations, nor does it use any indicators of human contributions to wastewater such as pepper mottle virus indicators as a way of normalizing the reported concentrations.

### Additional figures and tables

Below, we provide a table which describes the differences in the approach to forecast production between the real-time and retrospective forecasts.

| | **Real-time** | **Retrospective** |
|---|---|---|
| **Manual review and exclusions of hospital admissions data points** | yes | no |
| **Manual review of forecasts produced from wastewater-informed model** | yes | no |
| **Vintaged wastewater data used (data available as of the forecast date)** | yes | yes |
| **Vintaged hospital admissions data used (data available as of the forecast date)** | yes | yes |
| Forecast date range | February 5th, 2024 – March 25th, 2024 | October 10th, 2023- March 25th, 2024 |

Table S1. Overview of the differences between the real-time and retrospective approaches to forecast production.

| **Model name** | **Description** |
|---|---|
| `cfa-wwrenewal(real-time)` | The model CFA submitted in real-time to the COVID-19 forecast Hub as `cfa-wwrenewal`. Contains a mix of |

| | |
|---|---|
| | wastewater-informed forecasts and hospital admissions only forecasts (with hospital admissions only forecasts submitted when wastewater missing or wastewater-informed forecasts failed to converge/not deemed plausible by visual inspection). |
| `cfa-hosponlyrenewal(real-time*)` | The model CFA produced in real-time and saved locally, containing only hospital admissions-informed forecasts. This model was not submitted to the Hub. |
| `cfa-wwrenewal(retro)` | The model used to retrospectively produce wastewater-informed forecasts for the 2023-2024 winter epidemic wave. |
| `cfa-hosponlyrenewal(retro)` | The model used to retrospectively produce hospital admissions-only forecasts for the 2023-2024 epidemic wave. |
| `COVIDhub-4_week_ensemble` | An ensemble, or model average, of submitted forecasts to the COVID-19 Forecast Hub |
| `COVIDhub-baseline` | For predictions of incidence, the median prediction at all future horizons is the most recent observed incidence. |
| `UMass-sarix` | Equally weighted ensemble of simple time-series baseline models |
| `CMU-TimeSeries` | An ensemble of (a) 28-day-window AR + residual resampling, (b) direction-stratified quantile AR (QAR), and (c) windowed QARX with cases. |
| `UMass-gbq` | Bagged gradient boosting is used to obtain a predictive median. Features include the day of week, location, |

| | |
|---|---|
| | population, and summaries of timeseries behavior. Quantiles are obtained via a procedure similar to split conformal prediction, where quantiles of the out-of-bag residuals are added to the point predictions obtained from gradient boosting. |
| `MUNI-ARIMA` | This is an ARIMA model with outlier detection fitted to transformed weekly aggregated series. |
| `UMass-trends_ensemble` | Equally weighted ensemble of simple time-series baseline models. Each baseline model calculates first differences of incidence in recent weeks. These differences are sampled and then added to the most recently observed incidence. Variations on this method include (a) including the first differences and the negative of these differences to enforce symmetry, resulting in a flat-line forecast, (b) varying the number of time-units in the past for computing the first differences (typically 14, 21, or 28 days) to focus on capturing recent trends, and (c) using the original time-series or a variance-stabilizing transformation of it, e.g. square-root. Additionally, the resulting predictive distributions are truncated so that any predicted samples computed to be less than zero are truncated to be zero. |
| `SGroup-RandomForest` | Random Forest ensemble of the predictors generated from the USC-SIkJalpha submission as well as HHS data. |
| `UT-Osiris` | A semi-mechanistic model that combines aspects of mechanistic and statistical models. |

| | |
|---|---|
| `CEPH-Rtrend_covid` | Renewal equation based on R(t) estimation from hospitalization data. |

Table S2. Description of the COVID-19 forecasting models used in this manuscript. The first four rows describe the different variations of the wastewater-informed and hospital admissions only models that are the focus of this study, the remaining rows describe models submitted in real-time to the COVID-19 Forecast Hub. Descriptions of those models are taken from the COVID-19 Forecast Hub model metadata.

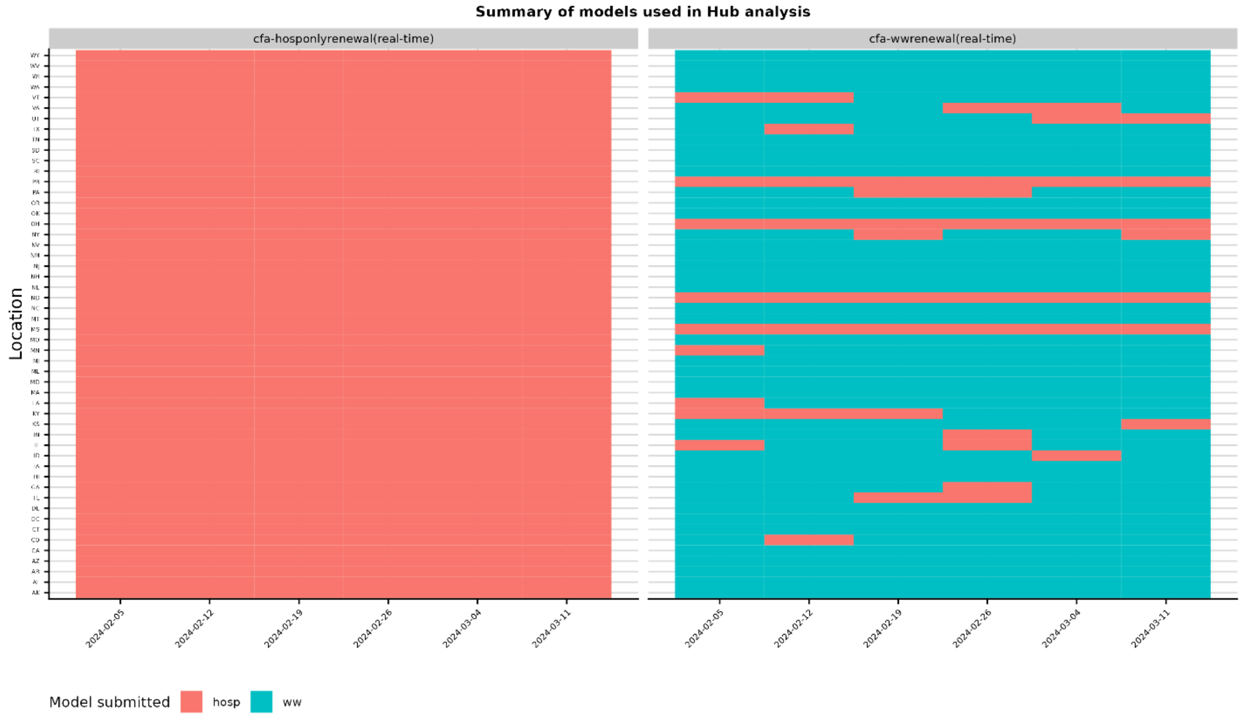


Fig. S1 Heatmap indicating which model was used in the real-time analysis for the Hub model comparison. The hospital admissions-only model was submitted for any location/forecast date without wastewater, when the wastewater-informed model failed to converge, or when the wastewater-informed model results were deemed implausible in manual review. For the real-time relative performance analysis (wastewater-informed versus admission-only), the forecast dates/locations for which the hospital admissions-only model was submitted were excluded.

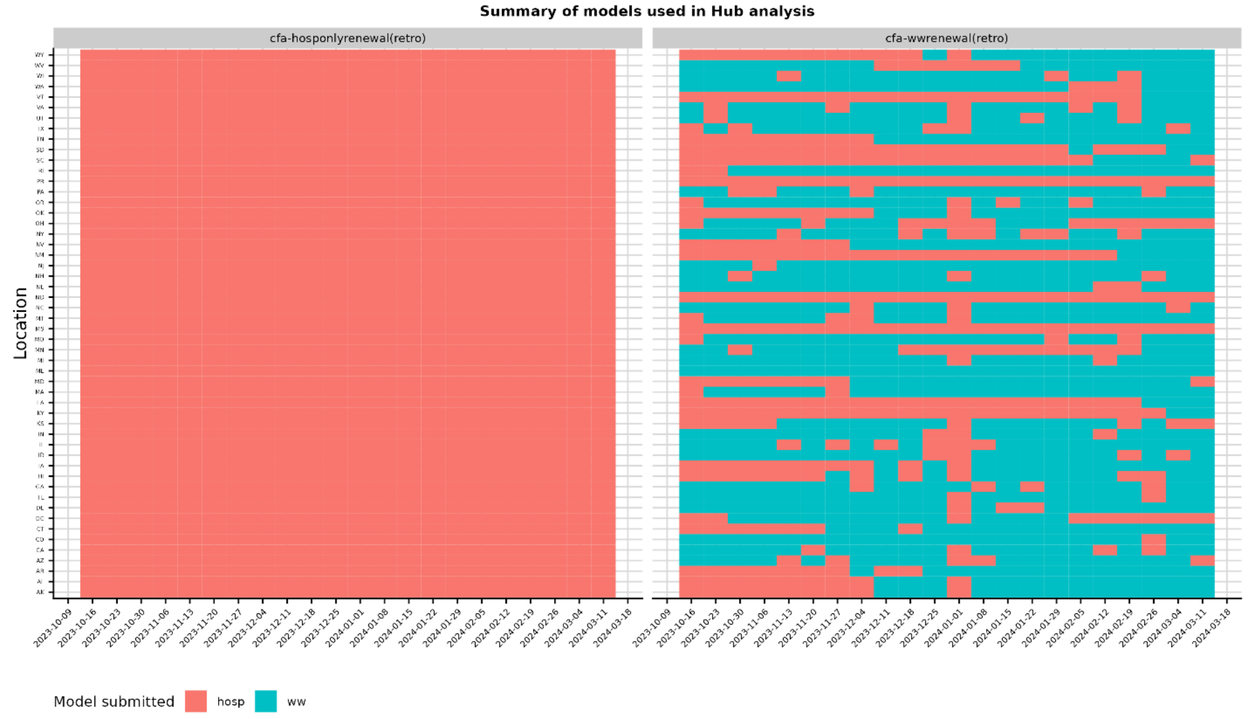


Fig S2. Heatmap indicating which model was used in the retrospective analysis of performance compared to the other Hub models. The hospital admissions-only model was used for a location/forecast date if there was insufficient wastewater or the wastewater-informed model had convergence issues.

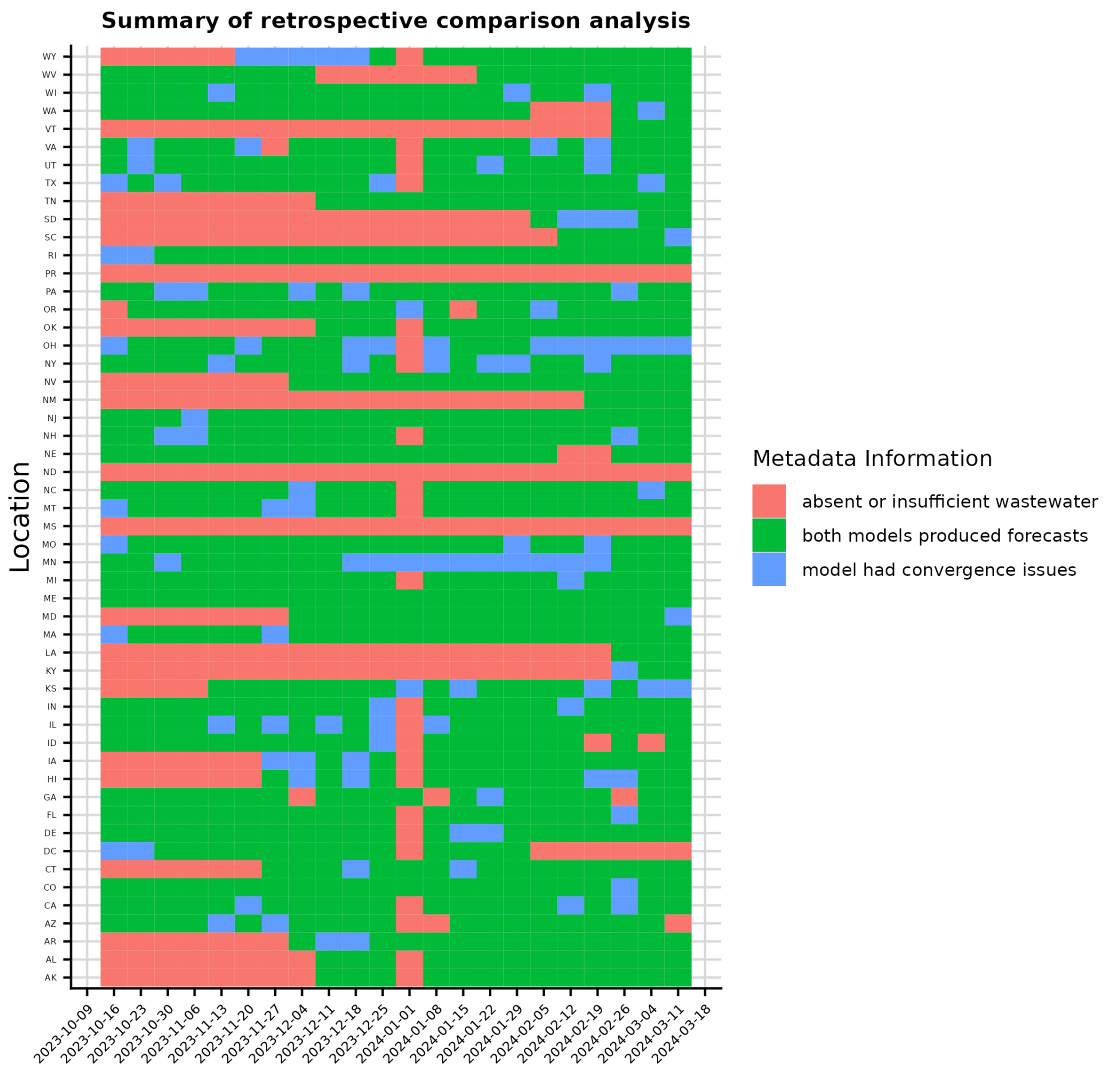


Fig. S3 Heatmap indicating the forecast dates and locations evaluated in the retrospective comparison of the wastewater-informed model and the hospital admissions-only model (green), and if not evaluated, the reason for being excluded (red: absent or insufficient wastewater data; blue: model convergence issues).

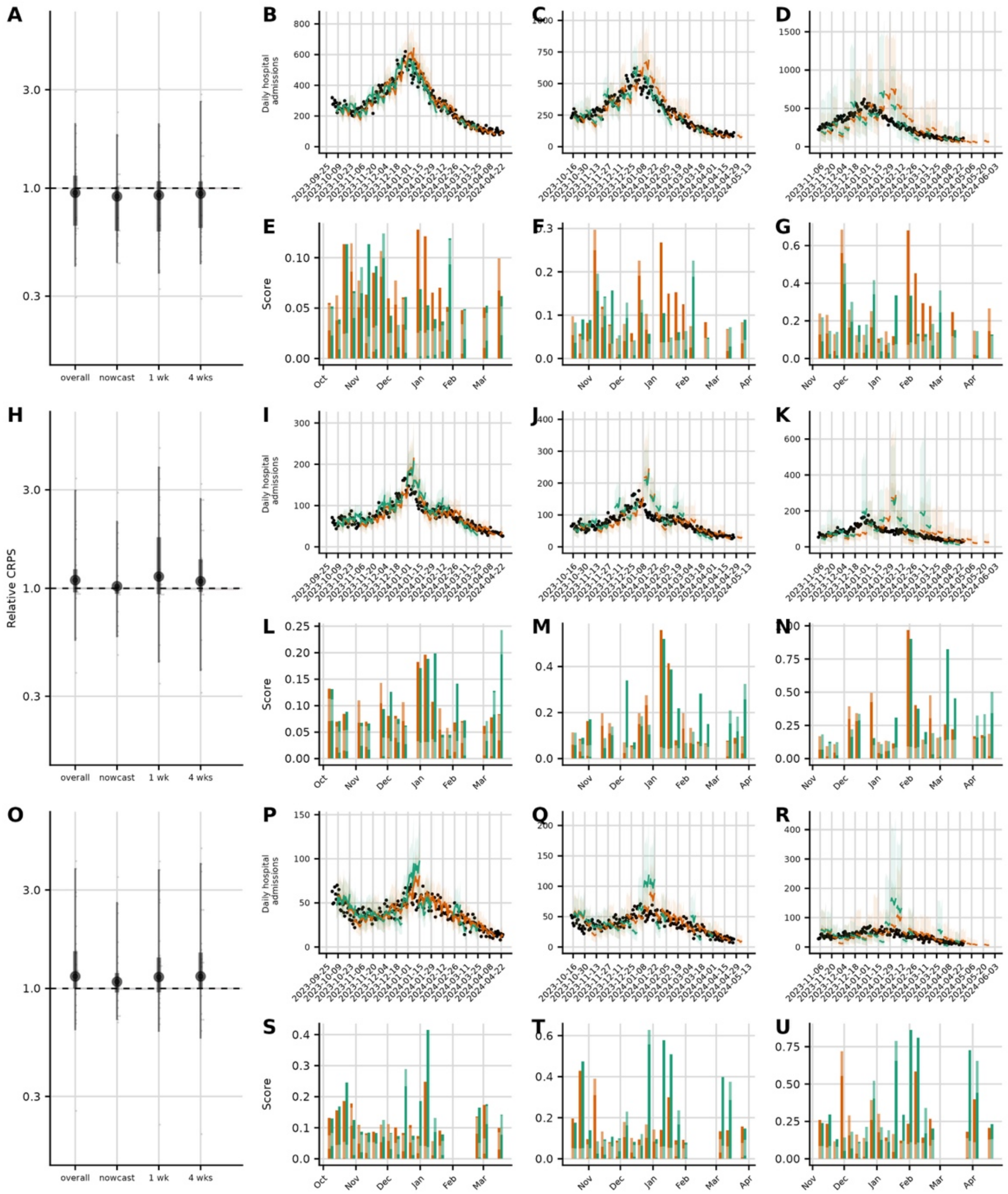


Fig S4. Example of retrospective forecast performance for three jurisdictions across the 2023-24 epidemic season. Relative continuous ranked probability score (rCRPS) is computed by first calculating the mean CRPS for the wastewater-informed model across horizon days and then then computing the ratio of the wastewater-informed model mean CRPS to the hospital admissions-only model mean CRPS. A rCRPS below 1 indicates improved performance; a relative CRPS above 1 indicates reduced performance. Rows correspond to California, Virginia, and Washington respectively (top to bottom). A, D, G. Distribution of relative CRPS for individual forecast dates, stratified by horizon (overall, nowcast, one week, and four weeks). Black points indicate the geometric mean of the individual relative CRPSes; thick bars indicate the central 68% interval, thin bars indicate the central 95% interval. B, E, H. Nowcasts and forecasts of hospital admissions each by forecast horizon (nowcast, one week, and four weeks) for the wastewater-informed model (green) and the hospital admissions only model (orange) with closed black circles indicating the final observed hospital admissions used for evaluation. C, F, I. Absolute CRPS averaged across all days within that horizon for each forecast date, colored by model. Gaps indicate forecast dates where either the wastewater data was insufficient or the model failed to converge.

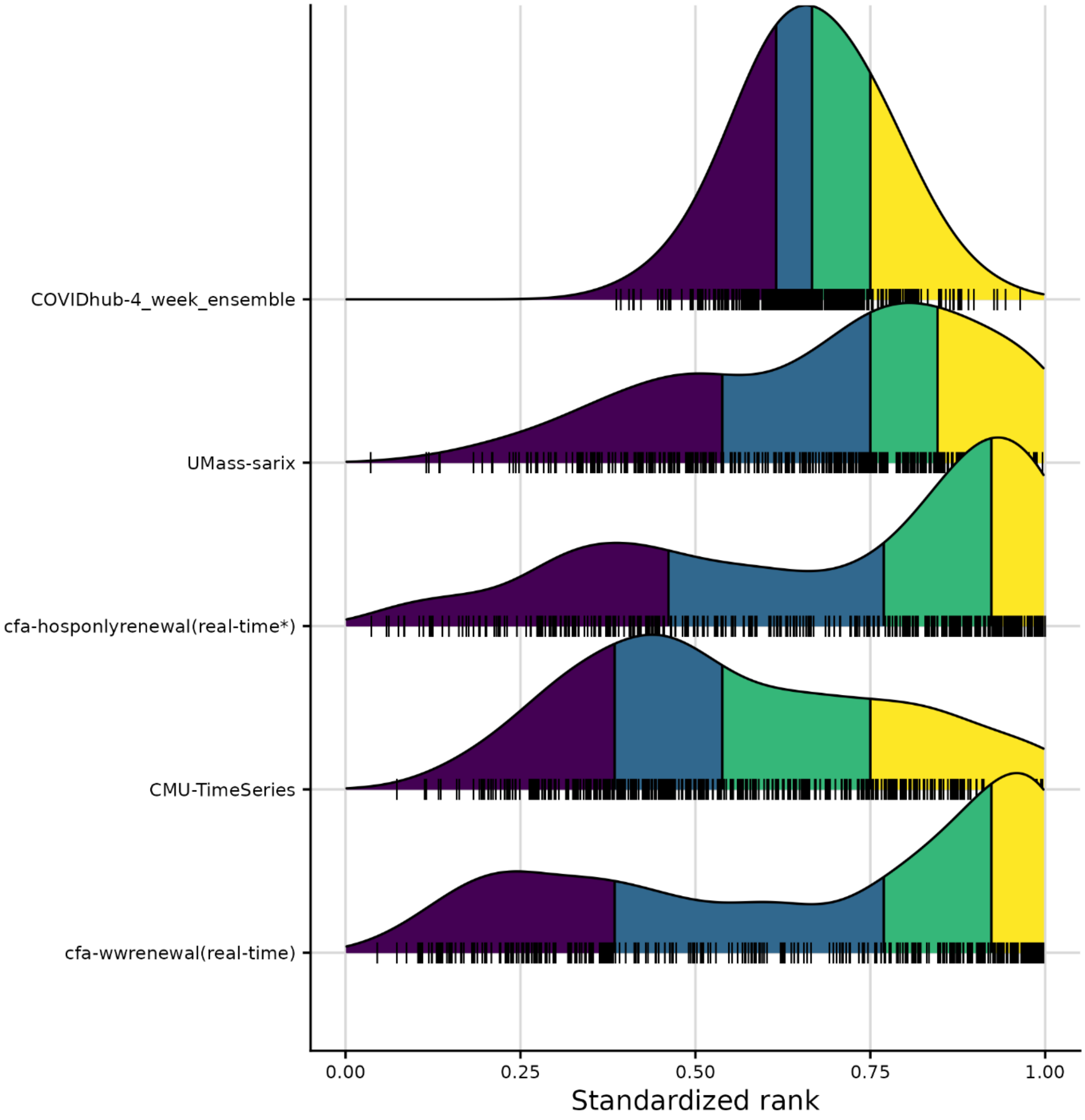

Fig. S5 Distribution of model standardized rank by WIS for each jurisdiction/forecast date evaluated for the real-time forecast evaluation. A standardized rank of 1 indicates that the model had the best WIS for that particular jurisdiction/forecast date, and the value closest to 0 indicates it had the worst WIS. For example, if there were 5 forecasts submitted for a location and forecast date, the one with the highest WIS (worst performing forecast) would have a standardized rank of 0.2 (1/5), while the second best would have a standardized rank of 0.8 (4/5). The density plots show interpolated distributions of the standardized ranks achieved by each model for every forecast date and location that the model forecasted. The quartiles of each model's distribution of standardized ranks are shown in different colors: yellow indicates the top quartile of the distribution and purple indicates the bottom quarter of the distribution. The models are ordered by the last quartile of the distribution, with models that rarely had a low rank near the top.

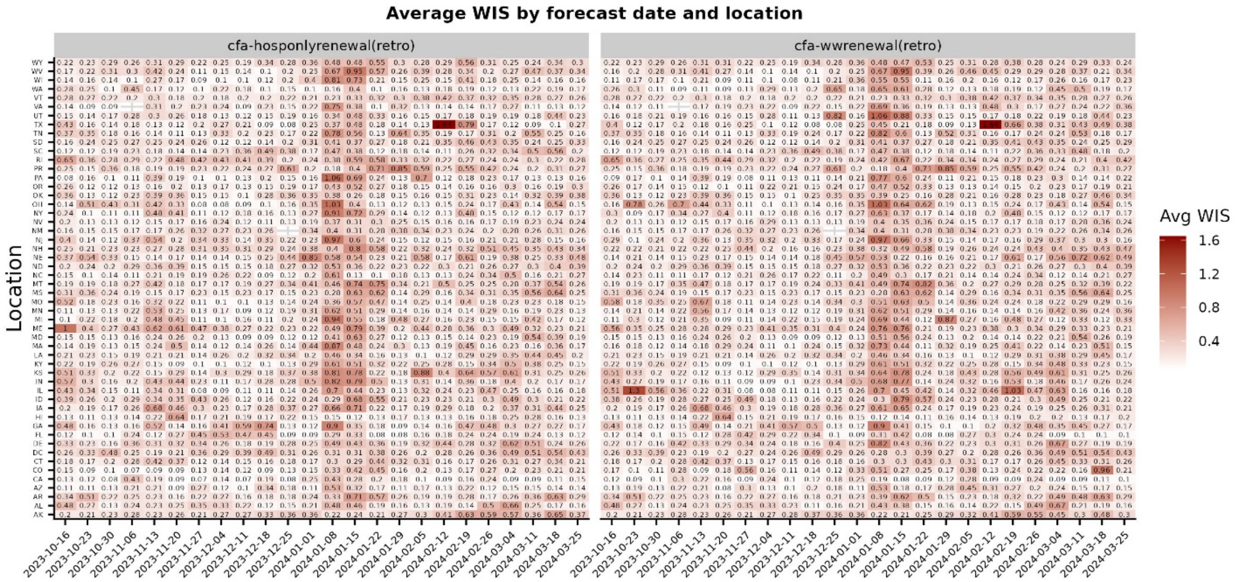


Fig. S6 Heatmap of average WIS by forecast date and location for the retrospective wastewater-informed model (*cfa-wwrenewal(retro)*) and the retrospective hospital admissions model (*cfa-hosponlyrenewal(retro)*). Fill indicates the WIS averaged across the 28 horizon days in that particular location on that forecast date.

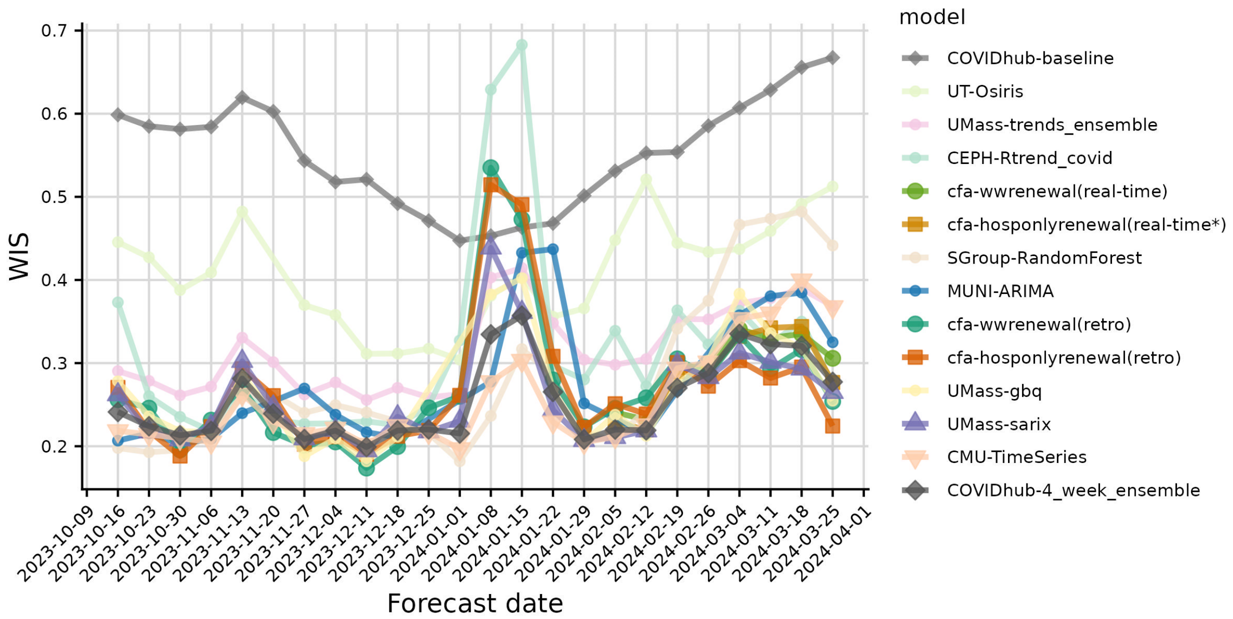


Fig. S7 Average WIS over time across jurisdiction for each model submitted to the COVID-19 Forecast Hub plus the retrospective submissions from the retrospective wastewater-informed model `cfa-wwrenewal(retro)` and the hospital admissions-only model (`cfa-hosponlyrenewal(retro)`).

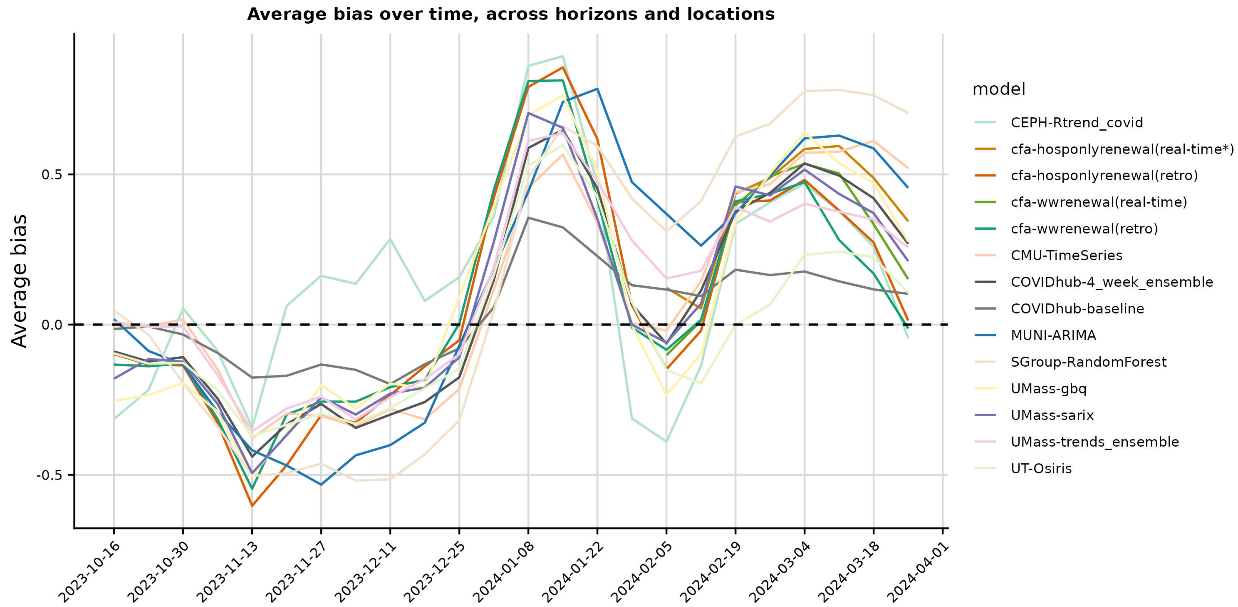


Fig. S8 Bias over time averaged across jurisdiction for each model submitted to the COVID-19 Forecast Hub plus the retrospective submissions from the retrospective wastewater-informed model `cfa-wwrenewal(retro)` and the hospital admissions-only model (`cfa-hosponlyrenewal(retro)`).

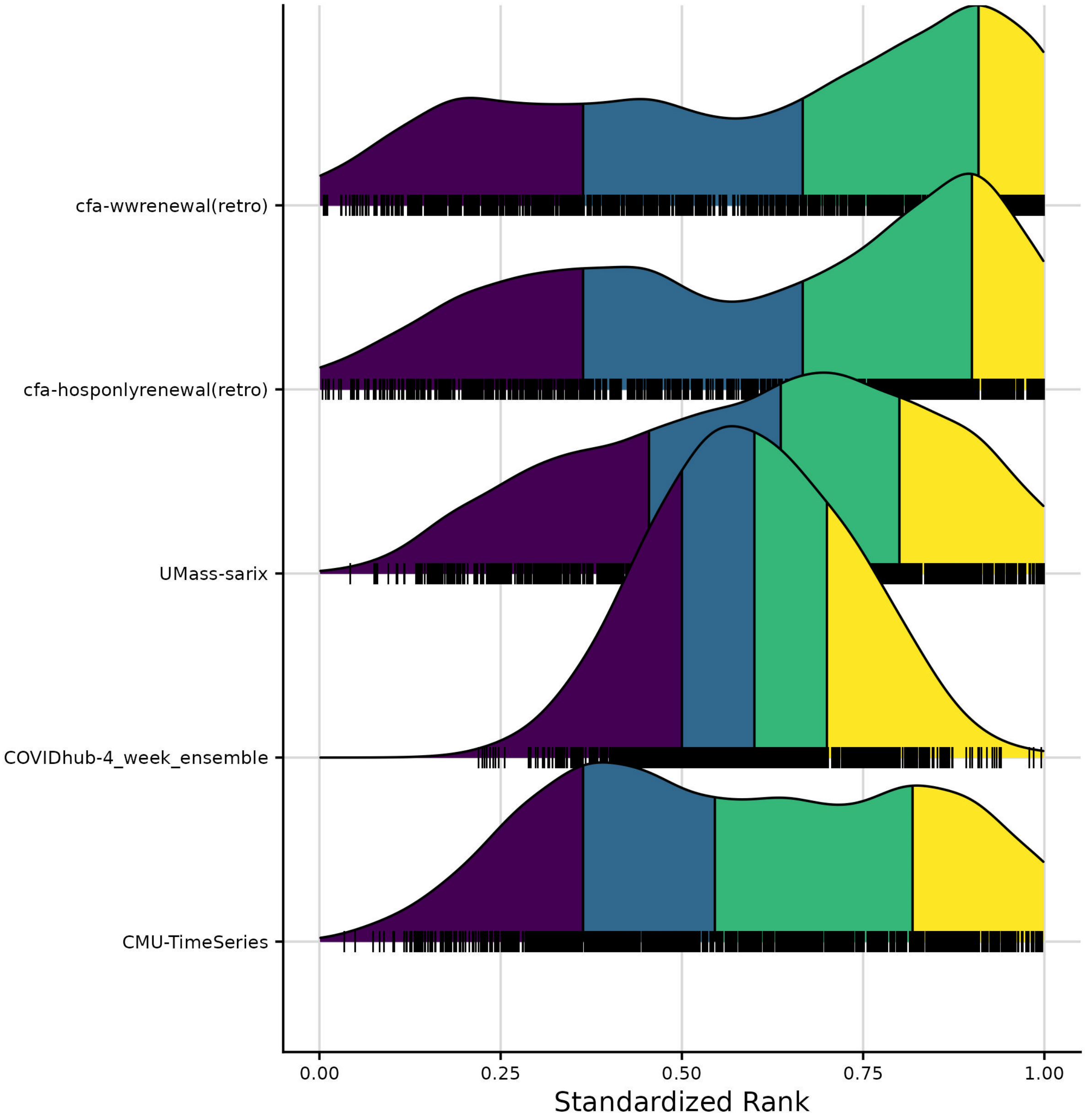


Fig. S9 Distribution of model standardized rank by WIS for each jurisdiction/forecast date evaluated for the retrospective forecast evaluation. A standardized rank of 1 indicates that the model had the best WIS for that particular jurisdiction/forecast date, and the value closest to 0 indicates it had the worst WIS. For example, if there were 5 forecasts submitted for a location and forecast date, the one with the highest WIS (worst performing forecast) would have a standardized rank of 0.2 (1/5), while the second best would have a standardized rank of 0.8 (4/5). The density plots show interpolated distributions of the standardized ranks achieved by each model for every forecast date and location that the model forecasted. The quartiles of each model's distribution of standardized ranks are shown in different colors: yellow indicates the top quartile of the distribution and purple indicates the bottom quarter of the distribution. The models are ordered by the last quartile of the distribution, with models that rarely had a low rank near the top.

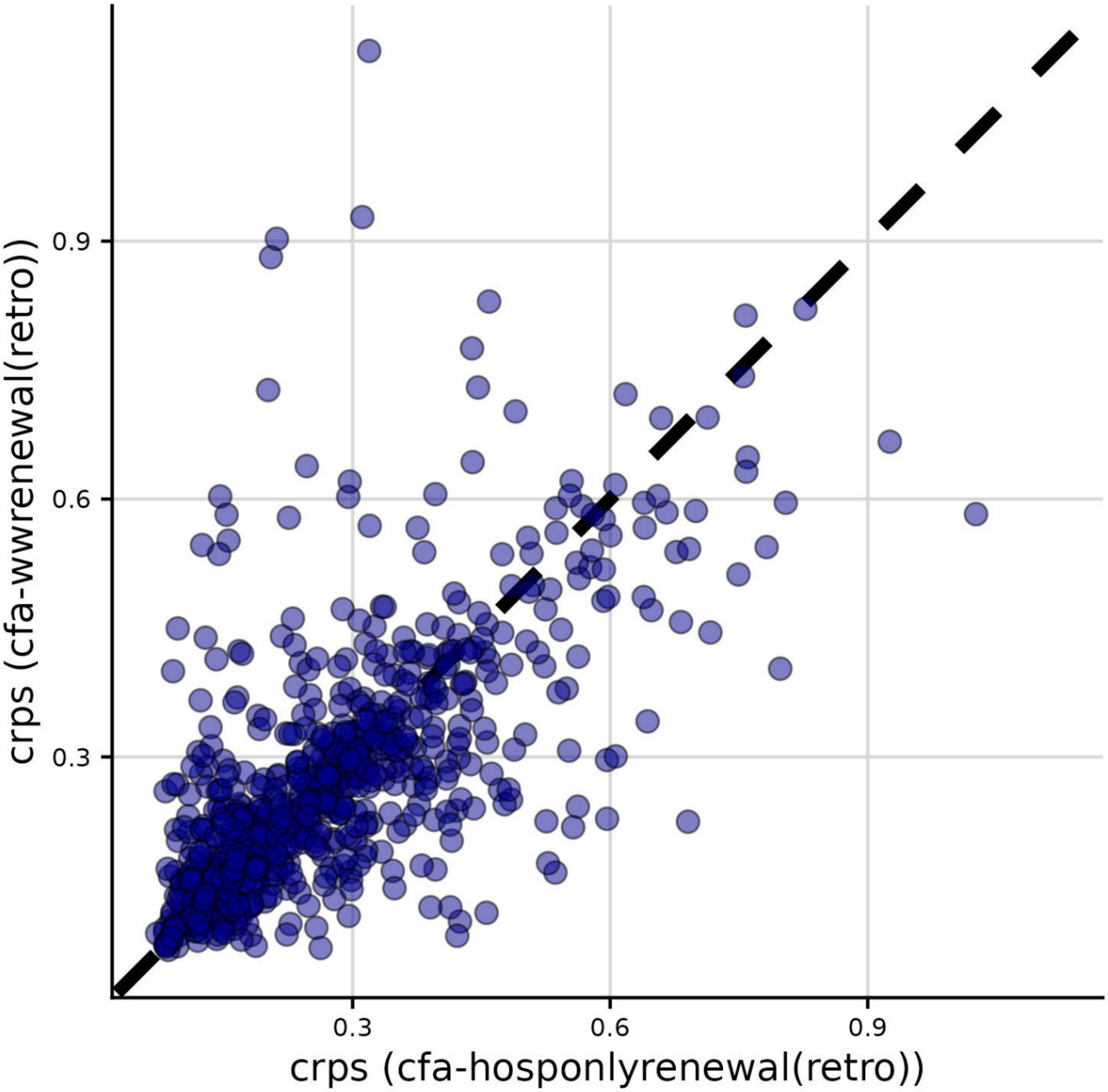


Fig, S10 CRPS of the retrospective wastewater-informed model vs. CRPS of the retrospective hospital admissions-only model, averaged across horizon days, for each forecast date and location.

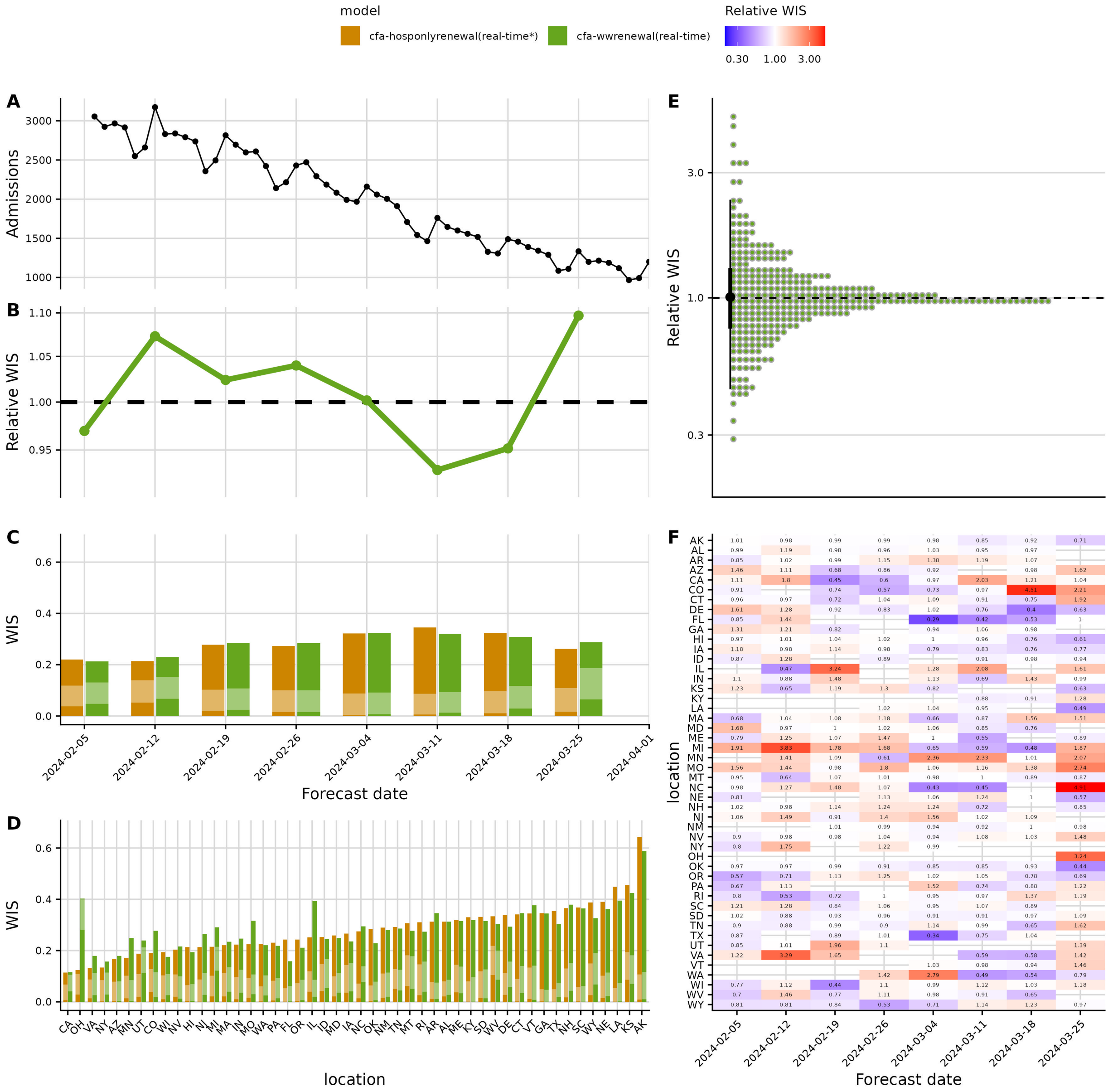


Fig. S11 Real-time forecast performance with and without wastewater data from Feb-Mar 2024. Relative weighted interval score (rWIS) is computed by first averaging across the 28 horizon days and then computing the ratio of wastewater-informed mean WIS to the hospital admissions-only model mean WIS. For rWIS, a value less than 1 indicates that the wastewater-informed model outperforms the hospital admissions-only model. A. Heatmap of relative WIS across forecast dates and locations for the 28 day-ahead forecasts. Blue indicates improved performance by rWIS; red indicates reduced performance. Gaps indicate forecasts that were excluded from the analysis because the wastewater-informed model did not make a forecast due to insufficient wastewater data, convergence issues, or real-time manual exclusion based on visual inspection. B. Distribution or relative CRPS for all individual forecast date/jurisdiction pairs. C. Timeseries of observed U.S. national daily hospital admissions during the period evaluated. D. rWIS by forecast date (rWIS is computed relative to the hospital admissions-only model). E, F. WIS by forecast date (E) and jurisdiction (F) for each model, decomposed into penalties for underprediction (bottom solid bar), dispersion (middle transparent bar), and dispersion (top solid bar).

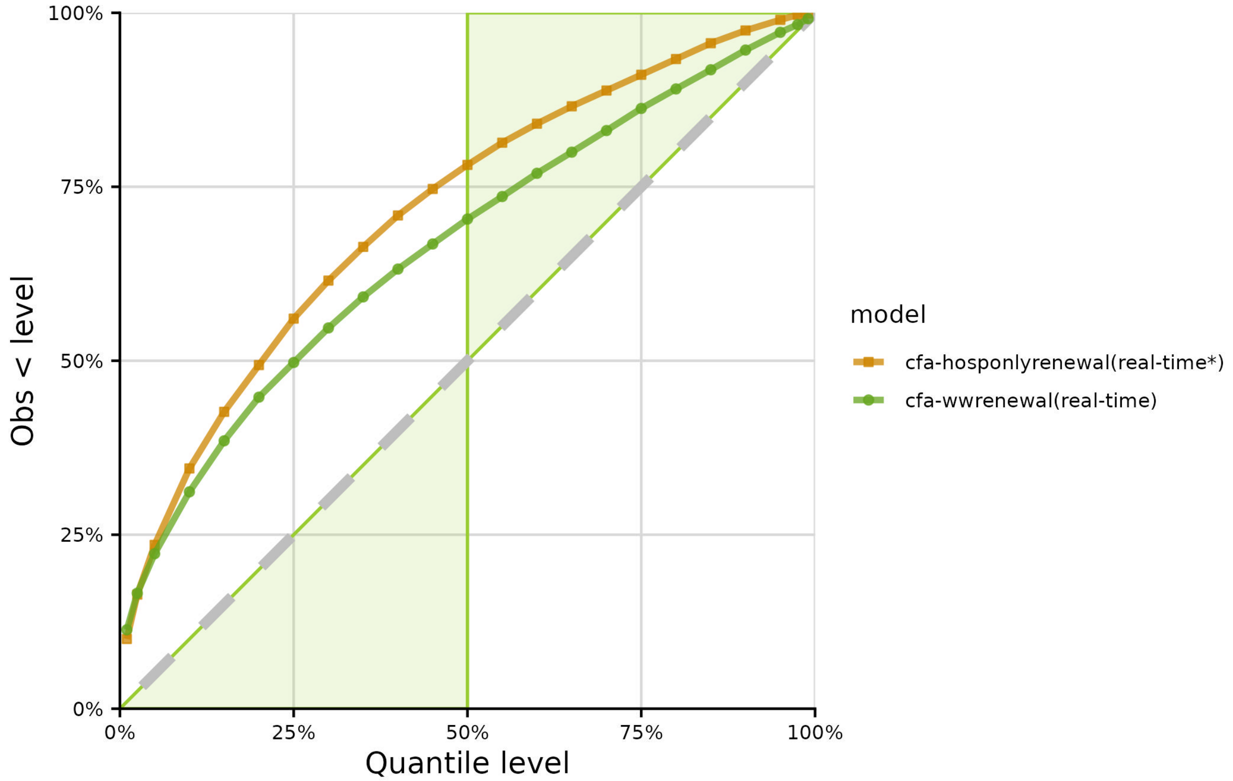


Fig. S12 Empirical quantile coverage for both the real-time wastewater-informed (green circles) and hospital admissions-only (orange squares) forecasts. A well-calibrated forecast should have empirical quantiles that match the theoretical ones (dashed diagonal line). Green shaded area corresponds to conservative forecasts.

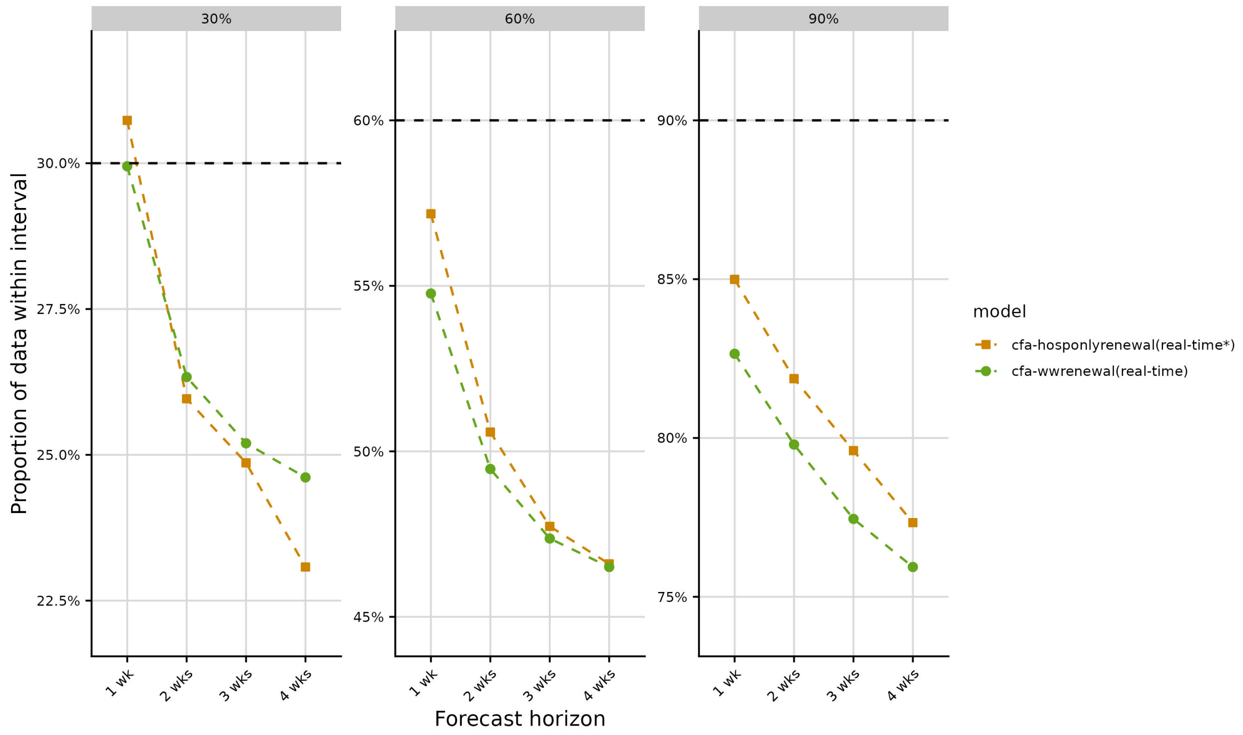


Fig. S13 Empirical interval coverage for the real-time wastewater-informed forecasts (green circles) and the hospital admissions-only forecasts (orange squares) at the 30 percent, 60 percent, and 90 percent prediction intervals, stratified by forecast horizon.

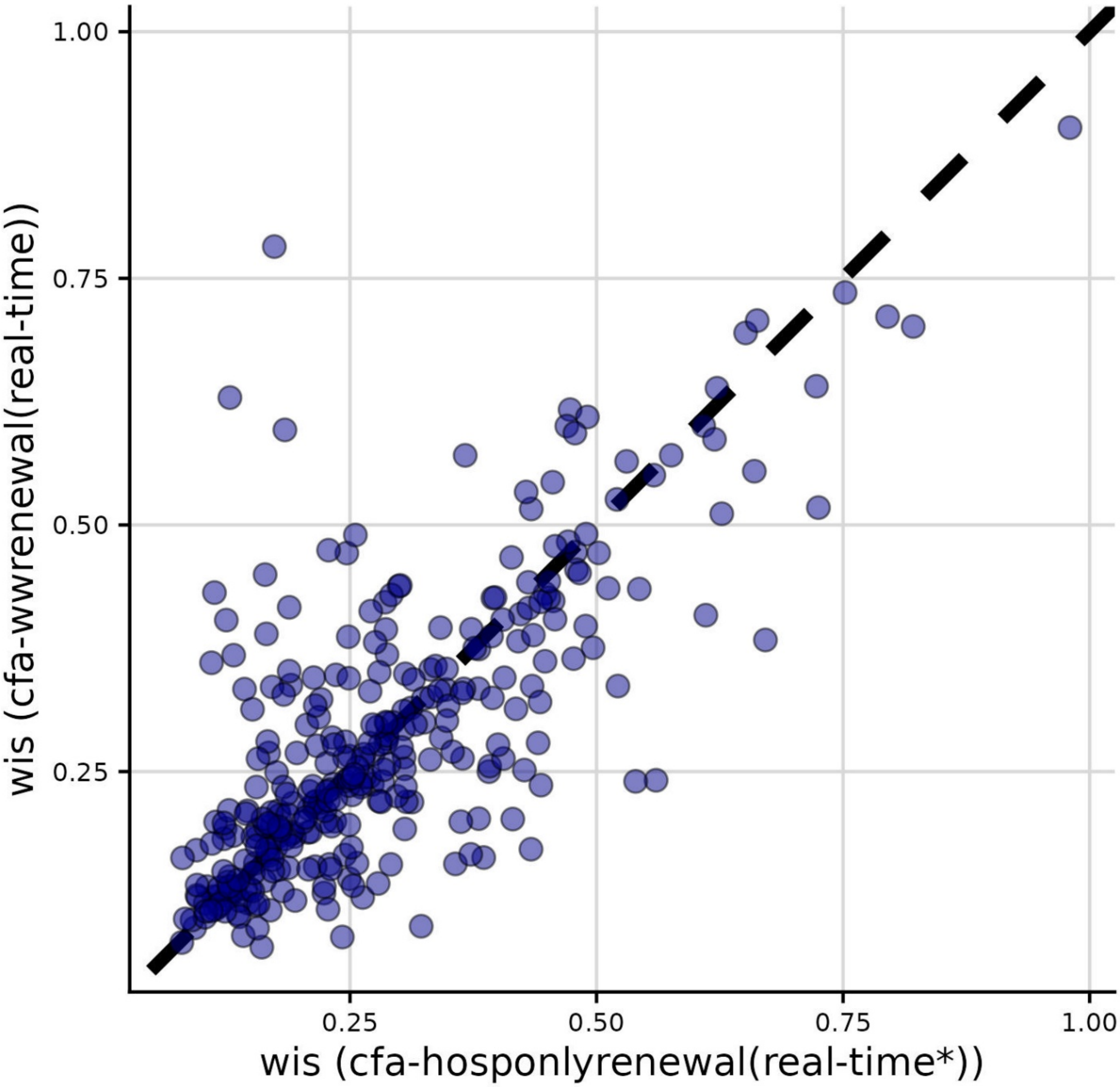


Fig, S14 WIS of the real-time wastewater-informed model vs. WIS of the real-time hospital admissions-only model, averaged across horizon days, for each forecast date and jurisdiction.

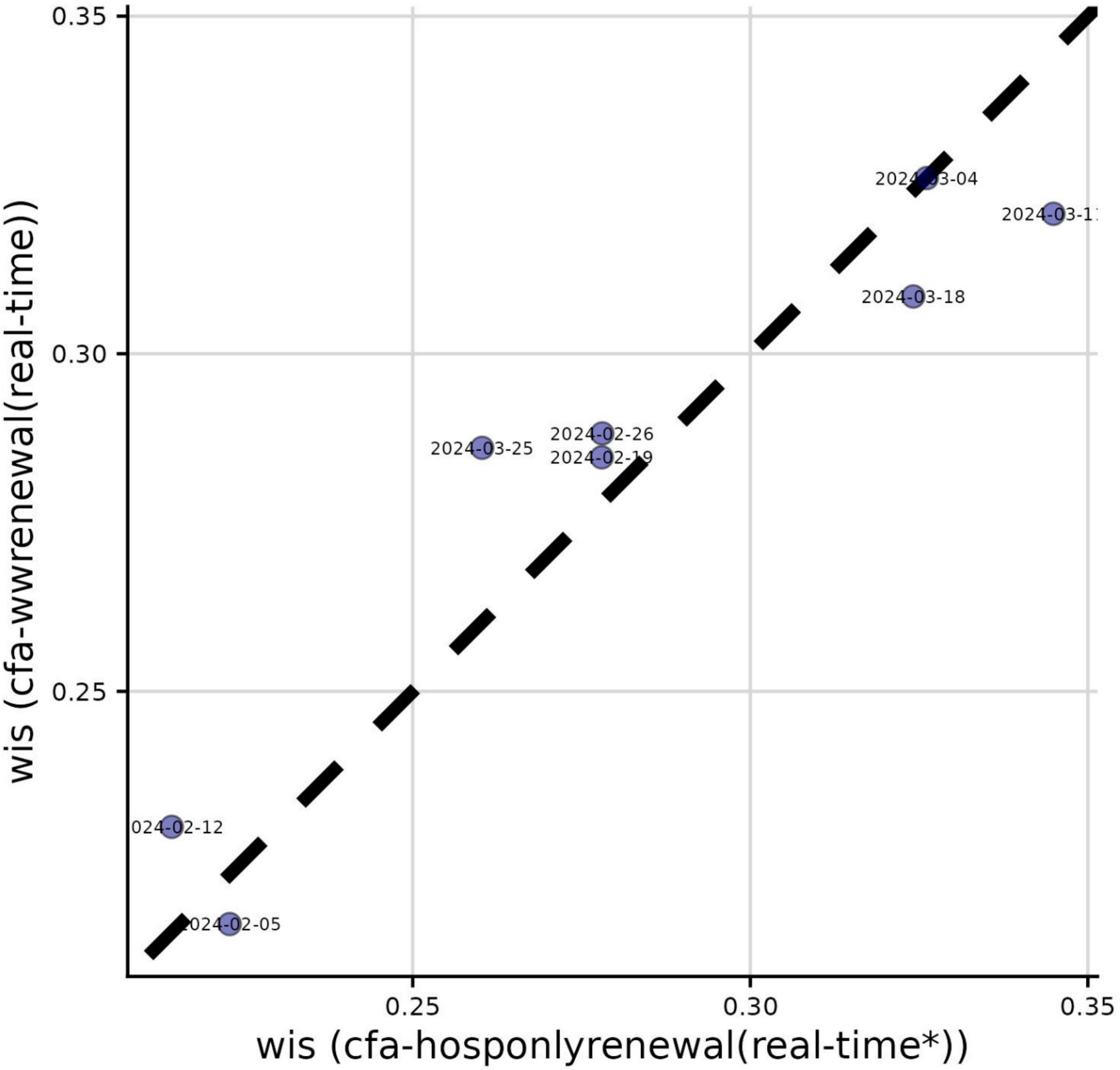


Fig, S15 WIS of the real-time wastewater-informed model vs. WIS of the real-time hospital admissions-only model, averaged across jurisdictions and horizon days, for each forecast date.

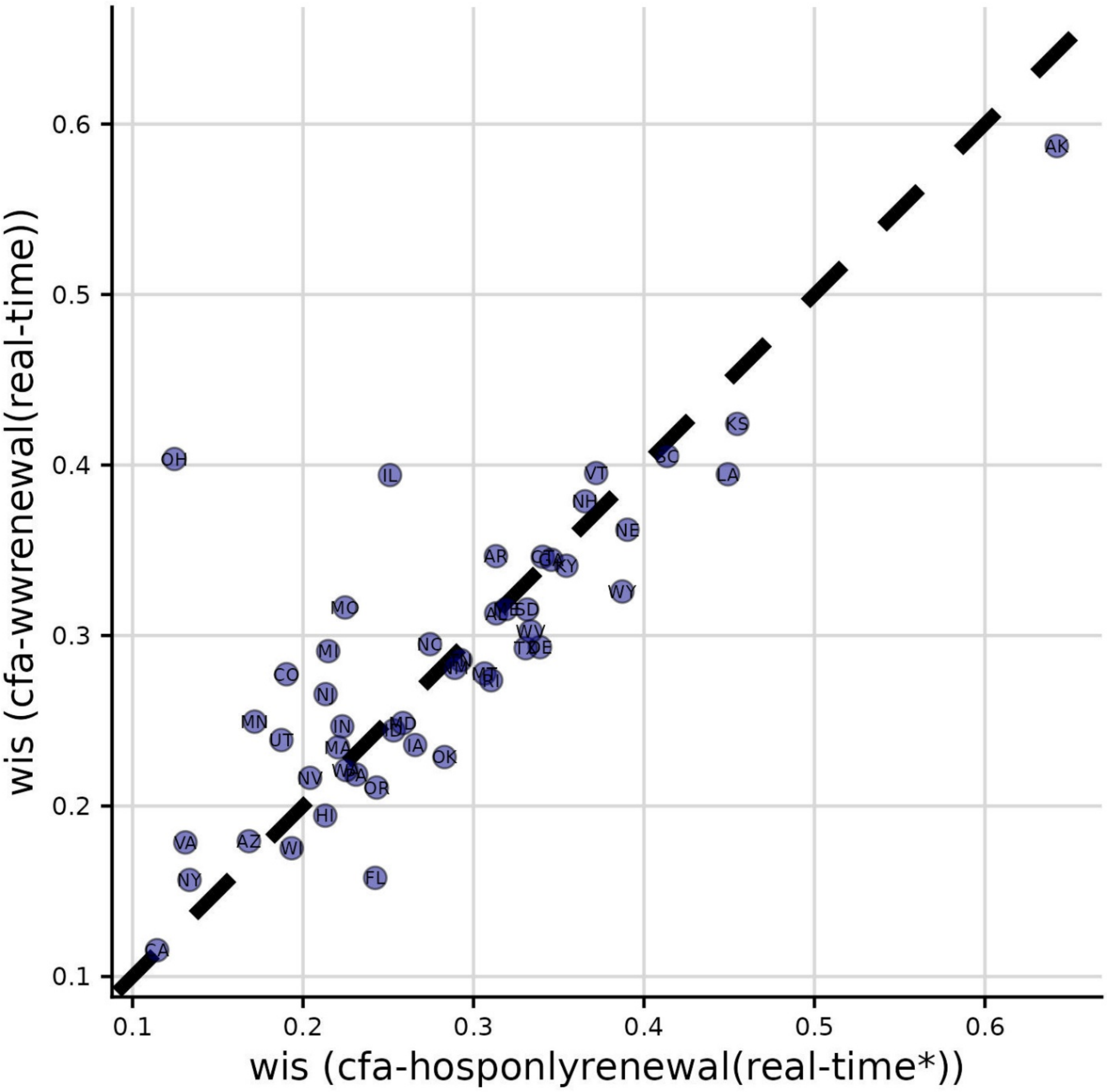


Fig, S16 WIS of the real-time wastewater-informed model vs. WIS of the real-time hospital admissions-only model, averaged across forecast dates and horizon days, for each jurisdiction.

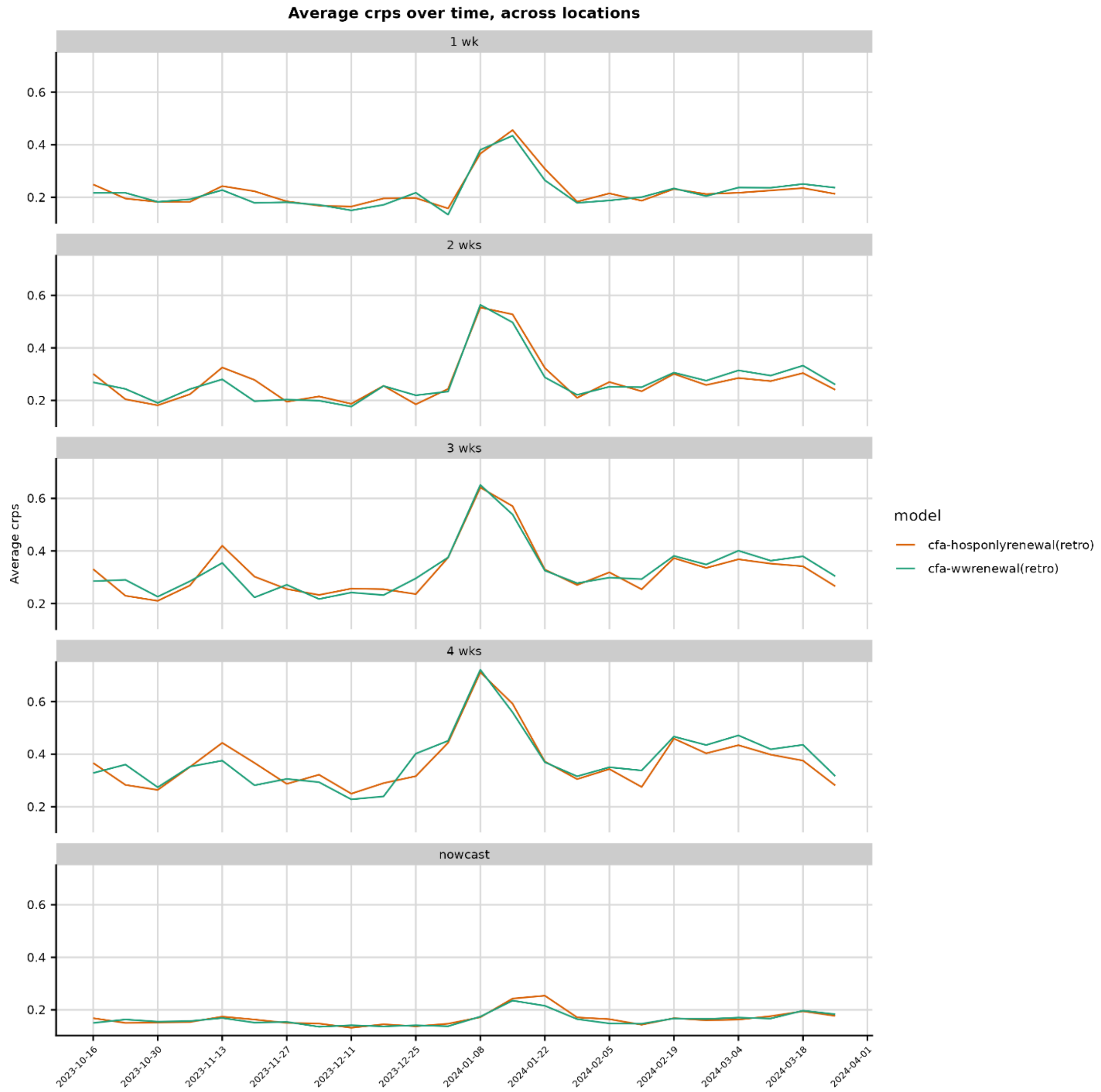


Fig. S17 Average CRPS across jurisdictions for retrospective forecasts on each forecast date, stratified by horizon and colored by model.

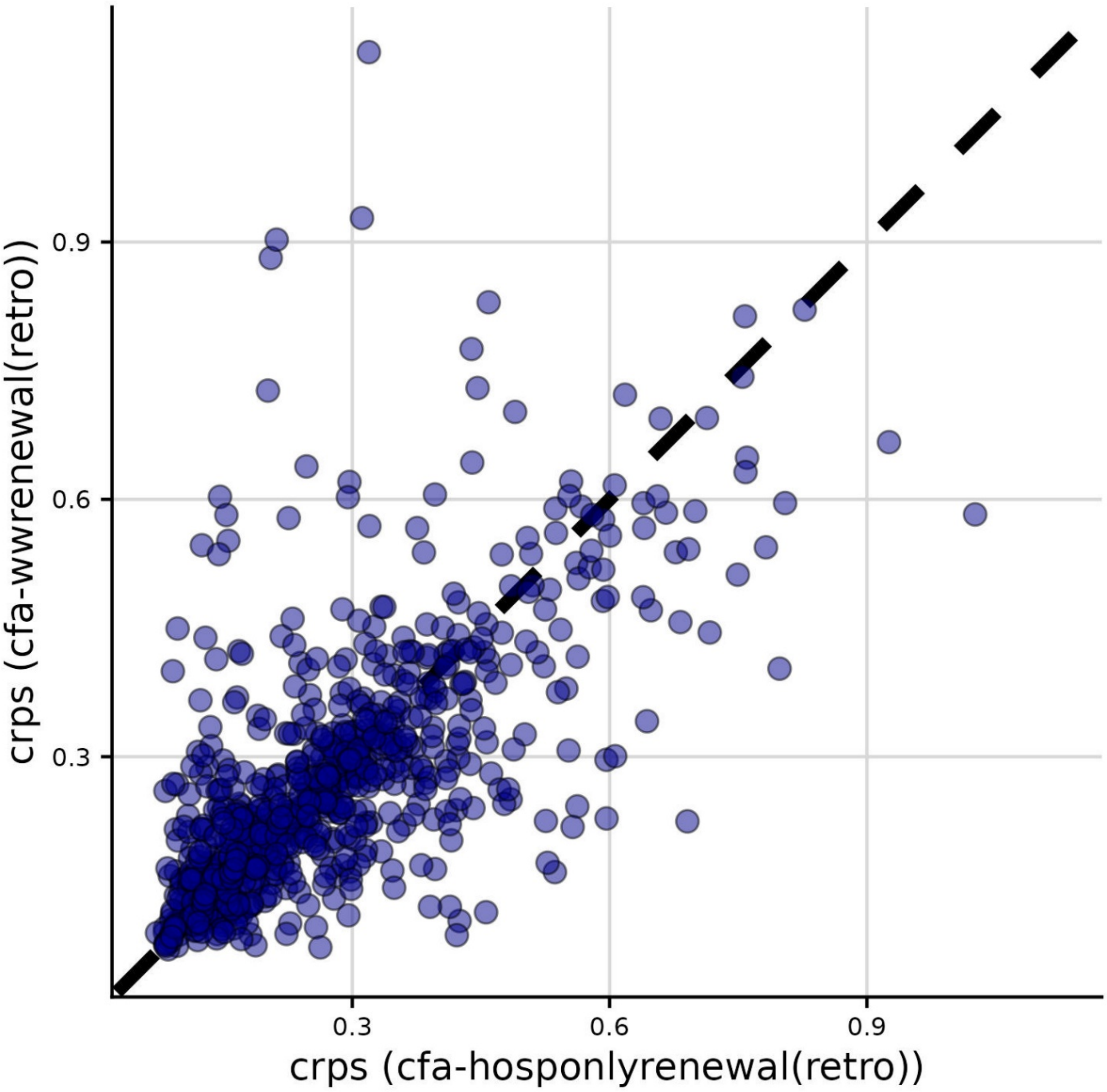


Fig, S18 CRPS of the retrospective wastewater-informed model vs. CRPS of the retrospective hospital admissions-only model, averaged across horizon days, for each forecast date and location.

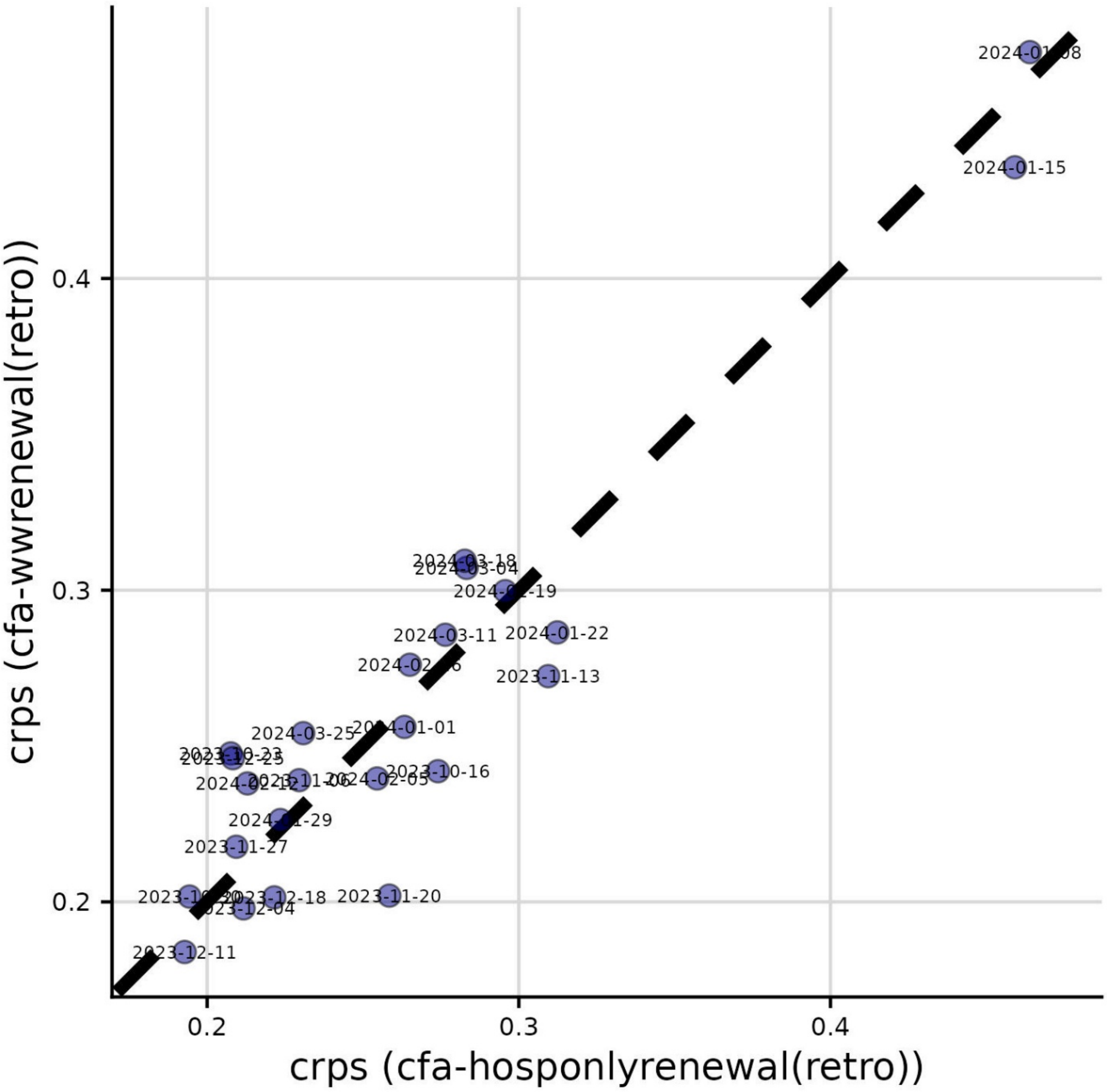


Fig, S19 CRPS of the retrospective wastewater-informed model vs. CRPS of the retrospective hospital admissions-only model, averaged across jurisdictions and horizon days, for each forecast date.

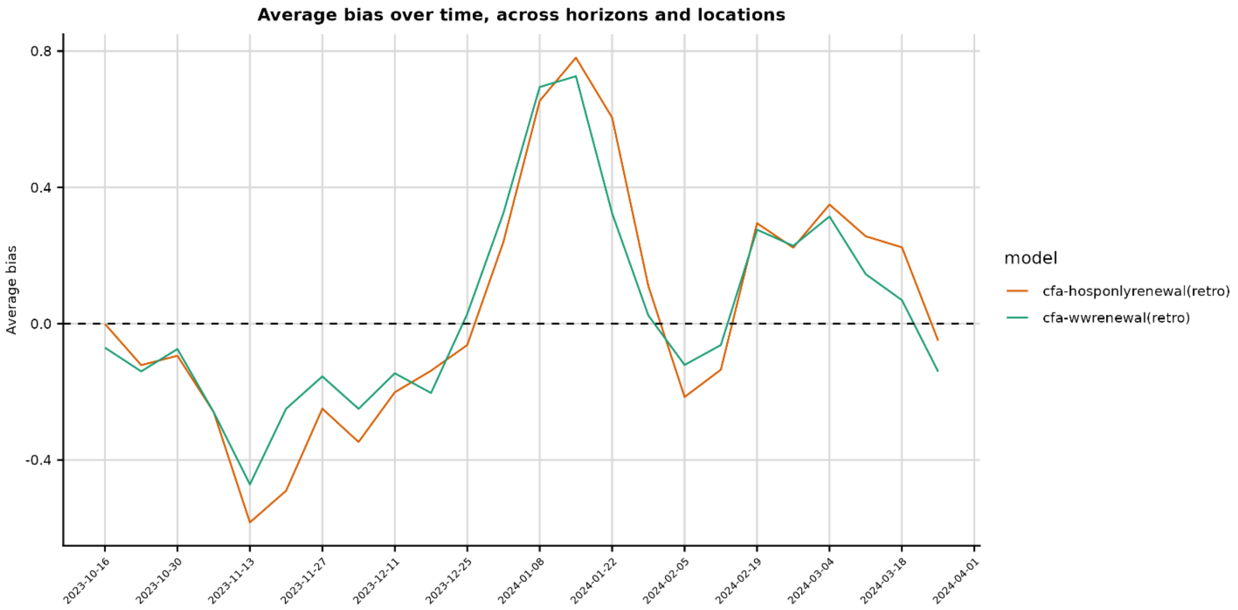


Fig. S20 Average bias of retrospective forecasts over time across jurisdictions and horizon days, stratified by forecast date, colored by model.

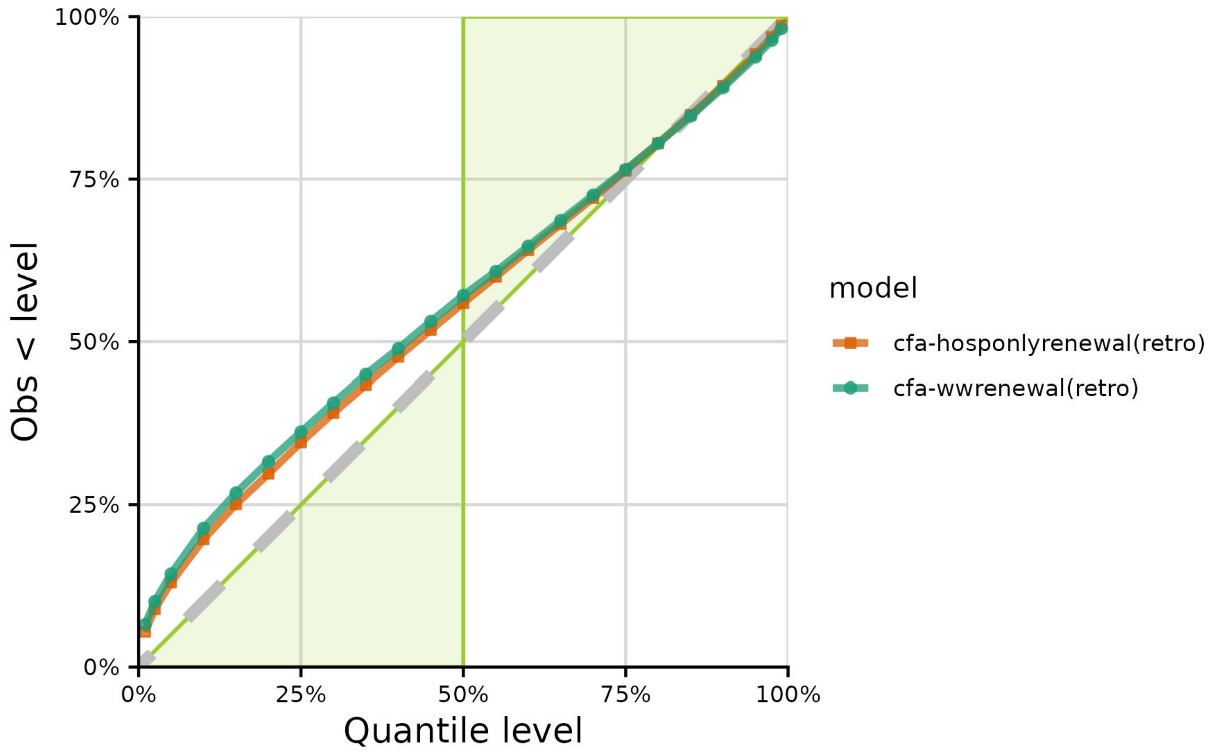


Fig. S21 Empirical quantile coverage for retrospective wastewater-informed (green circles) and hospital admissions-only (orange squares) forecasts. A well-calibrated forecast should have empirical quantiles that match the theoretical ones (dashed diagonal line). Green shaded area corresponds to conservative forecasts.

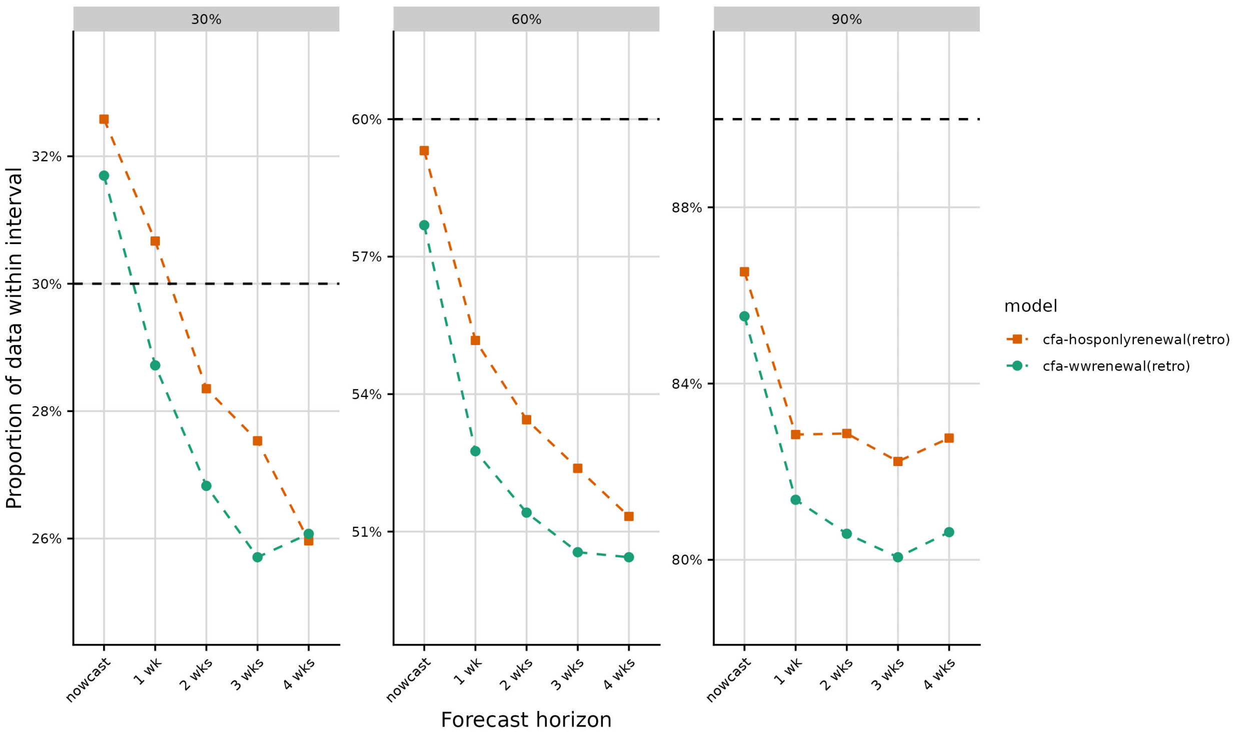


Fig. S22 Empirical coverage of the retrospective wastewater-informed forecasts (green circles) and the hospital admissions-only forecasts (orange squares) at the 30 percent, 60 percent, and 90 percent prediction intervals, stratified by forecast horizon.

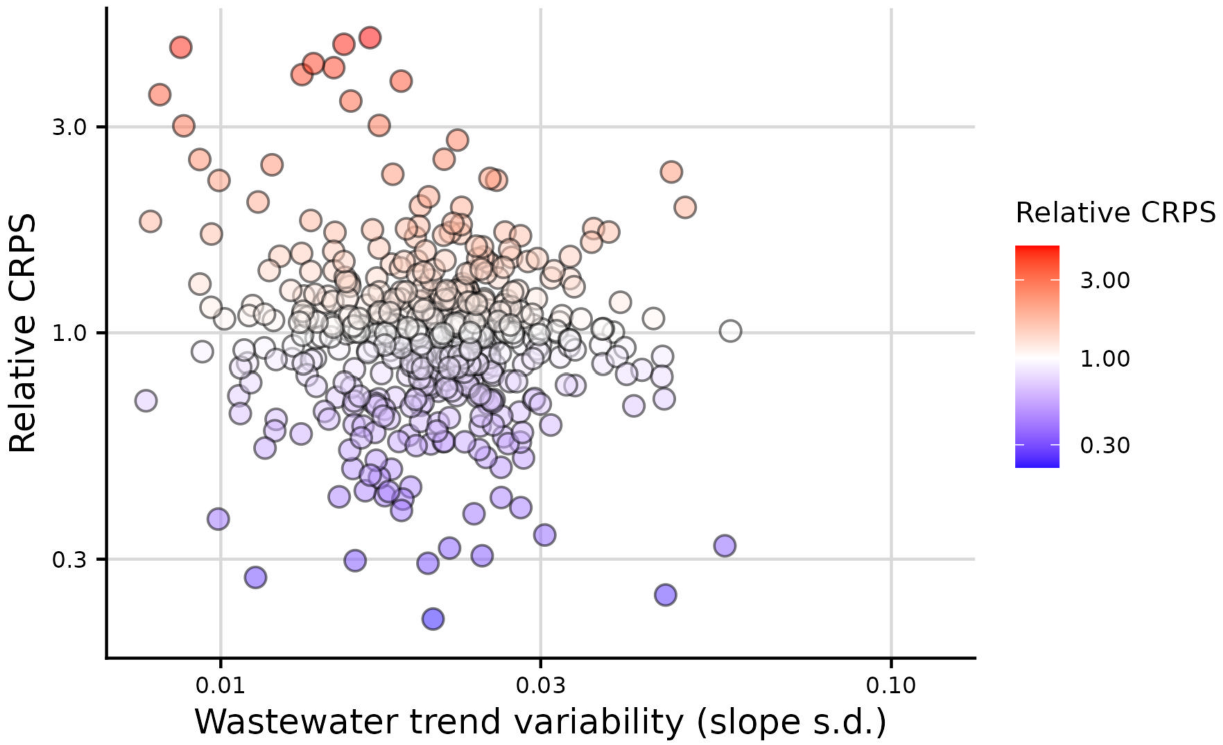


Fig. S23 Relative CRPS of the wastewater-informed model vs the hospital admissions-only model (lower means the wastewater informed model performed better) vs the standard deviation in the slope of the log-linear fit to the wastewater concentrations. Points are posterior median estimates.

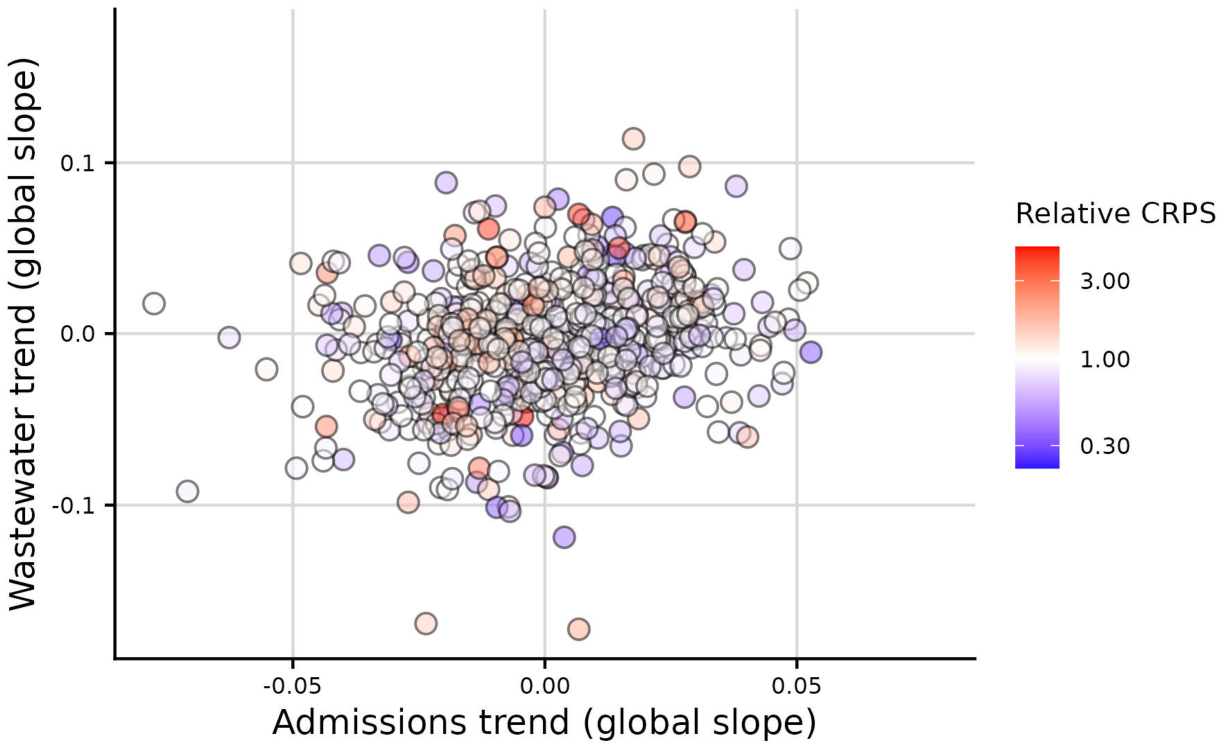


Fig. S24 Global estimate of the log-linear slope in wastewater concentrations across wastewater treatment plants over 23 days prior to the forecast date vs estimate of the log-linear slope in hospital admissions over the last 23 days of observed data, colored by the relative CRPS between the wastewater-informed model relative to the hospital admissions-only model (an rCRPS of 1 indicates equivalent performance, an rCRPS < 1 means the wastewater-informed model performed better than the hospital admissions only model). Points are posterior median estimates.

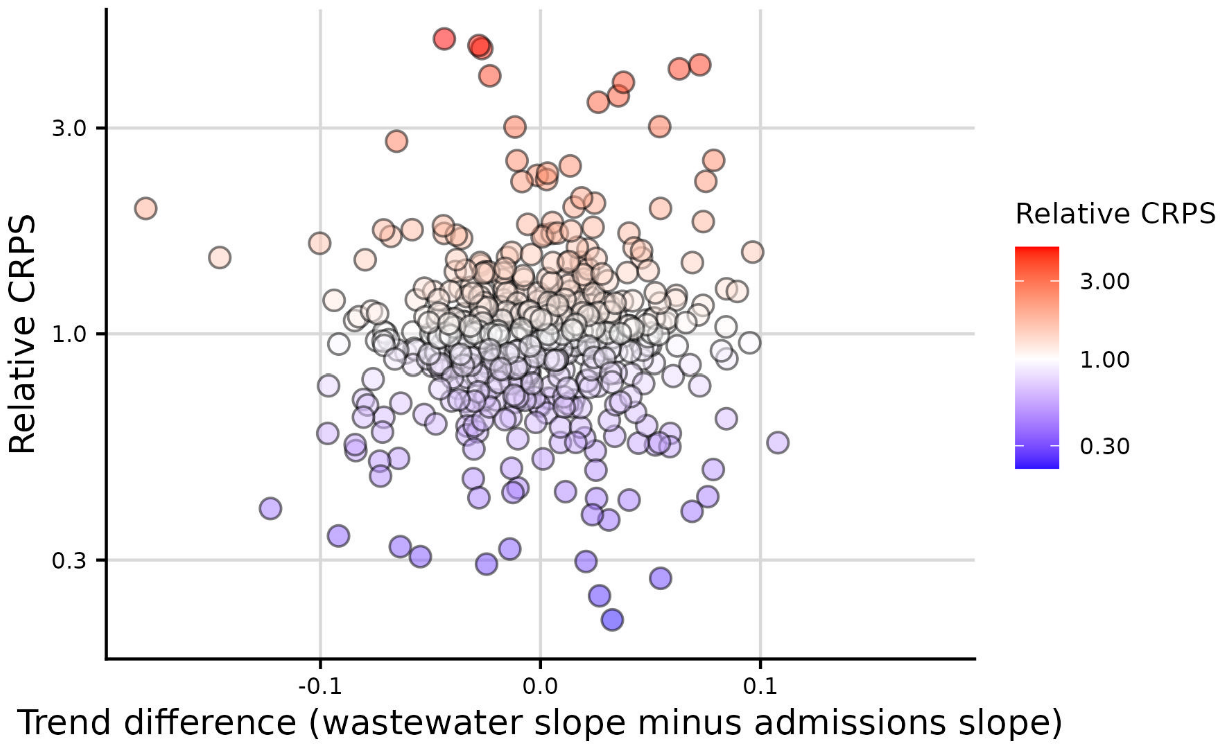


Fig. S25 Relative CRPS between the wastewater-informed model relative to the hospital admissions-only model versus the difference in the log-linear estimate of the slope between the wastewater concentrations across sites and the hospital admissions only trend. Larger differences on the x-axis indicate a greater magnitude in the difference between the two signals. Points are posterior median estimates.

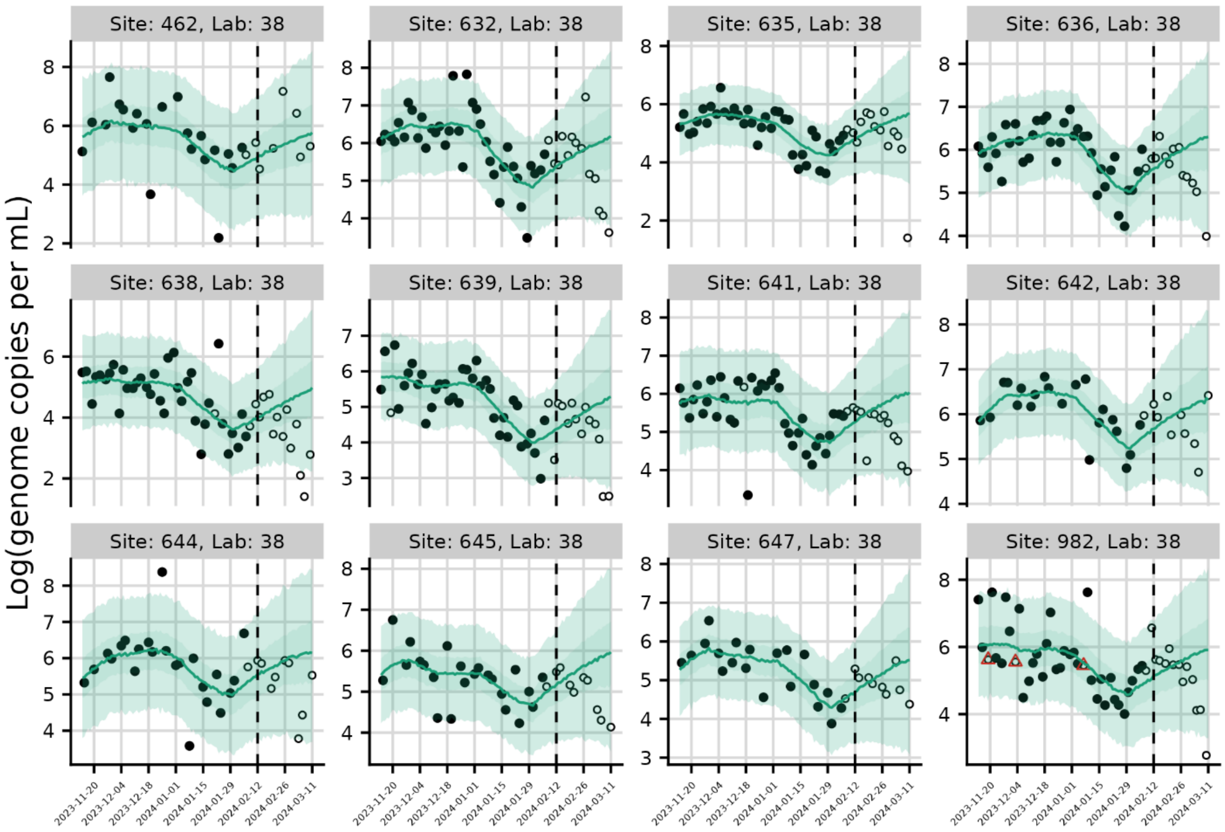


Fig. S26 Example retrospective wastewater concentration fit and forecasts from February 12, 2024 in Illinois. Red triangles indicate points that were flagged as outliers.

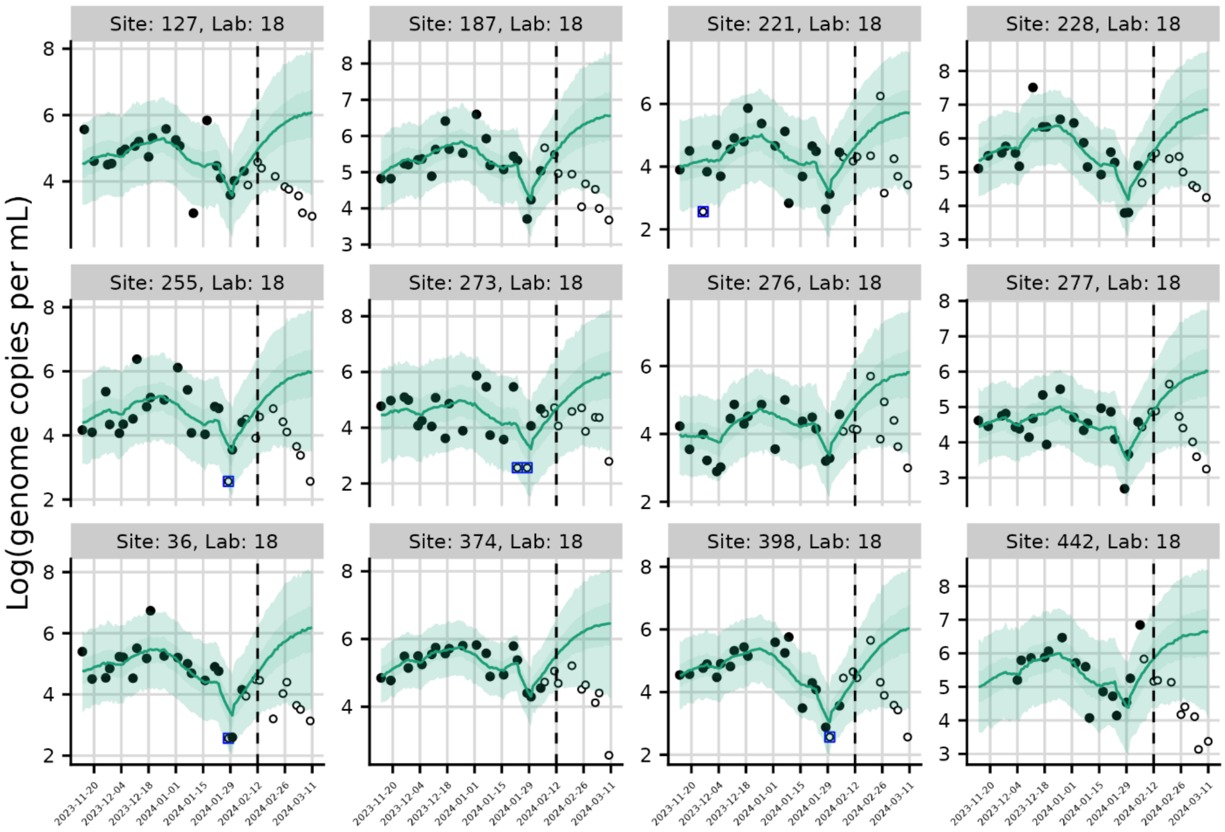


Fig. S27 Example retrospective wastewater concentration fit and forecasts from February 12th, 2024 in Ohio. Blue squares indicate observations below the limit of detection.

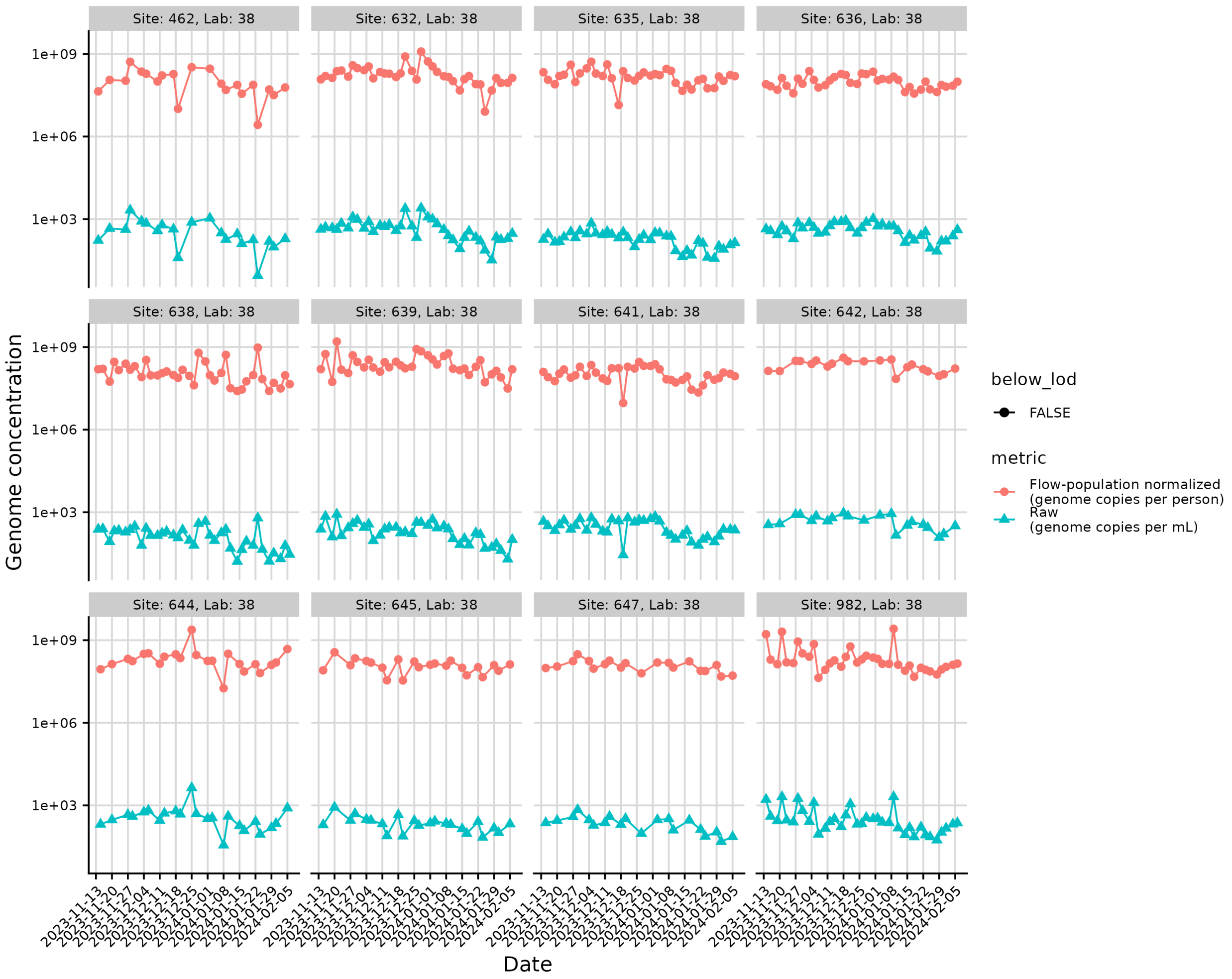


Fig S28 Comparison between raw wastewater genome concentrations (blue triangles) and flow-population normalized concentrations (red circles) wastewater data for Illinois as of the February 12, 2024 forecast date. Points for which the raw concentration was below the limit of detection are translucent, others are solid. Site lab combinations are the same as those shown in Fig S15. Raw concentrations are in units of genome copies per mL. Flow-population normalized concentrations are in units of genome copies per person. y axis is log scale.

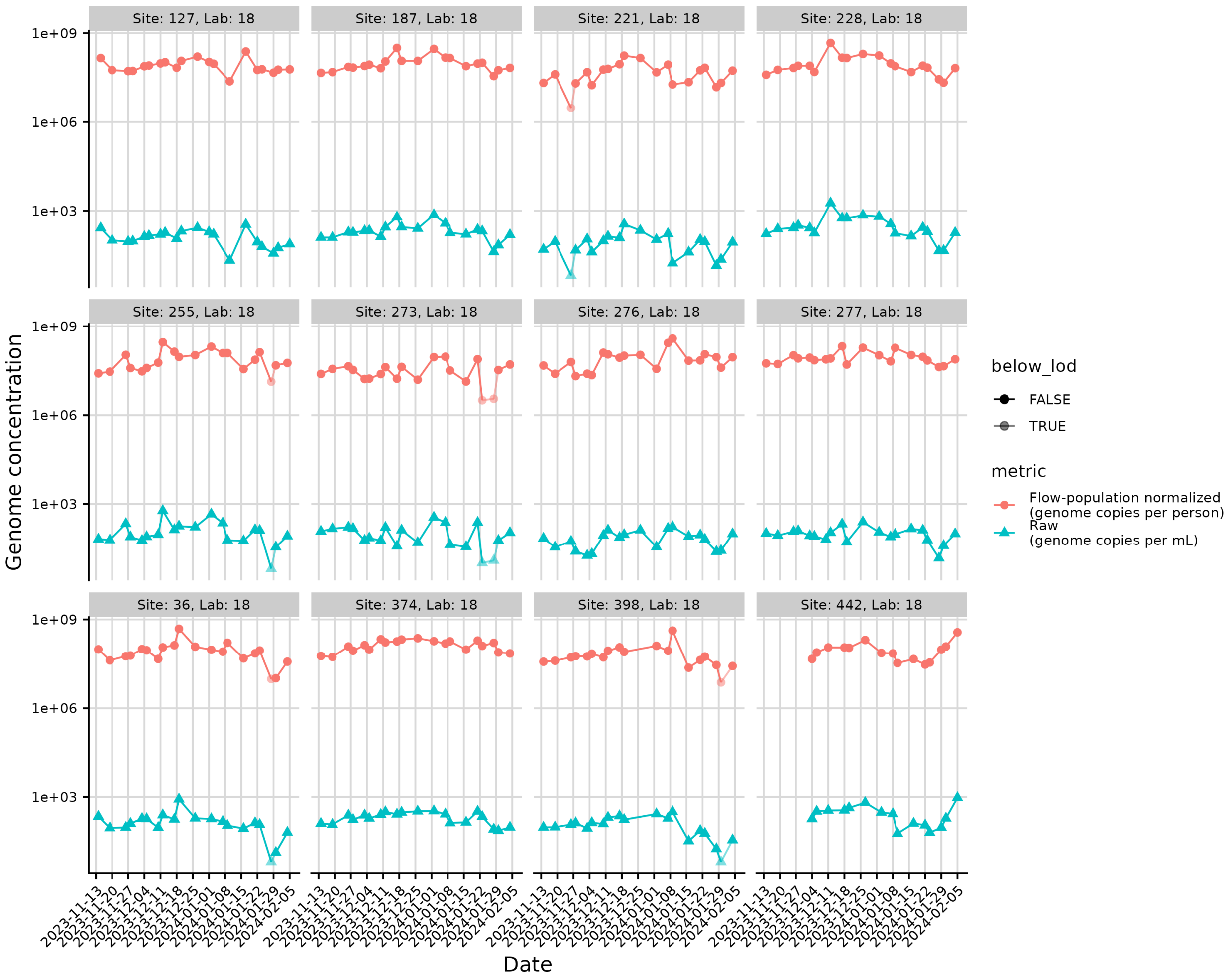


Fig S29 Comparison between raw wastewater genome concentrations (blue triangles) and flow-population normalized concentrations (red circles) wastewater data for Ohio as of the February 12, 2024 forecast date. Points for which the raw concentration was below the limit of detection are translucent, others are solid. Site lab combinations are the same as those shown in Fig S16. Raw concentrations are in units of genome copies per mL. Flow-population normalized concentrations are in units of genome copies per person. y axis is log scale.
